# Estimating the state of the Covid-19 epidemic curve in Mayotte during the period without vaccination

**DOI:** 10.1101/2021.12.30.21268571

**Authors:** Solym Manou-Abi, Yousri Slaoui, Julien Balicchi

## Abstract

We study in this work some statistical methods to estimate the parameters resulting from the use of an age-structured contact mathematical epidemic model in order to analyze the evolution of the epidemic curve of Covid-19 in the French overseas department Mayotte from march 13, 2020 to february 26,2021. Using several statistic methods based on time dependent method, maximum likelihood, mixture method, we fit the probability distribution which underlines the serial interval distribution and we give an adapted version of the generation time distribution from Package R0. The best-fit model of the serial interval was given by a mixture of Weibull distribution. Furthermore this estimation allows to obtain the evolution of the time varying effective reproduction number and hence the temporal transmission rates. Finally based on others known estimates parameters we incorporate the estimated parameters in the model in order to give an approximation of the epidemic curve in Mayotte under the conditions of the model. We also discuss the limit of our study and the conclusion concerned a probable impact of non pharmacological interventions of the Covid-19 in Mayotte such us the re-infection cases and the introduction of the variants which probably affect the estimates.

## 1 Introduction

Following the emergence of the Covid-19 and its spread outside of China, Europe and now the all world are experiencing the pandemic. The COVID-19 virus has caused a great disruption to the human health, social life, developments, and economics. In response, several countries have implemented unprecedented non-pharmaceutical interventions including case isolation of symptomatic individuals and their contacts, the closure of schools and universities, banning of mass gatherings and some public events, and widescale social distancing including local and national lockdowns of populations with all but essential internal travel banned. Also in response to the rising numbers of cases and deaths, and to maintain the capacity of health systems to treat as many severe cases as possible, France like European countries and other continents, have implemented some process measures to control the epidemic including his overseas department outside the metropolitan France namely in Mayotte island. To understand trends in the development of the epidemic in Mayotte from march 13,2020, to february 26, 2021 we explore the estimation of important parameters in a justified mathematical model. To this end, we describe the evolution of an individual through several possible states: susceptible, exposed, infected including symptomatics and asymptomatics cases and removed or isolated. Mathematical models can be defined as a method of emulating real life situations with mathematical equations to expect their future behavior. In epidemiology, mathematical models are relevant tools in analyzing the spread and control of infectious diseases. Many mathematical models of the COVID-19 coronavirus epidemic have been developed, and some of these are listed in the following papers [10, 13, 14, 6, 7]. Based on the development of epidemiological characteristics of COVID-19 infection, a model of type SEIR can be appropriate to study the dynamic of this disease including some statistical method to estimate the associate parameters.

In this paper, we first introduce the observed data in Mayotte and the mathematical tools that justified the modeling procedure in section2. In section 3 we estimate the serial interval distribution by several methods and the best-fit is given by a mixture model. We also derive an estimation of the generation time distribution. Section 4 provide numerical simulation of the effective reproduction number and Section 5 deal with the numerical approximation of the epidemic curve under some conditions. The results are presented and a discussion on the limit of our study in the last section.

## 2 Data and tools for the mathematical analysis

This work is concerned with some statistical analysis of the dynamic of the Covid-19 epidemic curve in Mayotte over a given period. Note that the observed epidemic in Mayotte started at march 13,2020 where the first detected and reported case travelled from the metropolitan France three days ago. Some others imported cases were confirmed later and lead to the actual state of the spread of the virus. In this paper we use a subset of the database from march 13,2020 to february 26,2021 for simplicity. This horizon time is chosen in such a way to justify the following fact: given that the starting of the vaccination around January, 25 and since it took on average of 3 or 4 weeks for a complete vaccination, this corresponds approximately at this end of study date. In addition, it coincided with the start of the second lockdown in Mayotte.

Note that data relating to covid-19 can be downloaded from https://www.data.gouv.fr/fr/pages/donnees-sante/. The following pictures illustrate the daily and cumulative reported number of confirmed infected cases as well as confirmed hospitalization, intensive care and death cases in Mayotte the above interval date.

**Figure 1:**
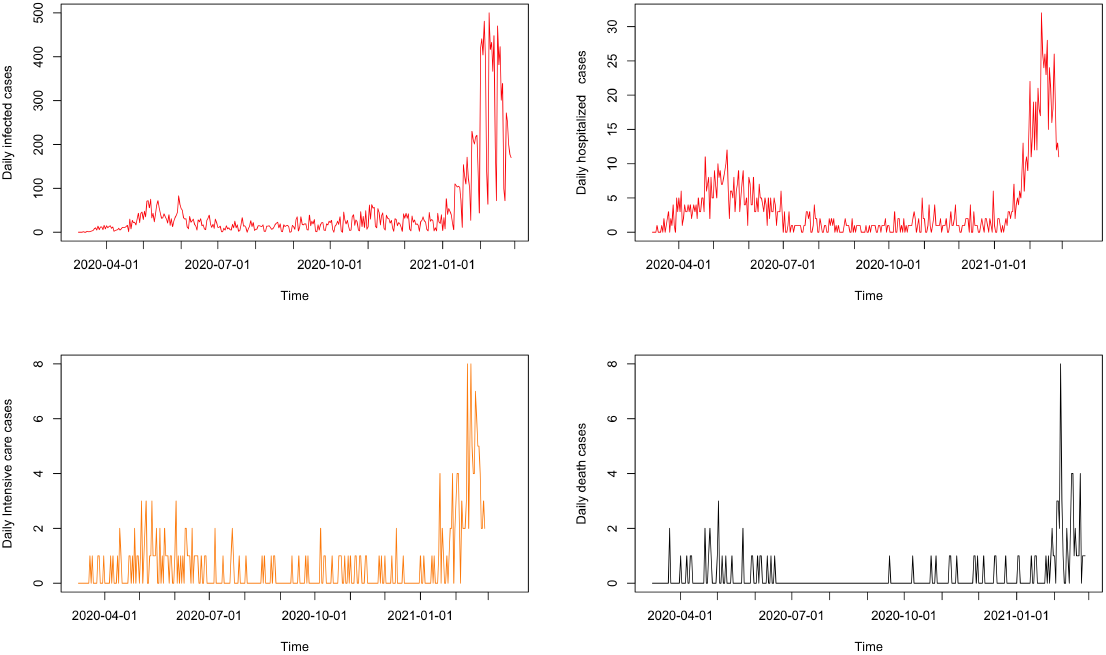
Reported daily cases of Covid-19 epidemic in Mayotte from march 13, 2021 to february 26, 2021

**Figure 2:**
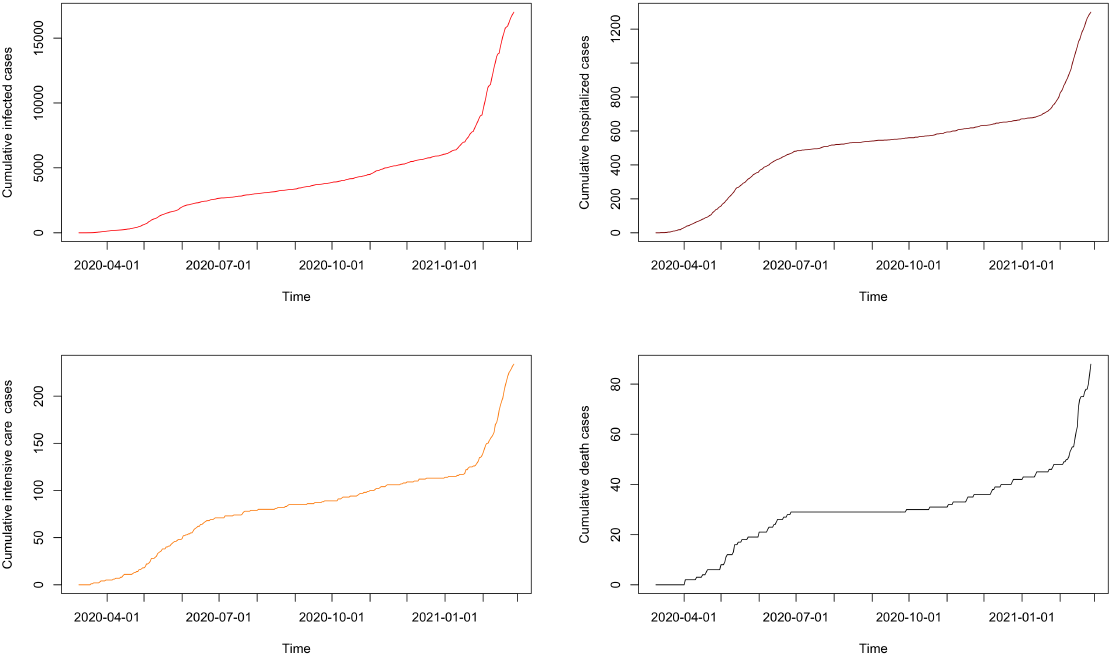
Cumulative cases of Covid-19 epidemic in Mayotte from march 13, 2021 to february 26, 2021

### 2.1 Statistical analysis by regression

Let *H*(*t*), *ICU*(*t*) and *D*(*t*) denote respectively the number of Hospitalized, Intensive care unit and Death cases at time *t*. Regression is a predictive statistical approach for modeling relationship between a dependent variable say *Y* with a given set of observed variables say *X* = (*X*_1_, *X*_2_, …, *X*_*n*_) in the form

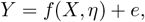

where *η* is unknown (vector) parameter and *e* representing an additive error that may stand for a white noise.

Now, let us analyse in the following lines the performance of some relevant regression linear model

- 90.84% of the variations in the cumulative intensive care *ICU*(*t*) are explained by the variations of t the cumulative infected cases *I*(*t*) with a coefficient around 0.013 and CI equal to [0.012, 0.013] with a p-value *<* 2 ∗ 10^−16^ and the constant around 2.78.
- 88.89% of the variations in the cumulative death *D*(*t*) are explained by the variations of the explanatory variable *I*(*t*) with a coefficient around 4.42.10^−3^ and CI equal to [0.0042, 0.0045] with a p-value *<* 2.2 ∗10^−16^ and a negligible effect of the constant.
- 86.88% of the variations in the cumulative hospitalized cases *H*(*t*) are explained by the variations of the cumulative infected cases *I*(*t*) with a coefficient around 0.07 with a CI equal to [0.068, 0.07] with a p-value *<* 2.2 ∗ 10^−16^ and an important effect of the constant (intercept).

Based on the above analysis, we summarize the obtained predictions in the following pictures.

**Figure 3:**
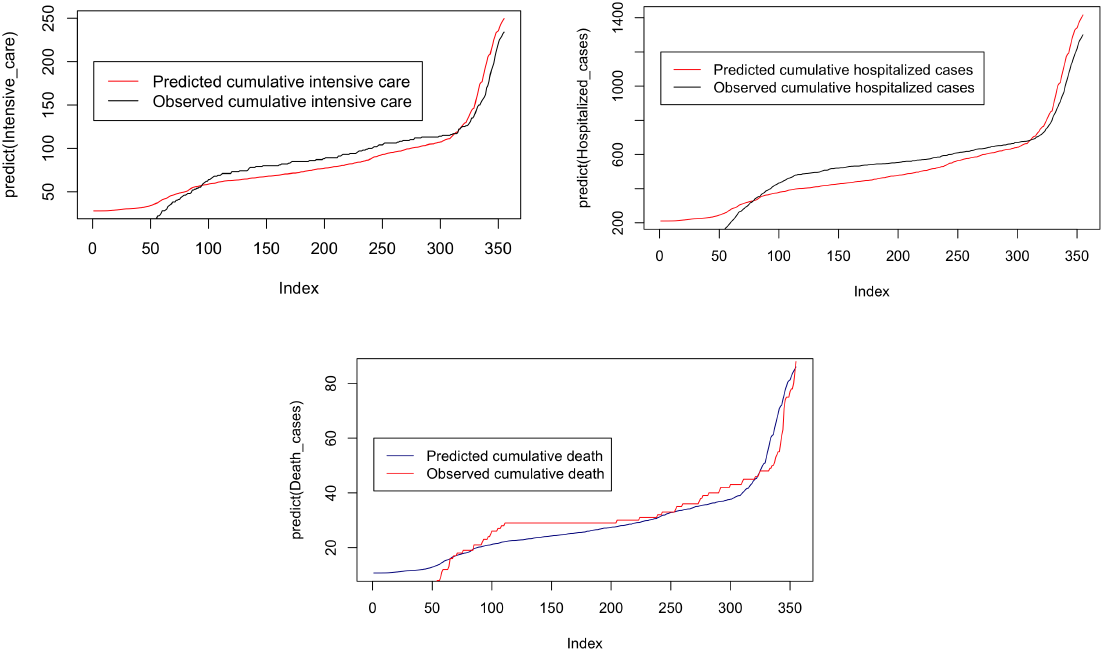
Predicted cumulative cases by linear regression

One can observed in the above pictures that the predicted cumulative cases based on the linear model is not better at certain points in time evolution. This may be due to some non-linear variations. In order to do better, let us deal with a fit based on Generalized Additive Model (GAM) which allows the to use of mix nonlinear, linear functions and categorial effects to data without overfitting. With a GAM, however, we can fit data with smooths, or splines, which are functions that can take on a wide variety of shapes. We fit a GAM using a polynomial transformation and predict the above desired cumulative cases as follows.

**Figure 4:**
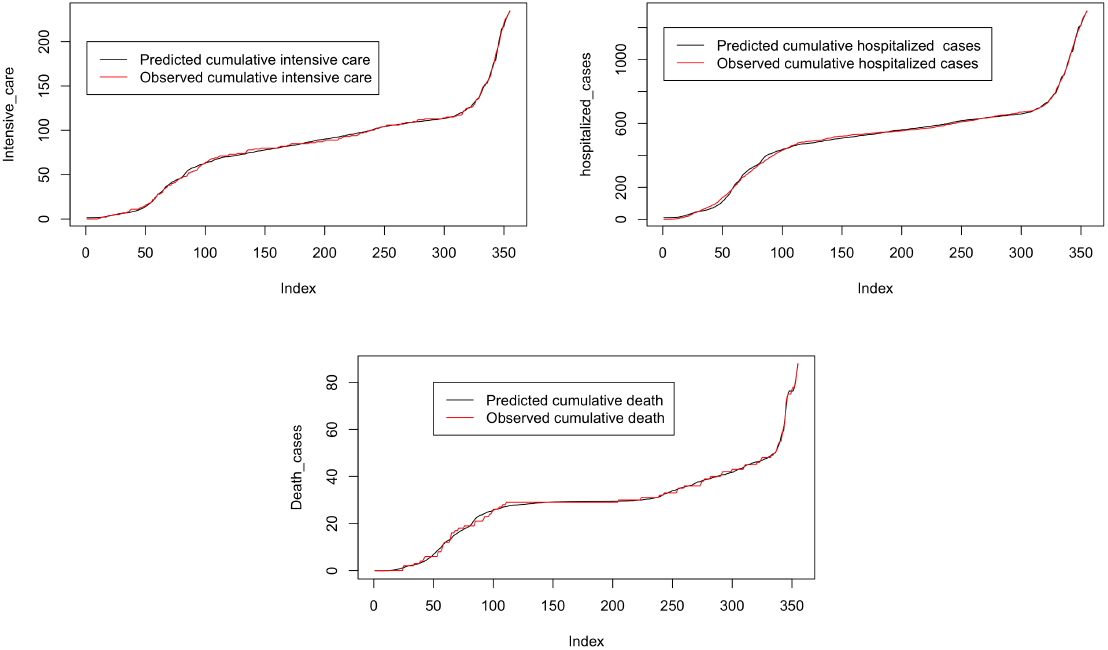
Predicted cumulative cases by GAM

### 2.2 Some definitions

Now let us specify some definitions concerning the infectivity process. Consider an infector *i* and infectee *j*.

#### Definition 1

*The generation time interval* 𝒢_*i,j*_ *is the time interval from the infection of i to infection of j*.

The generation time distribution 𝒢describes infectiousness with respect to the point of infection.

#### Definition 2

*The serial interval time 𝒮*_*i,j*_ *is the time interval between symptom onset of the infector i and symptom onset of the infectee j*.

#### Definition 3

*The incubation time interval ℰ is the time interval between symptom onset and the infection*.

The infectious profile denoted by 𝒫_*i,j*_ is the time interval from the symptom onset of *i* to infection of *j*. It describes infectiousness relative to symptom onset. The above distributions are typically derived from contact tracing and can be captured by the following equations:

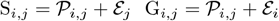

One can assume that *𝒫*_*i,j*_ and *ℰ*_*j*_ are plausibly independent as well as *ℰ*_*j*_ independent of *ℰ*_*i*_ so that *𝒮* = *𝒫* ∗ *ℰ*. Note that the incubation and serial interval distributions can be estimated without further assumptions but not the generation time and the infectious profile. We refer the reader to [11, 22] for further explanations as concerns the way to derive the generation time distribution from the serial interval.

### 2.3 Infection model from the Markovian SIR and SEIR

To model the number of infections over time, we need to understand how the above transmission distributions appeared in a mathematical modeling. In the context of population dynamics, let us recall the following formation of Lotka and Euler in [15, 5]:

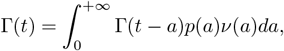

where Γ(*t*) is the number of births at time *t, p*(*a*) is the probability of survival to age *a* and *ν*(*a*) is the fertility at age *a*. The above renewal equation was adapted to epidemics in the seminal work of Kermack and Mc Kendrick in [9]. To understand the above renewal equation in an epidemiological context, let us consider firstly the following simple Markovian SIR model in a closed population with size *N* partionned into Susceptible individuals *S*, Infectious individuals *I* and Recovered individuals *R*. We consider only transmission from person-to-person and we assume that infectious individuals become permanently immune after their infectious period during our horizon time study *T* . Of course in the case of Covid-19 we do not have permanent immunity.

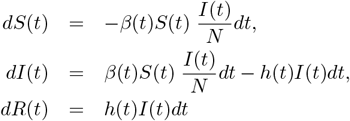

where *h*(*t*) is the hazard rate function associated to the survival function *G*(*t*) of the time infectivity. The time varying transmission rate is denoted by *β*(*t*). For simple computations, we assume that *h*(*t*) is constant and equal to *γ* the mean rate of the infectivity (being exponentially distributed with parameter *γ*) so that *G*(*t*) = *e*^*−γt*^. The second equation of the above system tell us that

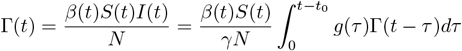

where *t*_0_ is the starting date of the epidemic and *g*(*τ*) = *γe*^*−γτ*^ is the density of the probability distribution function of the secondary infections according to primary-infection: the so called generation time distribution (assumed to be exponentially distributed). The above formula means that new susceptible individuals are reported at time *t* according to the contact rate *β*(*t*)*γ*^*−*1^ if they were infected before time *t* and remains infectious for the duration *τ* until date *t* with probability *g*(*τ*). Set

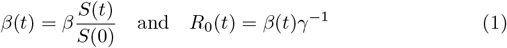

At the beginning of the epidemic *t* = *t*_0_, we obtain the formula for the basic reproduction number at the beginning of the outbreak:

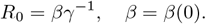

assuming that *S*(0) ∼*N* .

We have

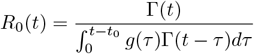

Therefore in a discrete time setting we have the following algorithmic formula for all *t > t*_0_:

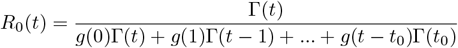

The following fact is easy to verify

#### Proposition 1

*In the above Markovian SIR model if γ* ∈ [*γ*_*min*_, *γ*_*max*_] *then in a discrete time setting we have*

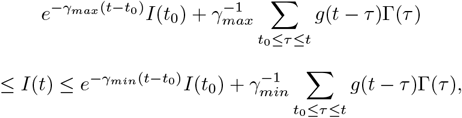

*for all t* ≥ *t*_0_ + 1.

We illustrate the above proposition as follow with *γ*^*−*1^∈ [7, 21] days.

The above Markovian SIR model under learn the estimation of *I*(*t*), this was predictable since it does not take into account the delay in the infectivity process due to the incubation.

Let is us consider a Markovian SEIR model where the population is partitionned into five categories : Susceptibles *S*; Exposed *E* infected but not yet infectious in a latent period with mean *λ*^*−*1^; Infectious *I* being reported or unreported with an infectious period with mean *γ*^*−*1^, Dead *D* and Recovered *R*.

The model meets the following assumption.

**Figure 5:**
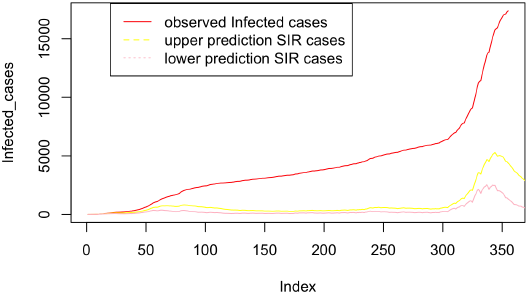
Estimated (lower and upper) cumulative infected cases with a markovian SIR model

- The natural average mortality rate *µ* and a recruitment rate Λ can be considered but we shall assume that the recruitment rate Λ compensates succeptibles individuals who died:

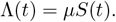

Given initial conditions *D*(0) = *D*_0_ and

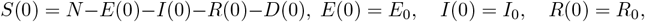

where *N* is the total fixed population size, the model consists of the following system of differential equations

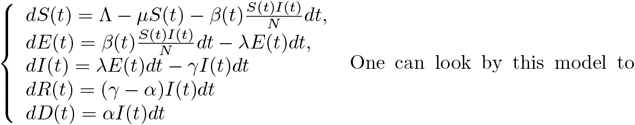

keep track of the cumulative cases from the time of onset of symptoms *C*(*t*) which is not a compartment and defined as *dC*(*t*) = *λE*(*t*).

Assume that the probability of a susceptible to be alive at time *t >* 0 given that we was alive at time *t* = 0 follows an exponential distribution of parameter *µ*. Assume also that the incubation and the infectiousness are independent and follows respectively an exponential distribution with parameter *λ* and *γ* and let *G*_*λ*_, *G*_*γ*_ and *G*_*µ*_ their associate survival functions.

The following proposition follows from the disease equations.

#### Proposition 2

*Under the above assumption, we have in the Markovian SEIR model* :

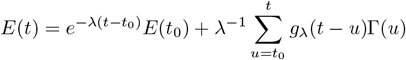

*and*

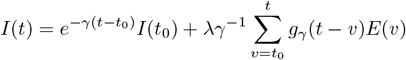

*so that if γ ≠ λ we have*

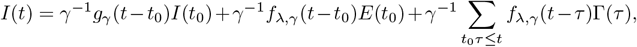

*where* 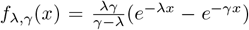 *is the density distribution of the two parameter*(*λ, γ*) *hypo-exponential distribution*.

**Figure 6:**
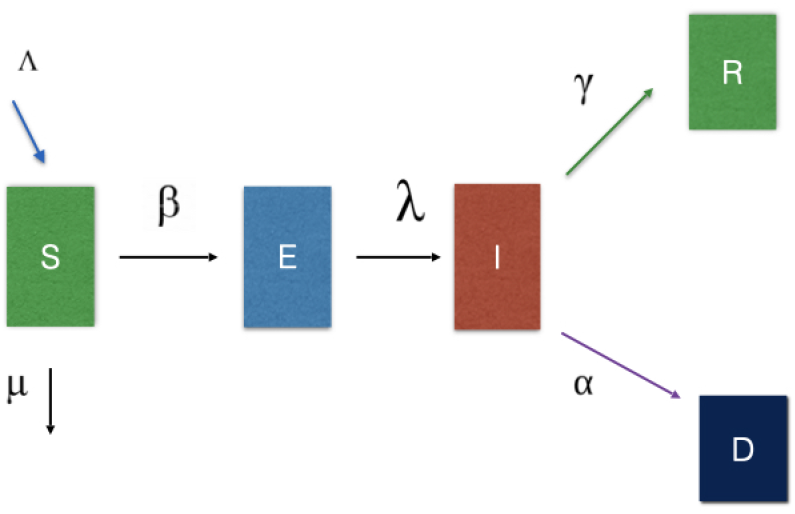
Compartments of the SEIRD model

We illustrate the above proposition as follow with *γ*^*−*1^ ∈ [7, 21] days and *λ*^*−*1^ ∈ [1.5, 7] days

**Figure 7:**
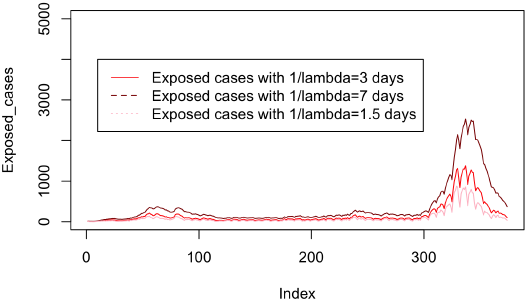
Estimated cumulative exposed cases with *E*(0) = 15 in a markovian SEIR model with various *λ*

**Figure 8:**
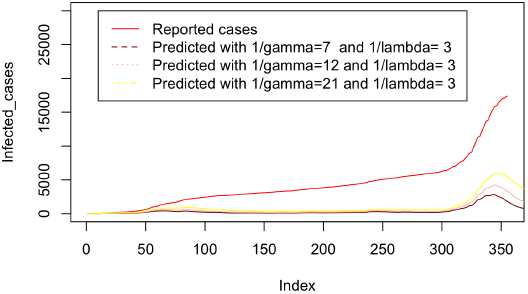
Infected cumulative cases with *E*(0) = 15 in a markovian SEIR model with various *γ* and fixed *λ*

**Figure 9:**
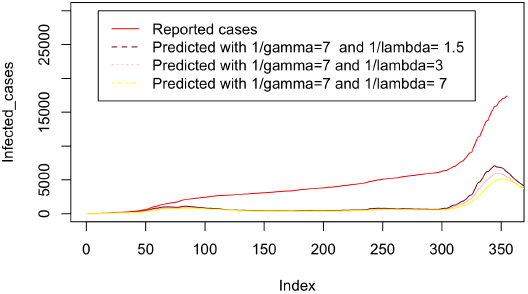
Infected cumulative cases with *E*(0) = 15 in a markovian SEIR model with various *λ* and fixed *γ*

Here again the Markovian SEIR model under learn the estimation of *I*(*t*).

This is due to the fact that the generation times are not exponential as we will see in the next section. For a deeply study of non-Markonvian SEIR model we refers the reader to the work of [6, 7].

## 3 Estimation of the serial interval and generation time distributions

Some major studies have been made on the determination of generation times through serial interval distributions and incubation time from real data collected, see for instance [8, 17]. For the rest of this paper, we will focus on the data collected in [8] where the authors report temporal patterns of viral shedding in 94 patients with laboratory-confirmed Covid-19 and modeled Covid-19 infectiousness profiles from a separate sample of 77 infector–infectee transmission pairs. The variation between individuals (infector/infectee) and the transmission process is summarized respectively by the distribution of incubation periods and the distribution of serial intervals. The database used contains the number of infected cases (in red), the lower limit of infectors (in black) and the upper limit of infectors (in green). The variation between individuals and chains of transmission is summarized respectively by the distribution of incubation periods and the distribution of serial intervals.

**Figure 10:**
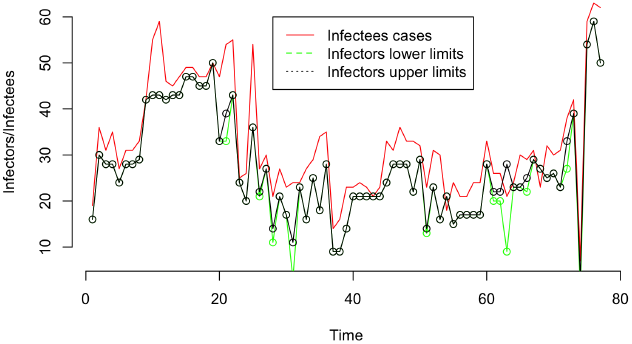
Evolution of the infector/ infectee pairs

The main purpose in this section is to study the above data of transmission pairs in order to be able to fit the best distribution of the serial interval which is computed as follow like in[8]:

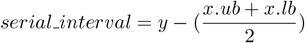

with

– *x*.*lb* : Lower limit dates of onset of symptoms of infectors
– *x*.*ub*: Upper limit dates of onset of symptoms of infectors
– *y*: Dates of onset of symptoms of the infectee.

The incubation period is calculated by the formula

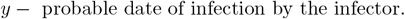

The following picture show the histogram of the serial interval from the underline database.

**Figure 11:**
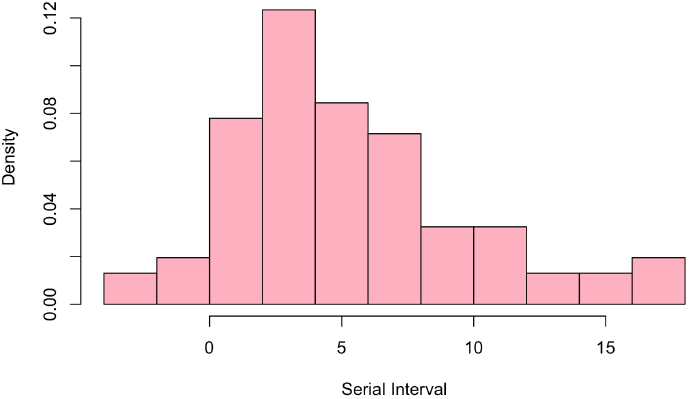
Histogram of the serial interval

Now, we will attempt to fit a theoretical distribution to the above histogram of the observed empirical distribution function of the serial distribution among a set of distributions that we will choose. A rapid analysis of the histogram seems to show that the serial interval can be fit by a family of exponential distribution or a mixture of them because of the appearance of several peaks. There are many methods for generating estimates of parameters for standard probability distributions such as the method of moments and the Maximum Likelihood.The use of others methods was generally confined to analytical work on mixture densities.

### Estimation of the serial interval distribution by the method of moments

In this section we use the method of moments amoung a set of classical laws. The method of moments consists in estimating the parameters of the distributions from the theoretical finite moments with their empirical estimates. The results of this method can be seen in the following picture.

**Figure 12:**
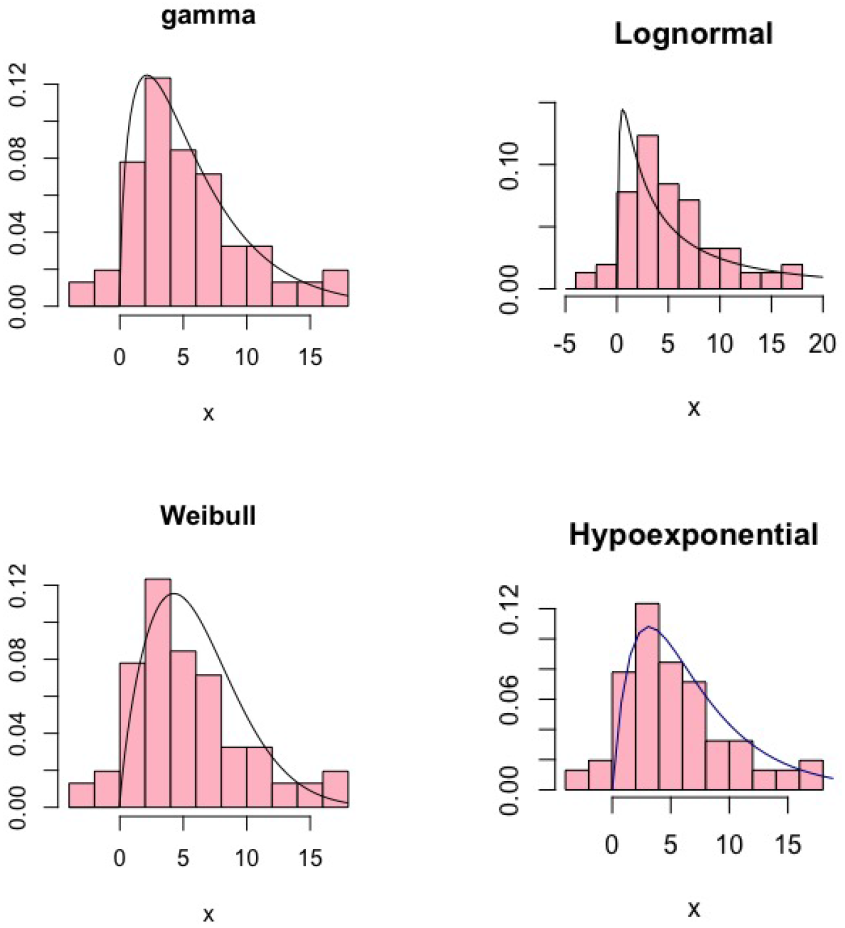
Fitted distributions with the method of moments

The above pictures mentioned namely fitted:

– Gamma distribution with parameters 1.584523 and 0.2735611
– Lognormal distribution with parameters 1.756514 and 1.526372
– Weilbull distribution with parameters 1.756514 and 6.87615
– Two parameter Hypo-exponential (denoted by *exp conv*) distribution with parameters 1*/*4.9 and 1*/*2.1

Note that the standard exponential distribution (here with parameter 0.1726457) does give a good fit, as it can be seen in the following picture.

**Figure 13:**
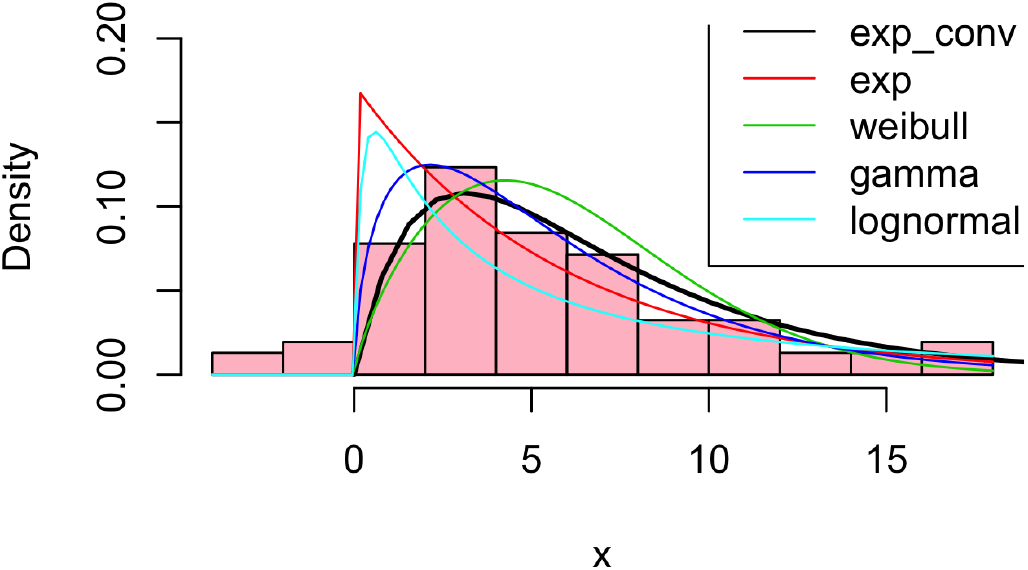
Summary (moment method) of fitted distributions.

As we can see, the gamma, weibull seems to matche if we do not take into account the left tails of the distributions. Using the Kolmogorov-Smirnov non parametric test with a risk *α* = 0.05 gives an p-value around 0.5229, 0.5237 ∗10^−9^ and 6.312572 ∗10^−4^ for Gamma, Weibull and Lognormal distribution respectively.

Note that the method of moments does not give best fit and this can be explained by the fact that this basic method does not use any optimization techniques as the following Likelihood method.

#### 3.1 Estimation of the serial interval distribution by the Likelihood method

Here we consider the maximum likelihood estimator which is a statistical method used to estimate the parameters of the probability law of a given observation *x* = (*x*_1_, .., *x*_*n*_) with probability densities *f*(*x*_*i*_| *θ*) by determining the values of the parameters maximizing the log-likelihood function given by :

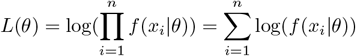

where *θ* is the (vector) parameter to be estimate. The likelihood function then represents the joint density of the individual observations *x*_*i*_ for any given level of the distribution parameters. The maximum likelihood estimate is the distribution parameter values, which maximize the likelihood function since these same values of the estimators also maximize the log-likelihood function. The *nlm* function of R-Package in [18] makes it possible to optimize the log likelihood functions of the different laws that we pass as an argument in addition to the initialization of the parameters to be optimized and of the serial interval that we seek to determine its better distribution. The following picture represents several fitted densities estimated by this method.

**Figure 14:**
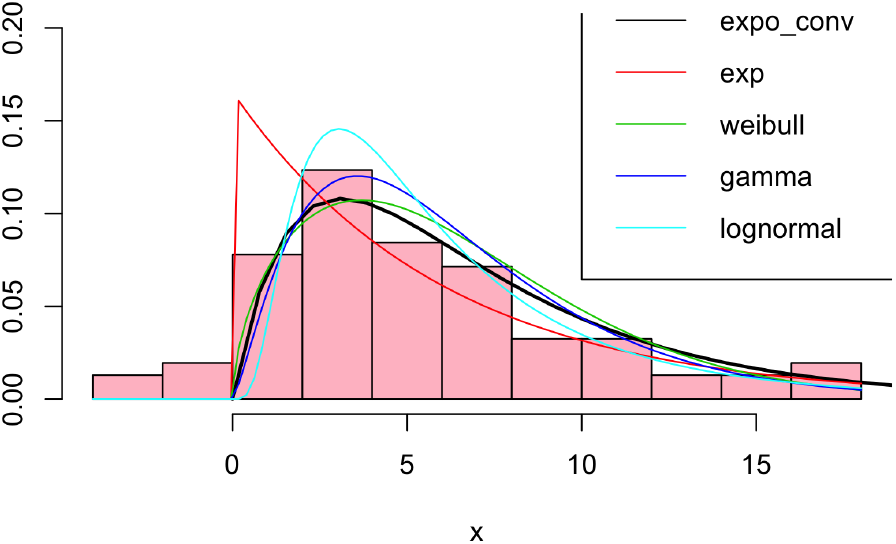
Estimation of distributions by the maximum likelihood method

**Table 1:**
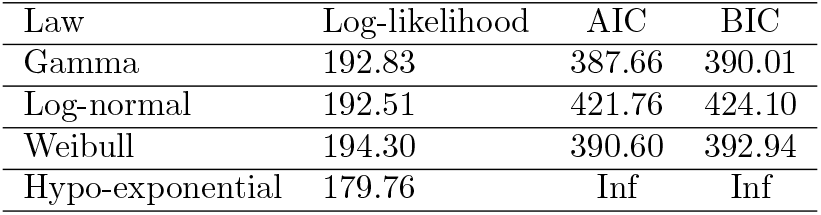
Log-likelihood, AIC and BIC values

We can see that compared to the method of moments, the Maximum Likelihood improve in some way the estimation. The log-likelihood values and widely applicable information AIC and BIC criteria is summarise in the following table

We note that that the gamma, log-normal and weibull models are the best to fit data. In addition one can see that the two parameter hypo-exponential distribution does not converge, which explains why the estimated parameters are negative.

**Figure 15:**
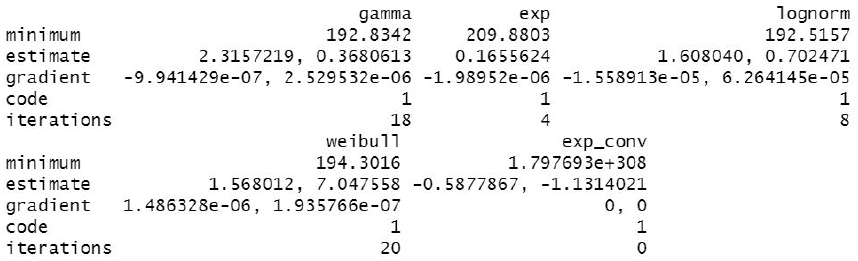
Maximum likelihood method results

Note also that the left tails of the distributions don’t matches and since the underline left tail data represents 7% of the data we neglect this part of the data. Another upcoming work will deal with the theoretical aspects of estimating heavy tails of distribution that may be appropriate for doing cross mixtures of heavy tails and exponential family laws.

#### 3.2 Estimation of the serial interval distribution by mixture methods

Now let us deal with mixture modeling with exponential families. Mixture models are a powerful and flexible tools (see [16, 2]) to model an known smooth probability density function as a weighted sum of parametric density functions *f*_*j*_(*x*|*θ*_*j*_) in a possibly multivariate independent observations *x* = (*x*_1_, .., *x*_*n*_) drawn from *k* several densities :

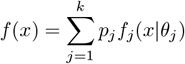

where *k* represents the number of classes or groups of the mixing, *p*_*j*_ *>* 0 the relative proportion of observations from each class *j* such that 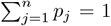 . The mixture density will allow to find the parameter Θ = (*p*_*j*_, *θ*_*j*_) of the model where *θ*_*j*_ is the (vector) parameter from class *j*.

The log-likelihood of the mixing density is defined by:

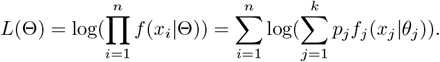

The maximum likelihood estimates are:

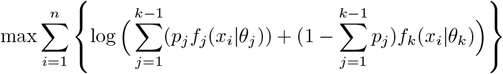

subject to

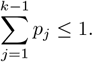

For *k >* 1, the sum of term appearing inside a logarithm makes this optimization quite difficult. Solution of this non-linear optimization problem has long been a difficult task for applied researchers. There are, of course, many general iterative procedures which are suitable for finding an approximate solution of the likelihood equations and which have been honed to a high degree of sophistication within the optimization community. For the data, a two-component mixture model is clearly a reasonable model based on the bi-modality evident in the histogram.

Using the R software function **optim**, it possible to minimize the log likelihood functions of the different laws taken as arguments of this function in addition to the initialization of the parameters to be optimized and the serial interval distributions we want the best-fit. The figures below summarize the estimation results of the mixture models. Gamma mixing, lognormal and weibull mixing seem to give better results compared to the others.

**Figure 16:**
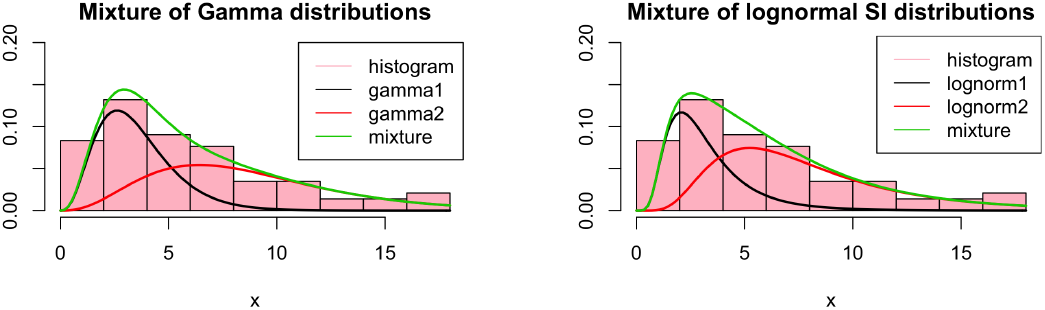
Mixture distributions by the Optim method

**Figure 17:**
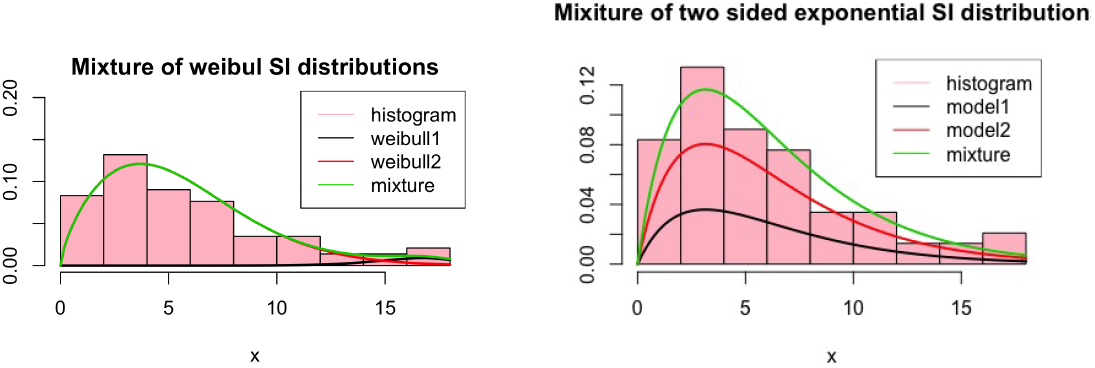
Mixture distributions by the Optim method

The Kolmogorov-Smirnov test, log-likehood, AIC and BIC values is summarised in the following table We conclude that the best-fit mixture is given by *p* = 0.0406 and two Weibull distributions with parameters (9.96140450, 16.90693027) and (1.70447135, 6.20140484).

We can see the visualization of the results of the mixtures of each law as follows.Note that one can still go further in the estimation by a cross-mixture among the best three-fit distributions and the a heavy tail distribution by the use of the EM algorithm. This will be done in another forthcoming theoretical and practical work.

**Table 2:** Kolmogorov-Smirnov test, log-likehood, AIC and BIC values

| Law | P-value | alpha | Log-likelihood | AIC | BIC |
| --- | --- | --- | --- | --- | --- |
| Gamma | 0.686 | 0.05 | 192.06 | -374.12 | -362.40 |
| Log-normal | $6.65 \cdot 10^{-4}$ | 0.05 | 191.90 | -373.81 | -362.0935 |
| Weibull | $1.16 \cdot 10^{-16}$ | 0.05 | 191.60 | -373.21 | -361.49 |

**Figure 18:**
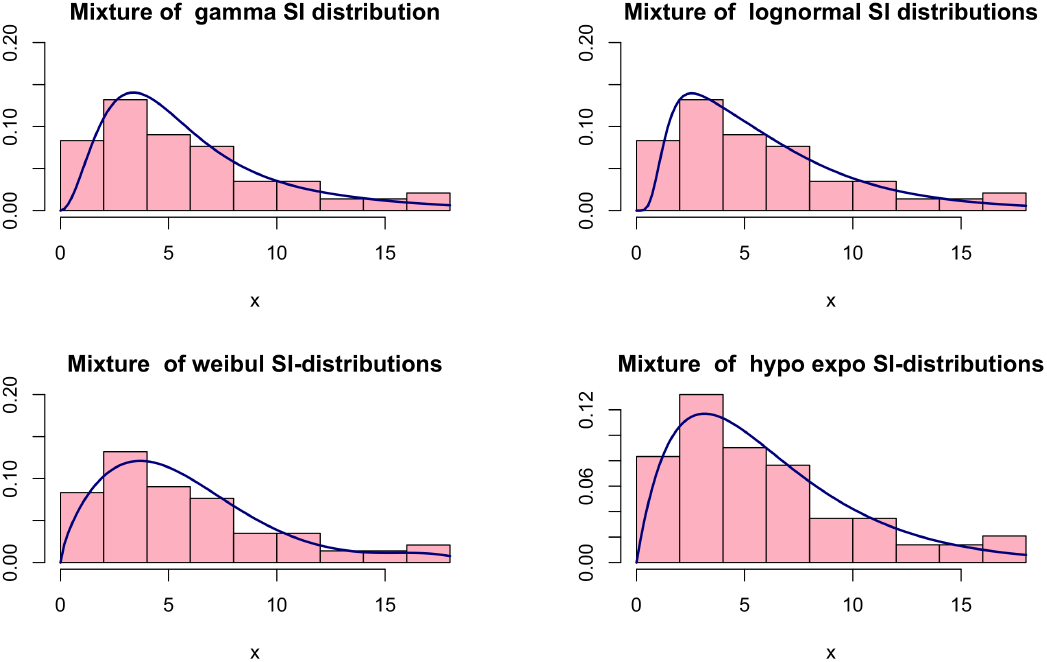
Result of each mixture density distribution

#### 3.3 Results of the estimation on the incubation and infectious time distribution

Let us summarise here some well known works about the estimation of the infectiousness and incubation period of [12]. In this work, the authors established that the best fit bincubation period distribution is of mean 5.2 days with 95% CI equal to [4.1, 7]. Using statistical estimation method, the authors also show that the best-fit distribution of this incubation rate is managed by a log normal distribution of parameters 1.434065 and 0.6612.

Their analysis also suggests that viral shedding may begin 5 to 6 days before the appearance of the first symptoms and inferred that infectiousness started from 12.3 days (95% CI equal to [5.9, 17] days before symptom onset and peaked at symptom onset (95% CI equal to [–0.9, 0.9] days). The authors pointed out that with sensitivity analysis, the infectiousness start from 5, 8 and 11 days before symptom onset and infectiousness was shown to peak at 2 days before to 1 day after symptom onset, and the proportion of pre-symptomatic transmission ranged from 37% to 48%.

### Estimation of the generation time distribution

Our aim here is the estimation of the generation time distribution by the use of the above best-mixture distribution result. Let us mentioned that in the package *R*_0_ (see [19, 3]), the best generation time distribution is fitted by the *generation*.*time* function, based on the epidemic incidence curve and the best-fit serial interval distribution choose from “gamma”, “log-normal” and “weibull” distributions using the Newton-Raphson optimization algorithm. In our case, we have used the Optim function to fit the best serial interval distribution, which is given for instance by the Weibull distribution (since it turns out to be the best to match the data in mixture model). Thus, using our best-fit mixture model together with the Mayotte epidemic curve, we obtain a modified generation time distribution from the Package *R*0.The result can be seen in the following picture.

**Figure 19:**
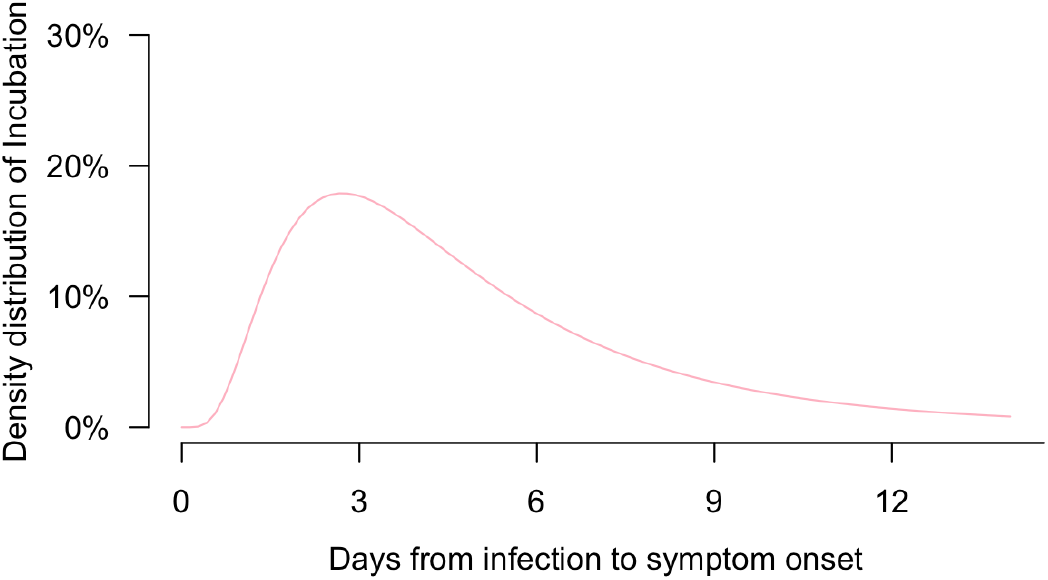
Density distribution of incubation by a best-fit log-normal distribution with parameters 1.4340 and 0.6612

**Figure 20:**
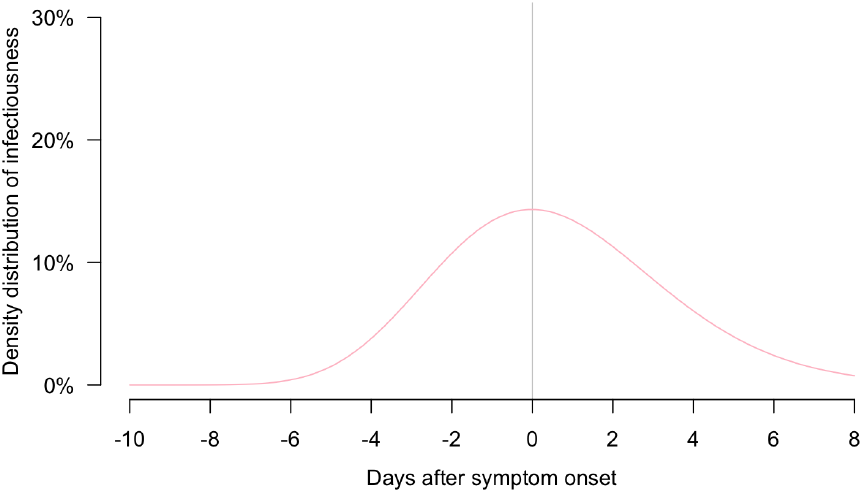
Density distribution of the infectious time distribution

**Figure 21:**
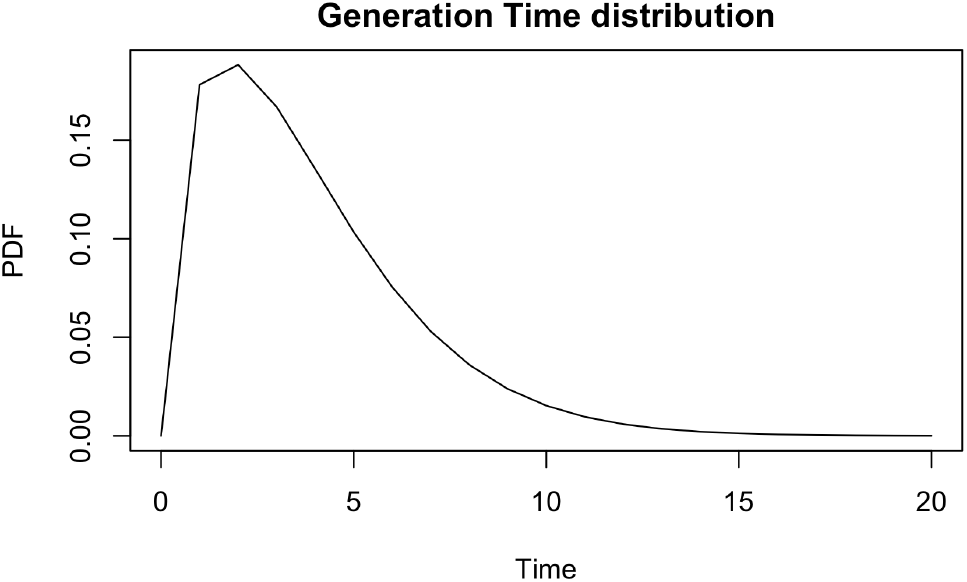
Generation time distribution by the best-fit mixture of the SI distribution (Weibull) by the optim method associated to package R0

## 4 Numerical estimates of the effective reproduction number

Understanding the development of an epidemic is important in order to control its spread. For this, various models point out the importance of the the basic reproduction number *R*_0_ at the start of an epidemic. It is historically defined as the average number of new cases of infection generated by an individual during a period of infectivity (see [4]). As stated above, if this number is less than 1 then the epidemic will tend to die out. There are several methods for computing the parameter *R*_0_ as well as its time dependent version as explained above. Let us mentioned some others parametric and non-parametric approaches. In the parametric approach, we will approach the estimation of *R*_0_ by two method based on the attack rate of an epidemic and an exponential growth rate. The non-parametric approach will also be described from several angles.

### 4.1 Estimation based on attack rate

Let us consider again the elementary Markovian SIR model. Recall that the basic reproductive number and the time varying reproductive number are defined by the relation

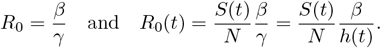

In epidemiology, the attack rate *AR* is the percentage of an at-risk population that contracts the disease during a specified time interval. The term attack rate is sometimes used interchangeably with the term incidence proportion.The attack rate is calculated as the number of people who became ill divided by the number of people at risk for the illness. if we denote by

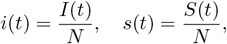

it is straighfoward to see that

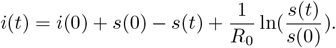

If at time *t* = 0,we assume that no one is retired then *i*(0) + s(0) = 1 and

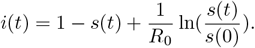

By taking the limit and setting s_∞_ = 1 – *AR* we will have:

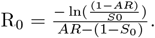

where *S*_0_ represents the percentage of the initial homogeneous and closed population assuming no intervention during the epidemic. From the relation

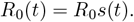

The varying attack rate is then defined by

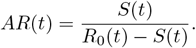

The confidence interval with respect to the rate attack is given by

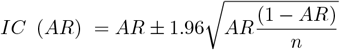

Note that this approach assumes that the transmission rate *β* is known which is not always true. This proves the limits of the parametric method.

### 4.2 Non parametric estimation based on exponential growth rate

The exponential growth rate *r* is the change per capita in the number of new cases per unit time during the early phase of an outbreak. It is an important measure for the speed of spread of an infectious disease. It being zero is, like the basic reproduction number *R*_0_ = 1, a disease threshold. The disease can invade a population if the growth rate is positive and cannot invade (with a few initially individuals) if it is negative. In fact, it can be used to infer *R*_0_. We describe here a non parametric method following J.Wallinga and M. Lipsitch in [23, 24]

Let *η* be the probability that an individual will remain infectious over a unit of time after being infected (i.e. age of infection) and *β* be the transmission rate at age of infection *a*. Then *τ*(*a*) = *η*(*a*)*β*(*a*) is the transmissibility of an infectious individual at the age of infection *a*, assuming that the entire population is susceptible. Hence

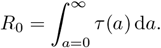

We can normalize *τ*(*a*) to be probability density function:

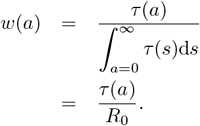

so that

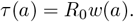

Let Γ(*t*) be the number of new infections during the time interval ]*t*; *t*+*dt*[. Note that the new infections at time *t* are the sum of all infections caused by infectious individuals infected at a unit of time (i.e. at time *t* = *a*) if they remain infectious at time *t* (with an infectious age *a*) and their contact is sensitive. In other words

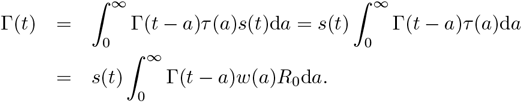

At the start of an epidemic (*t* = 0), where the epidemic grows exponentially (with an exponential growth rate *r*). *s*(*t*) ∼ 1 and Γ(*t*) = *c*_0_*e*^*rt*^ where *c*_0_ is the initial number of cases at time *t* = 0. Thus

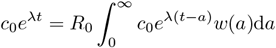

so that 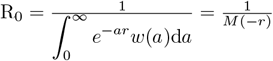 where

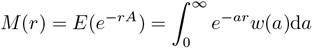

is the the moment generating function of the generation time distribution.For *t ≠* 0,

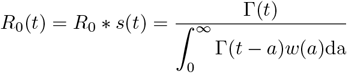

### 4.3 Non parametric estimation based on Maximum likehood

This estimation method is proposed by White and Pagano in [25] and is based on the assumption that the number of secondary cases caused by an index case follows a poison distribution with the expected value *R*. Let (*N*0, *N*1, …, *NT*) be the incident case as a function of time with a time horizon *T* and *G* a distribution of the generation time. We estimate *R* by maximizing the log-likelihood

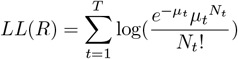

where

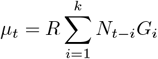

By derivation it is easy to show that

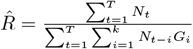

### 4.4 Numerical results of the estimation

In this section we make numerical simulation based on the observed data in Mayotte island to estimate the time varying reproduction number using not only the non parametric method based on the generation time distribution fitted by the daily observed infected cases and the best mixture of the serial interval distribution given by the Weibull distribution.

The following picture represents the evolution of the time varying reproduction number *R*_0_(*t*)

**Figure 22:**
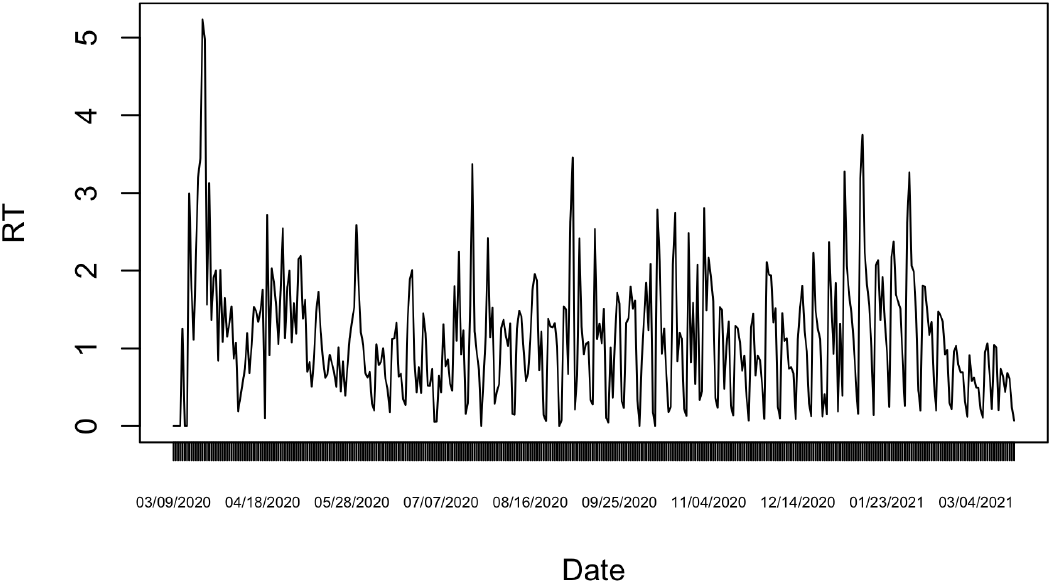
Evolution of the time varying reproductive number in Mayotte from march 13, 2020 to february 26, 2021

**Figure 23:**
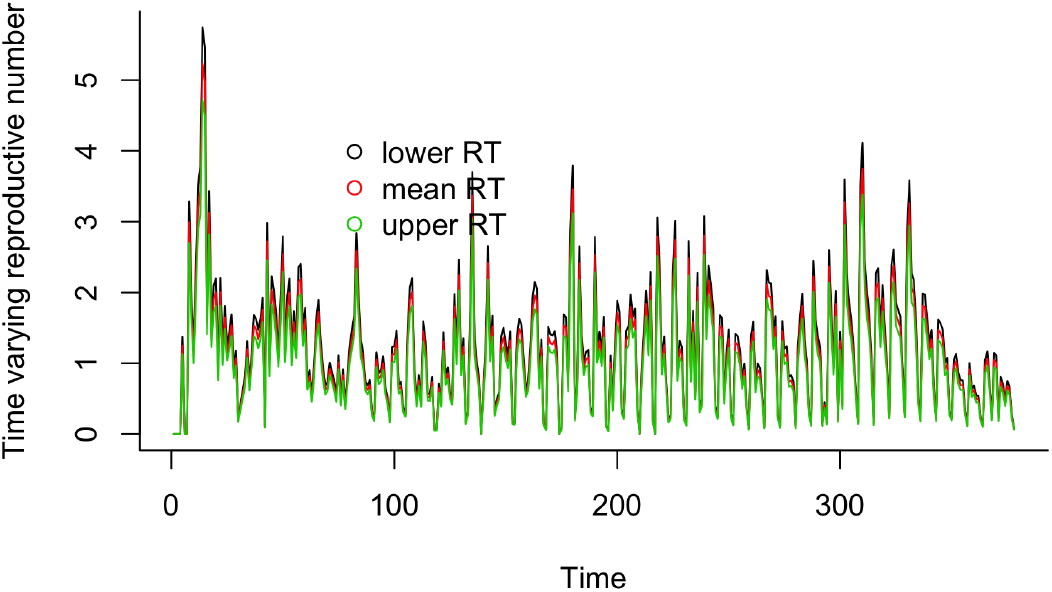
Time varying reproductive number in Mayotte with confidence interval of 95 per cent

**Figure 24:**
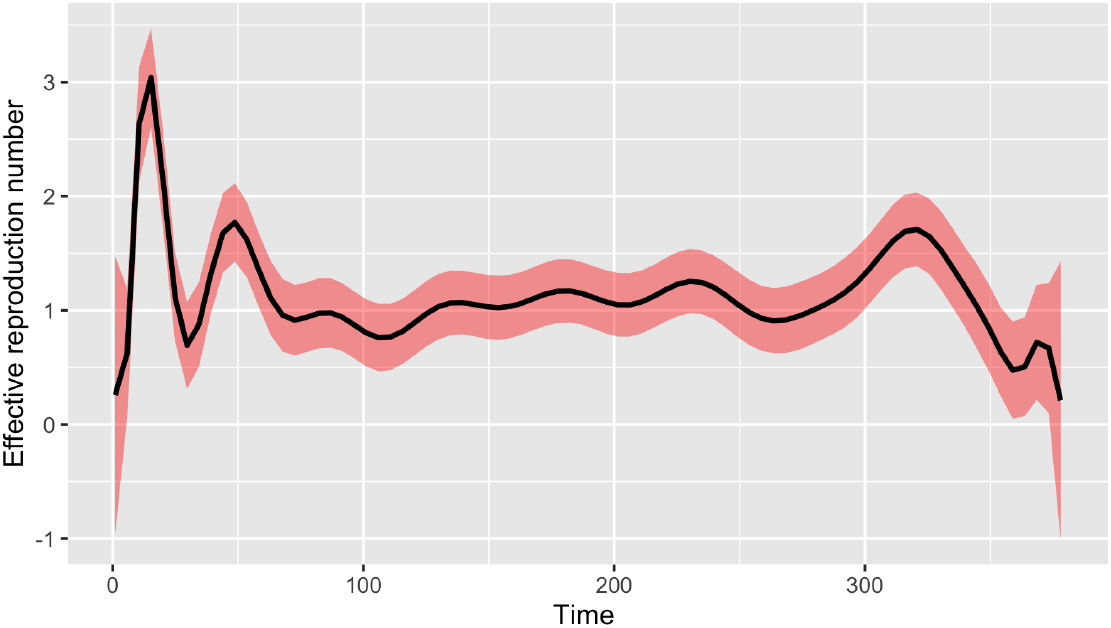
Evolution of the time varying reproductive number in Mayotte

**Table 3:** Estimate of *R*_0_ with a time horizon *T* = 35

| Method | Mean value |
| --- | --- |
| Maximum likelihood | $R_0 = 1.018325$ |
| Attack rate | $R_0 = 1.022491$ |
| Exponential growth rate | $R_0 = 1.056758$ |

We can see that at the start of the epidemic the basic reproductive number is around 3, which is consistent with the litterature. In the following picture, we make the curve smooth using estimates values.

Estimating of the number of reproduction number by the maximum likelihood, attack rate and exponential growth rate is given in the following table.

## 5 Numerical approximation of the Covid-19 epidemic curve in Mayotte

We set up here the numerical simulations by incorporating the above statistical estimates. The model equations are an age structured contact model. Such model allow the heterogeneous structure of the population since there is variation by age in the contact. The inclusion of the age and the contact improves the realism of a model,[21, 1]. Consider a population aggregated by age into *n* groups labelled by *i* = 1, 2, …, *n*. The population within age group *i* is partitioned into Susceptibles *S*_*i*_, Exposed *E*_*i*_, infected but not yet infectious, in a latent period; Infected and Infectious *I*_*i*_ being reported or unreported; Recovered *R*_*i*_. Let *N*_*i*_ be the total population of the age group *i*. We have

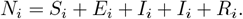

If we assume that there is no change in the age group over time, we have

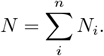

### Contact matrix

Denote by *C*_*i,j*_ the number of contacts of a susceptible individual in age group *i* with a susceptible individual in age group *j*;

#### Definition 4

*The matrix*(*C*_*i,j*_)_1*≤i,j≤n*_ *is called the contact matrix. It represents the average number of daily contacts of an individual of class i with an individual of class j*.

Clearly, the total number of contacts between group *i* to group *j* must be equal to the total number of contacts between group *j* to group *i*. Thus for closed population, we have the following reciprocity relation

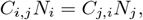

where *N*_*i*_ is the population in group *i*. By dividing the places of contact we obtain the following model of social contact

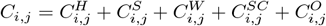

where 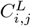 is the number of contacts of a susceptible individual in age group *i* with a susceptible individual in age group *j* in a place *L* (Home, School, Work,Shopping Center, Other).

A database of contact matrix from eight European countries was the subject of a study projected in many other countries, see [20] for more details.

Unfortunately Mayotte island is not referenced in this study which provides the contact matrix for several countries. A natural idea is to find an approximate contact matrix of a country with a similar population density and age pyramid. We choose that of Rwanda because the 2019 population density in Rwanda is 525 people per *Km*^2^ and that of Mayotte is 682 people per *Km*^2^. The following picture show the population pyramid of Rwanda and Mayotte as weel as the contact matrix of Rwanda.

**Figure 25:**
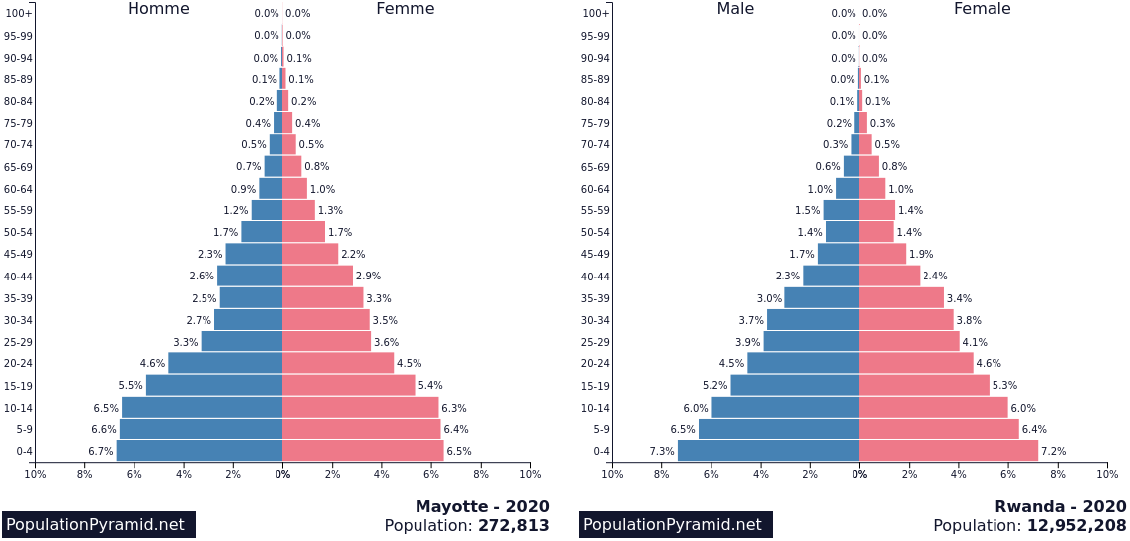
Age pyramid of Mayotte (left) and Rwanda (right) in 2020. Source (https://www.populationpyramid.net/rwanda/2020/)

**Figure 26:**
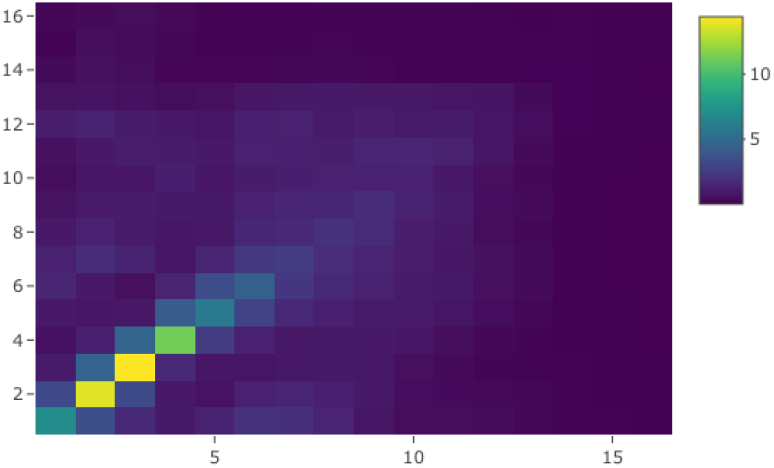
Contact matrix of Rwanda

### 5.1 Simulation based on the age-structured contact SEIRD model

The differential equations that govern the trajectories of our model are formulated as follow:

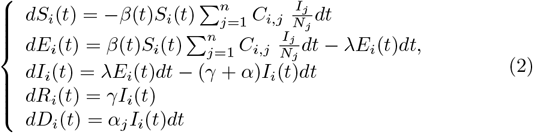

with

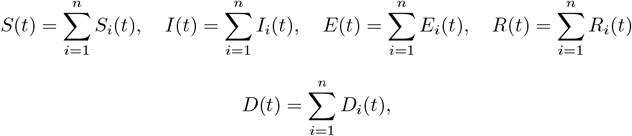

*β* is the rate of infection on contact assumed intrinsic to the pathogen. We consider closed populations because we assume that deaths are balanced by births and immigration. The induced mortality rate *α*_*j*_ depend on the age in this model. For simulation purpose we choose *n* = 16 to correspond to the 16 age group into which the contact matrix data is partitioned. Then the 5 ∗*n* = 80 ordinary differential equations are then numerically integrated using *R*-software. The following table lists the parameters of the model we’ll use for numerical simulation. These estimates are often wide due to wide variations from one infection to another. We also give the initial values of the model.

**Table 4:**
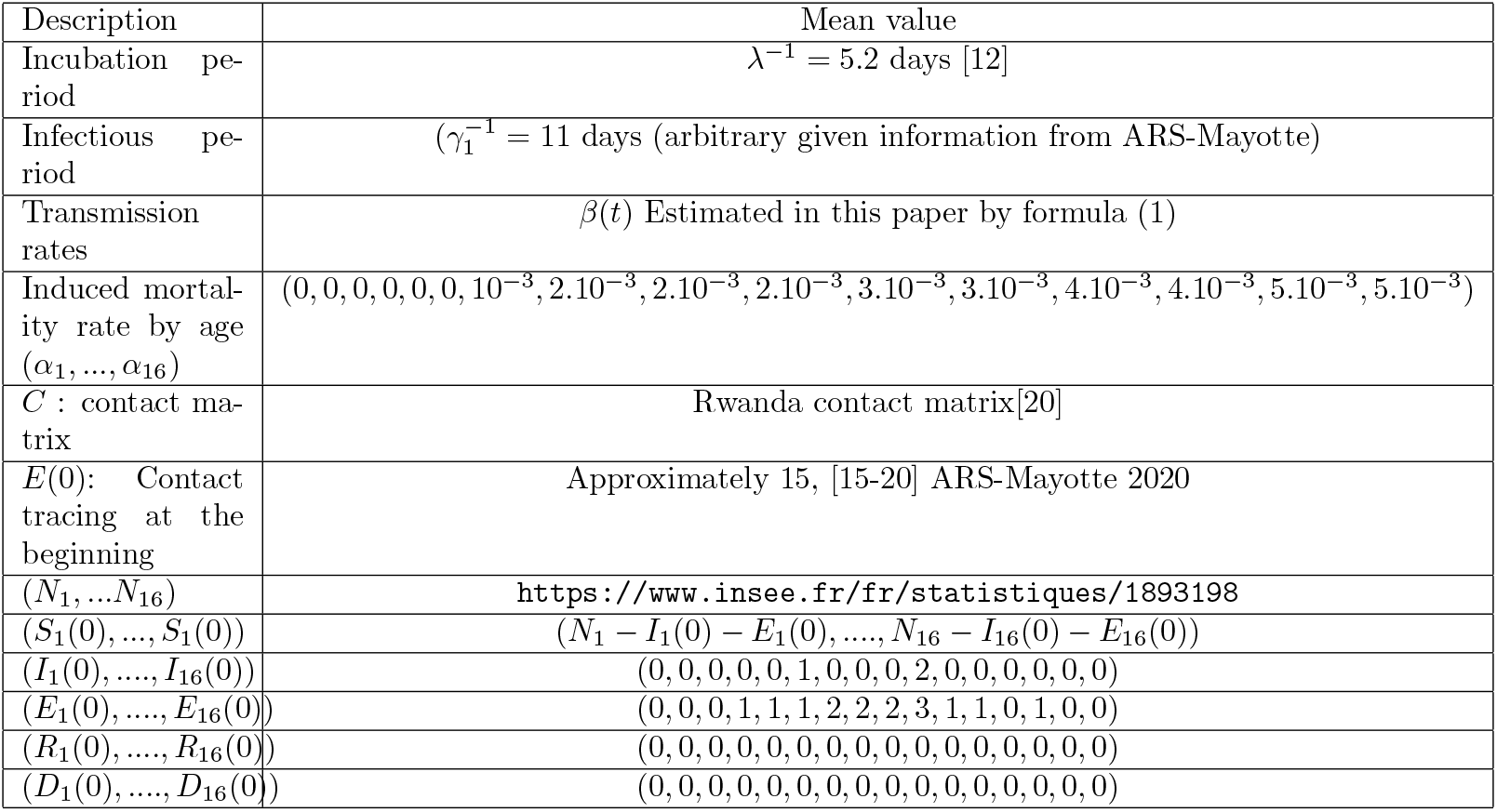
List of the parameters and initial values of the model

The following pictures show by simulation the progress of the epidemic by age. By observing the curve of infected by age group, we observe that the epidemic peak is greater in the age group 1 to 4 corresponding to the age class [0 − 19], followed by age group 5 to 9 ([20 − 44]), then age group 10 to 12 ([45 − 59]) and then age group 13 to 16 ([60+]).In this last class the age group [60 − 64] is more affected than the class 65+. This shows that there is a sensitivity according to the age group. The age group [0 − 19] is more exposed which means that it is a population to be protected by various means such as vaccination or isolation.

**Figure 27:**
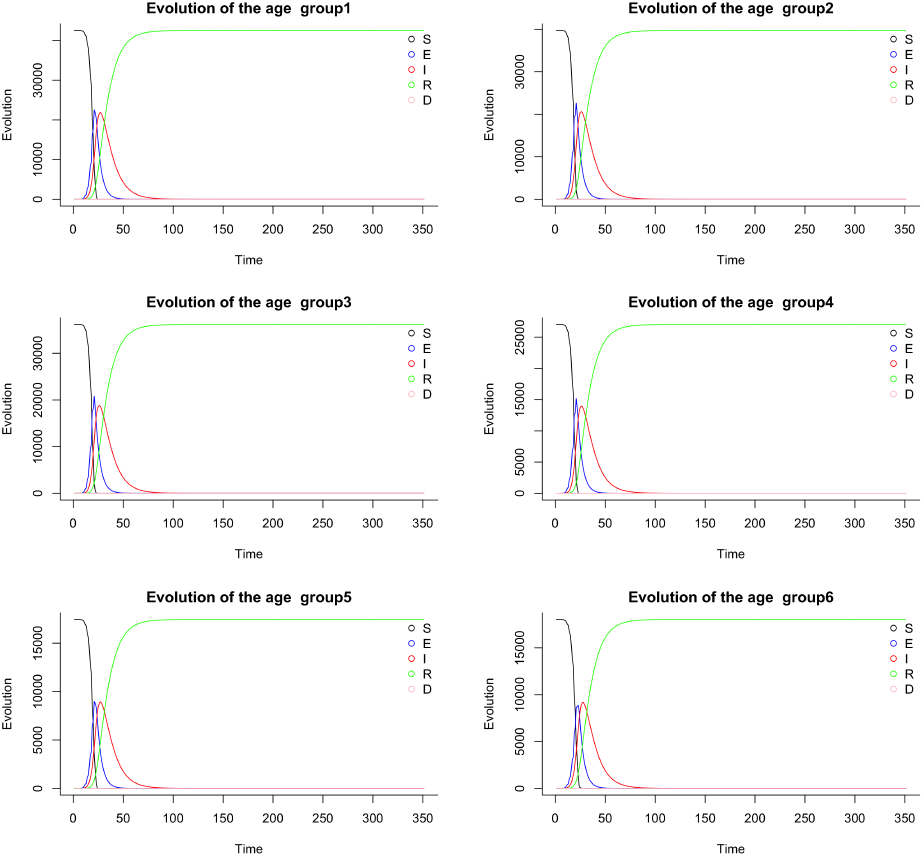
Simulating the age-structured contact model

**Figure 28:**
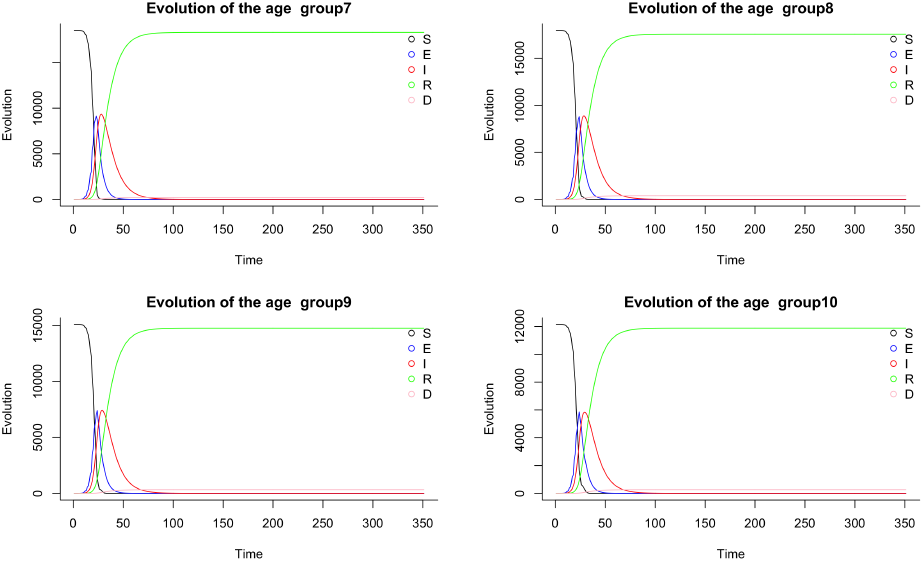
Simulating the age-structured contact model

**Figure 29:**
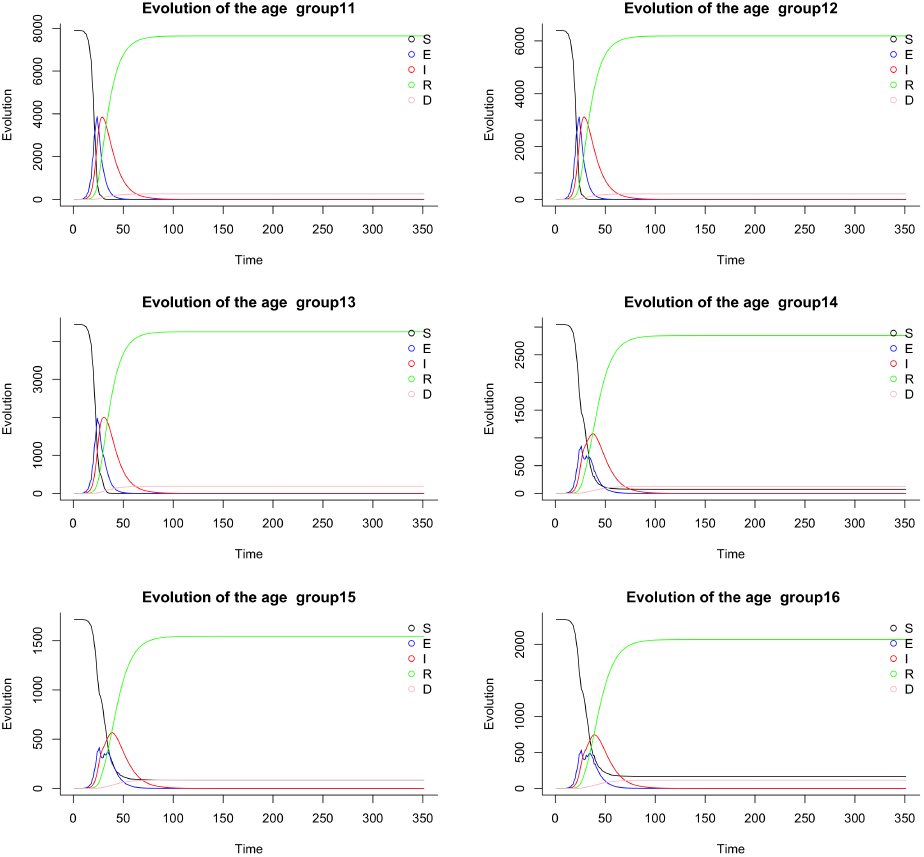
Simulating the age-structured contact model

## 6 Discussion and conclusion

In the above simulation, we did not integrate the vaccination because it was assumed that during the study interval of the epidemic curve, vaccination in Mayotte had no effect because it started in February 2021, see https://www.mayotte.ars.sante.fr. Given the current state of the epidemic in Mayotte, the above numerical simulation of the epidemic curve show that, new factors have contributed to modify the dynamics of the epidemic. It is important to note that the simulation is only theoretical and restricted by given conditions. A conclusion that can be drawn subject to assumptions is that mitigation and non-pharmacological interventions of the Covid-19 of any kind can possibly be considered in the model such as the factors of reinfection, immunity and the appearance of variants. Note also that some conditions can be dramatically different in reality for instance the infectious and the virus mortality rate by age. It should also be noted a bias in the approximate use of the contact matrix. In a forthcoming work, we will develop a model taking into account the possibilities of reinfection, vaccination, immunity over a longer period.

## Data Availability

All data produced are available online at https://www.data.gouv.fr/fr/pages/donnees-sante.

## 7 Supporting information

Additional supporting information of the original contributions presented in the study are included in the article/supplementary material, further inquiries can be directed to the corresponding author/s. Other data about the population used in this study are openly available from the French INSEE web site. Note that data relating to covid-19 can be downloaded from https://www.data.gouv.fr/fr/pages/donnees-sante/. As concerned the data on contact matrix, they are available in the supplementary material of [20]. We provide in the supporting information our complete R codes in an R-markdown format.

---

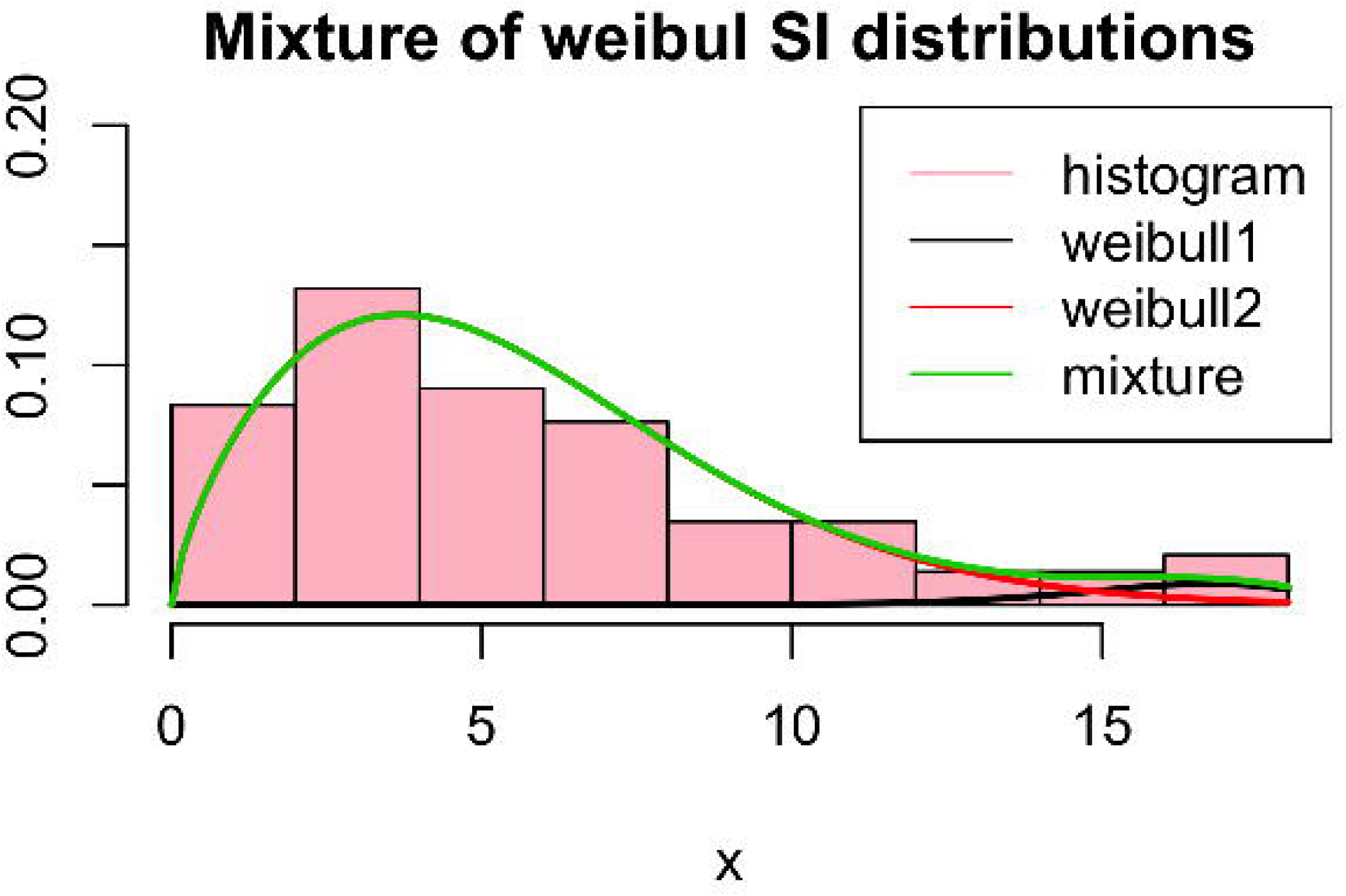

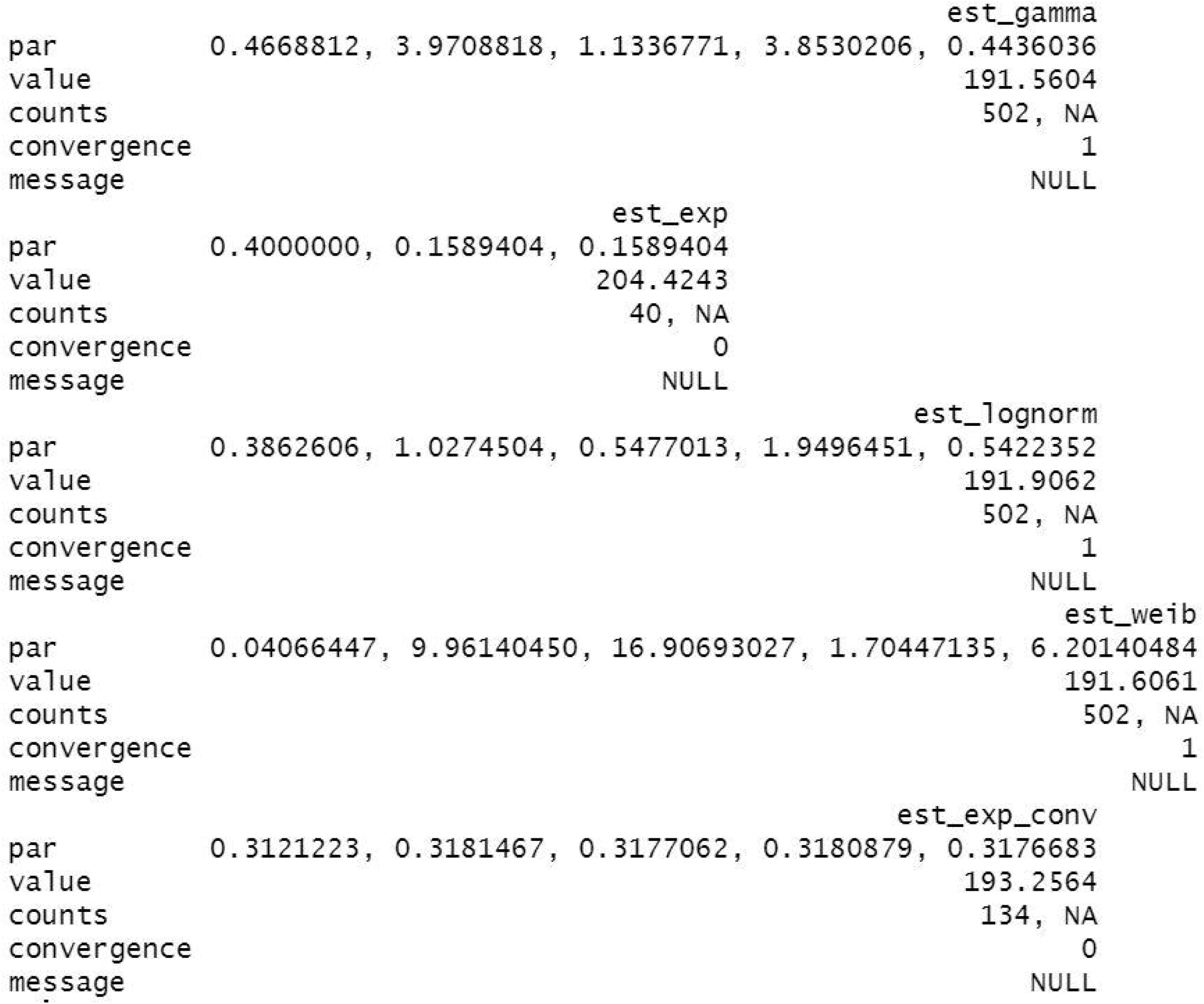

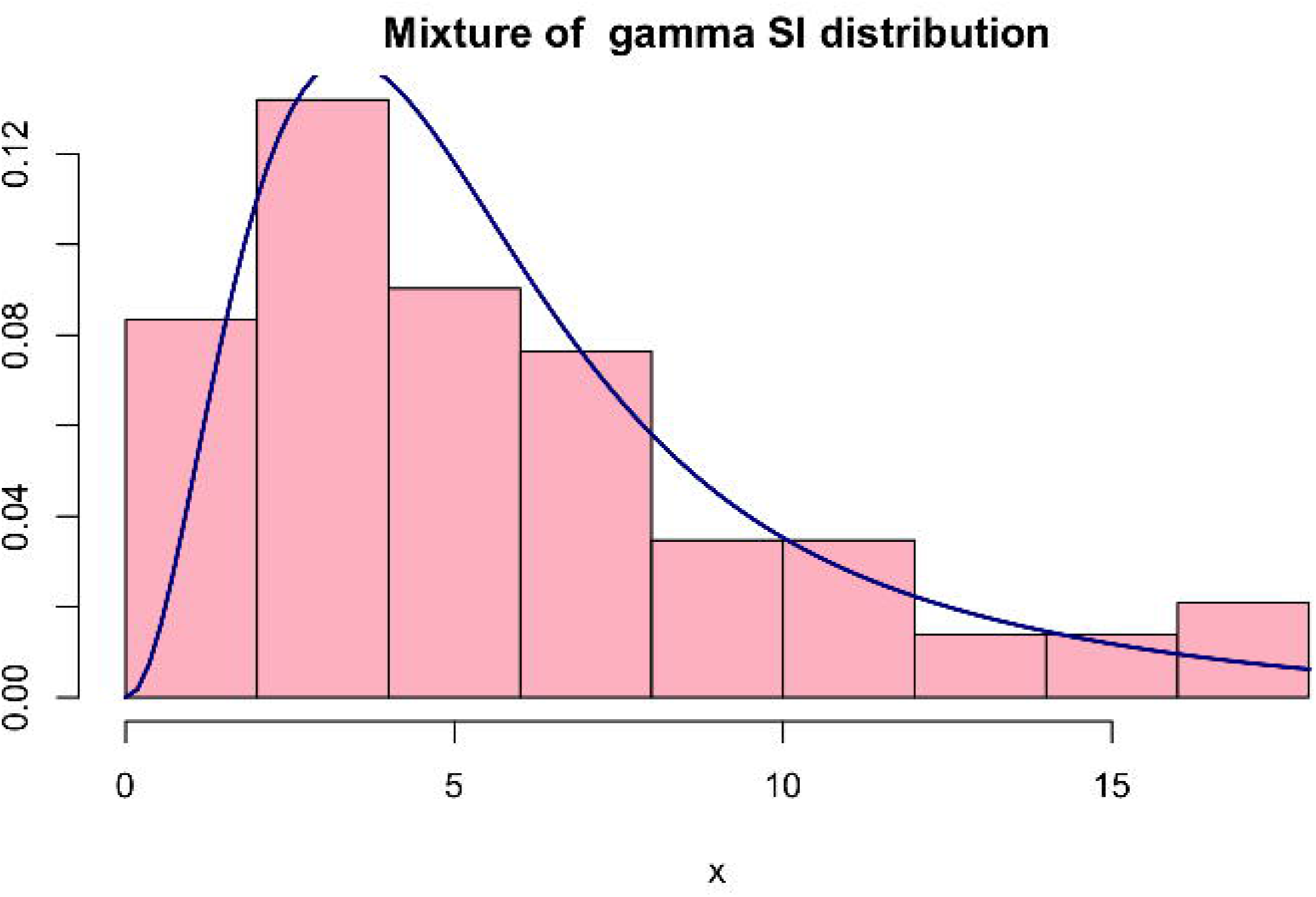

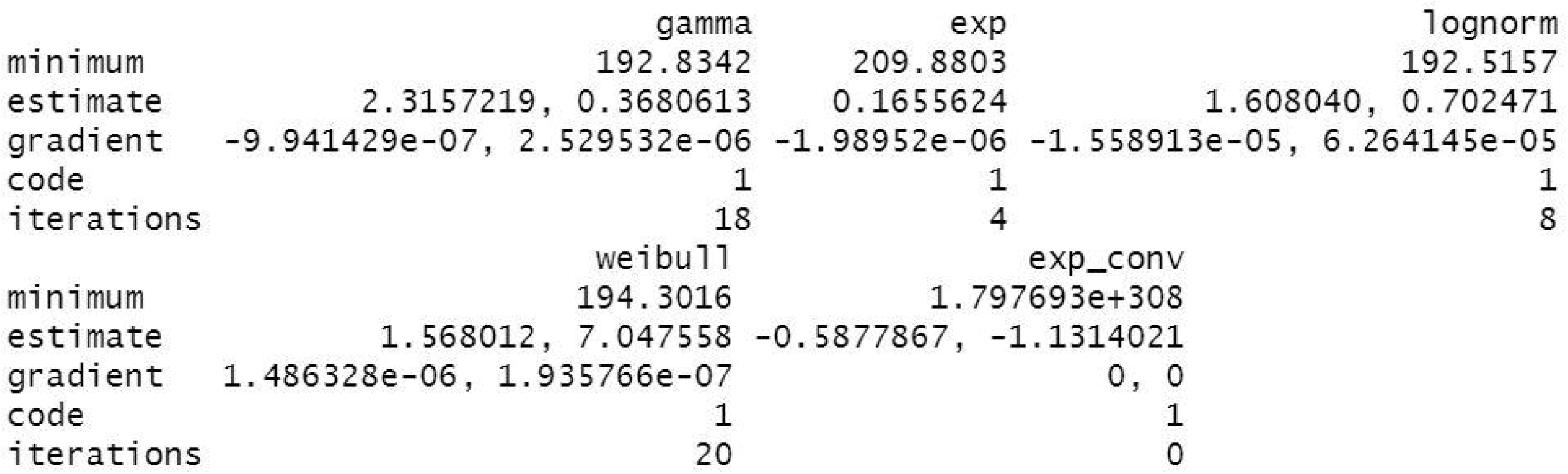

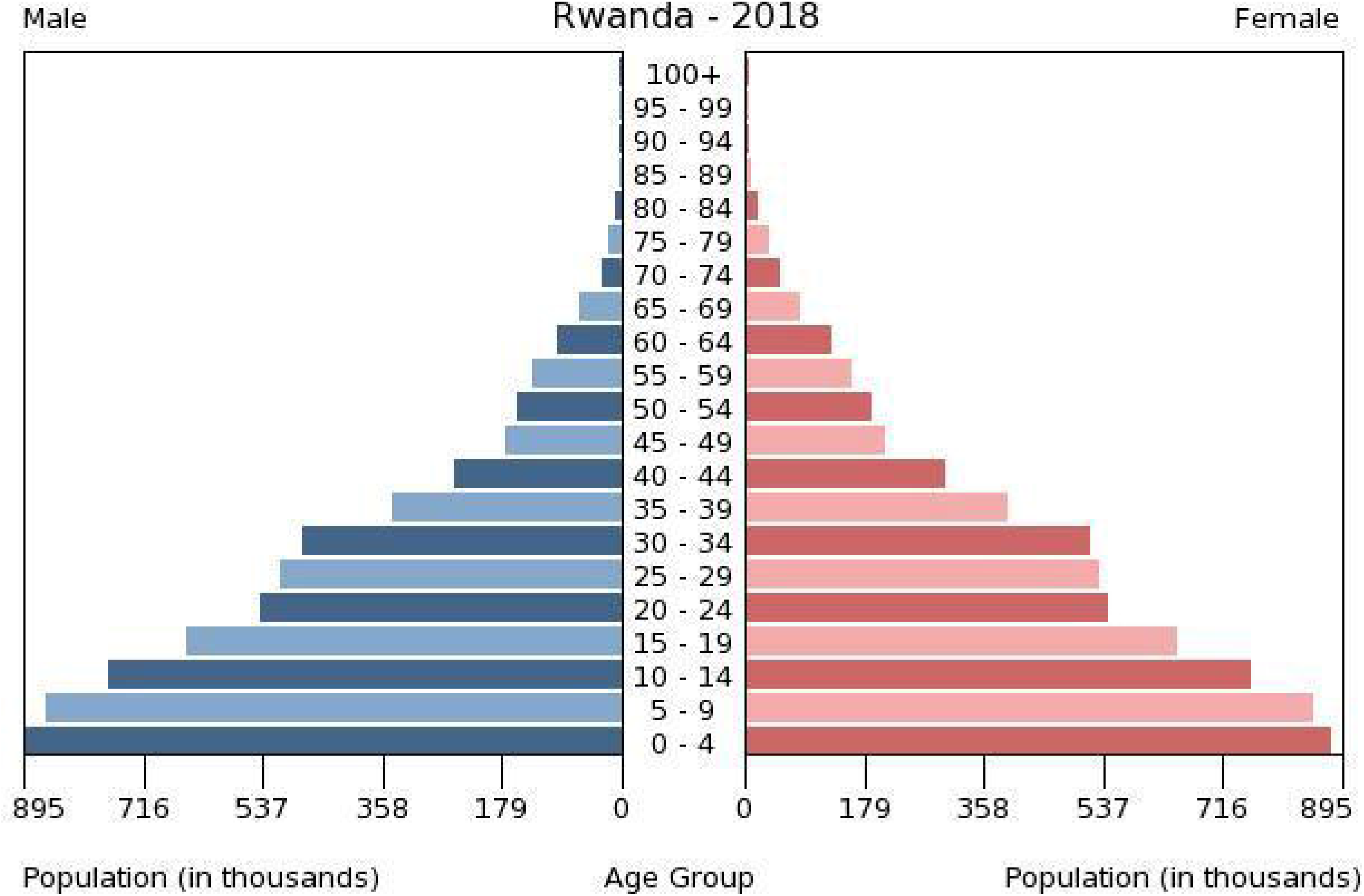

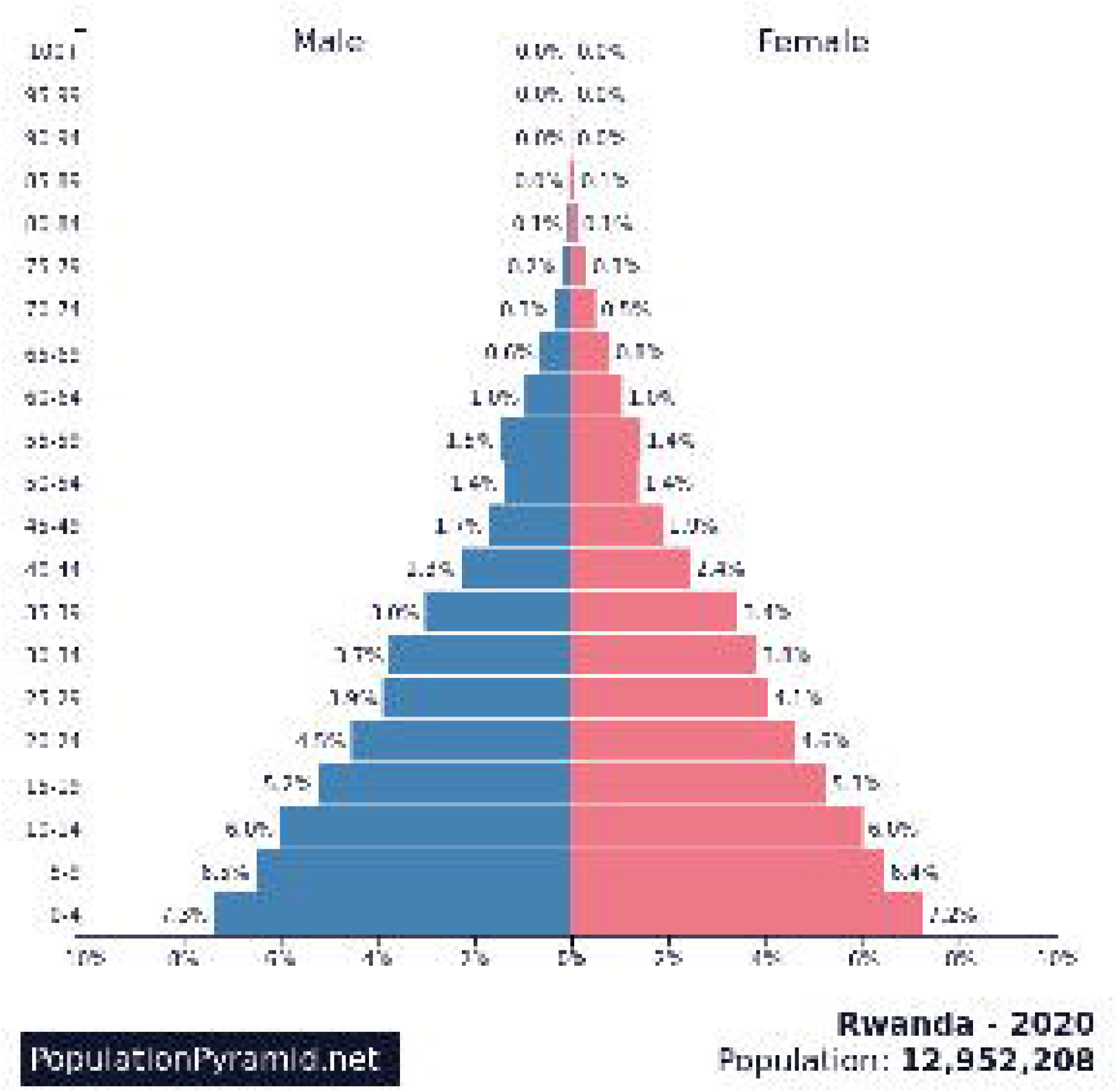

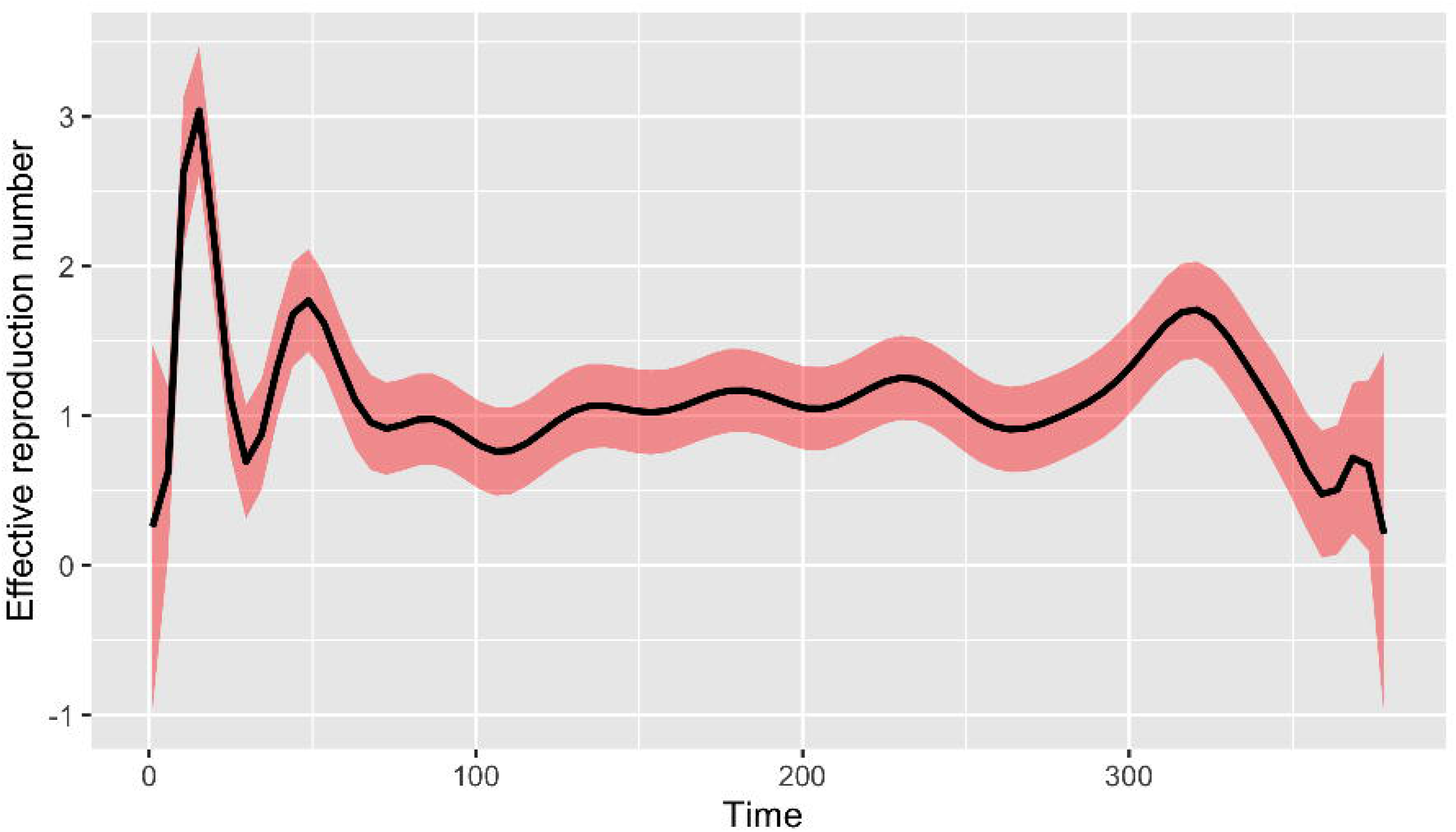

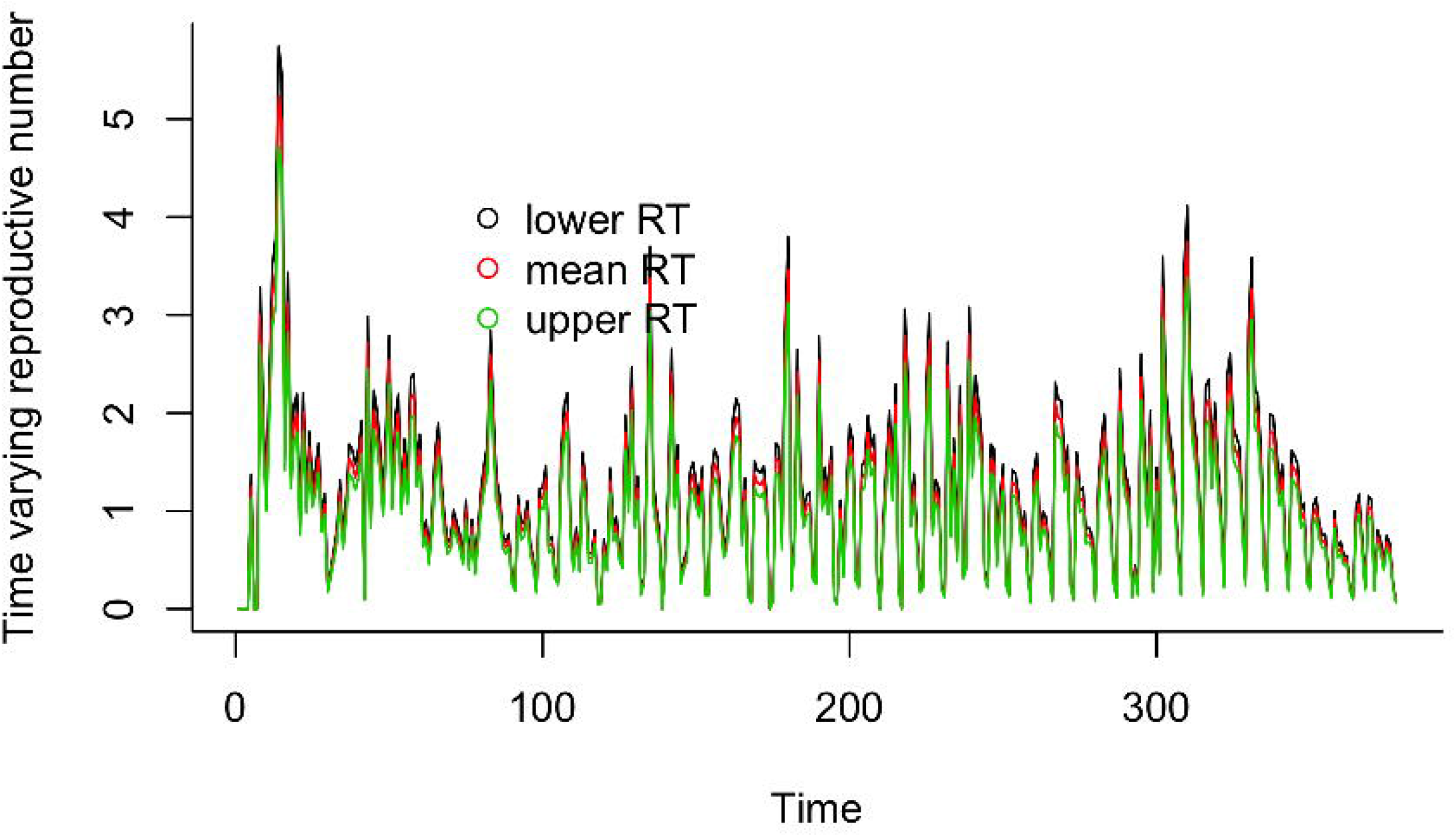

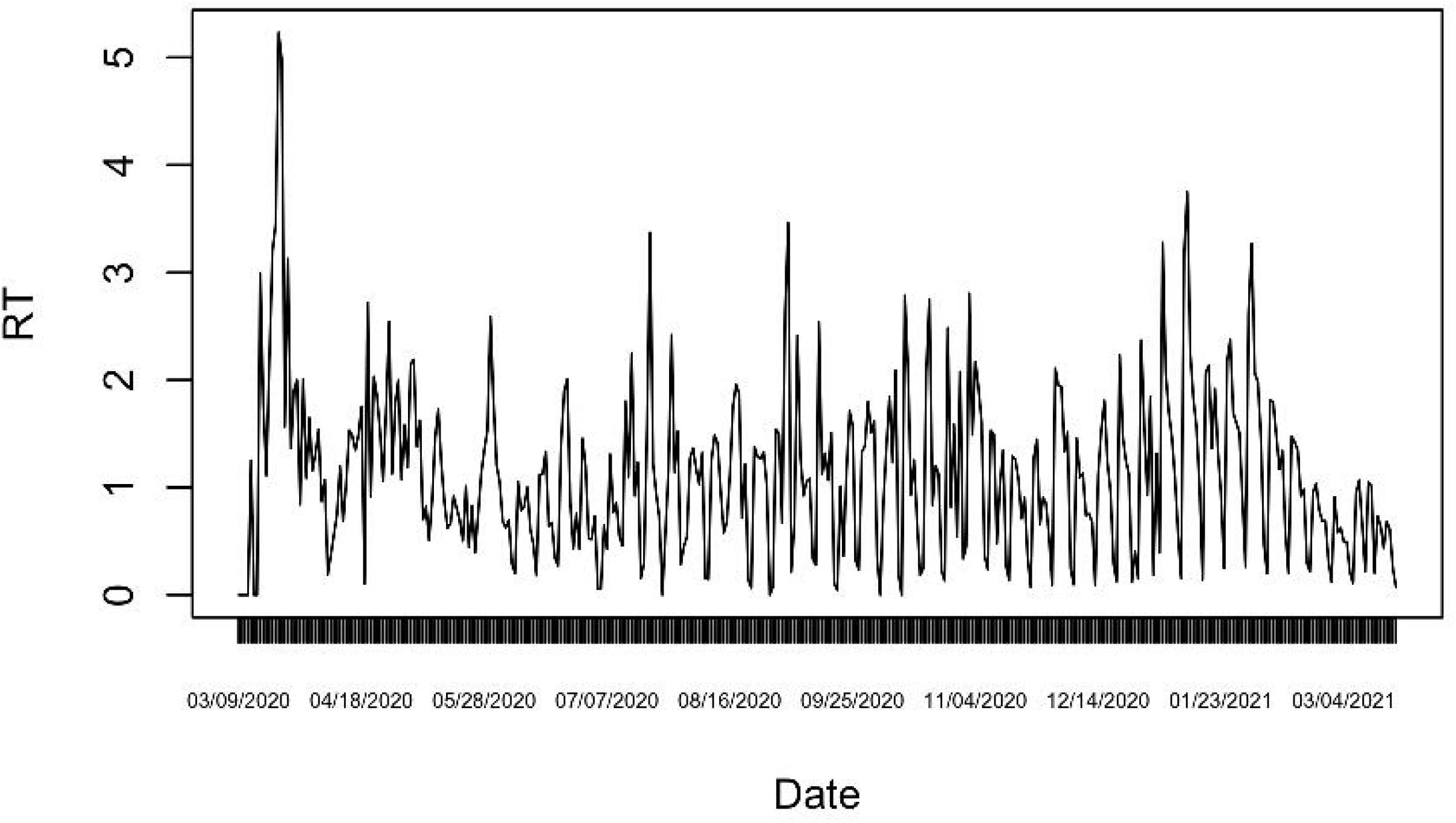

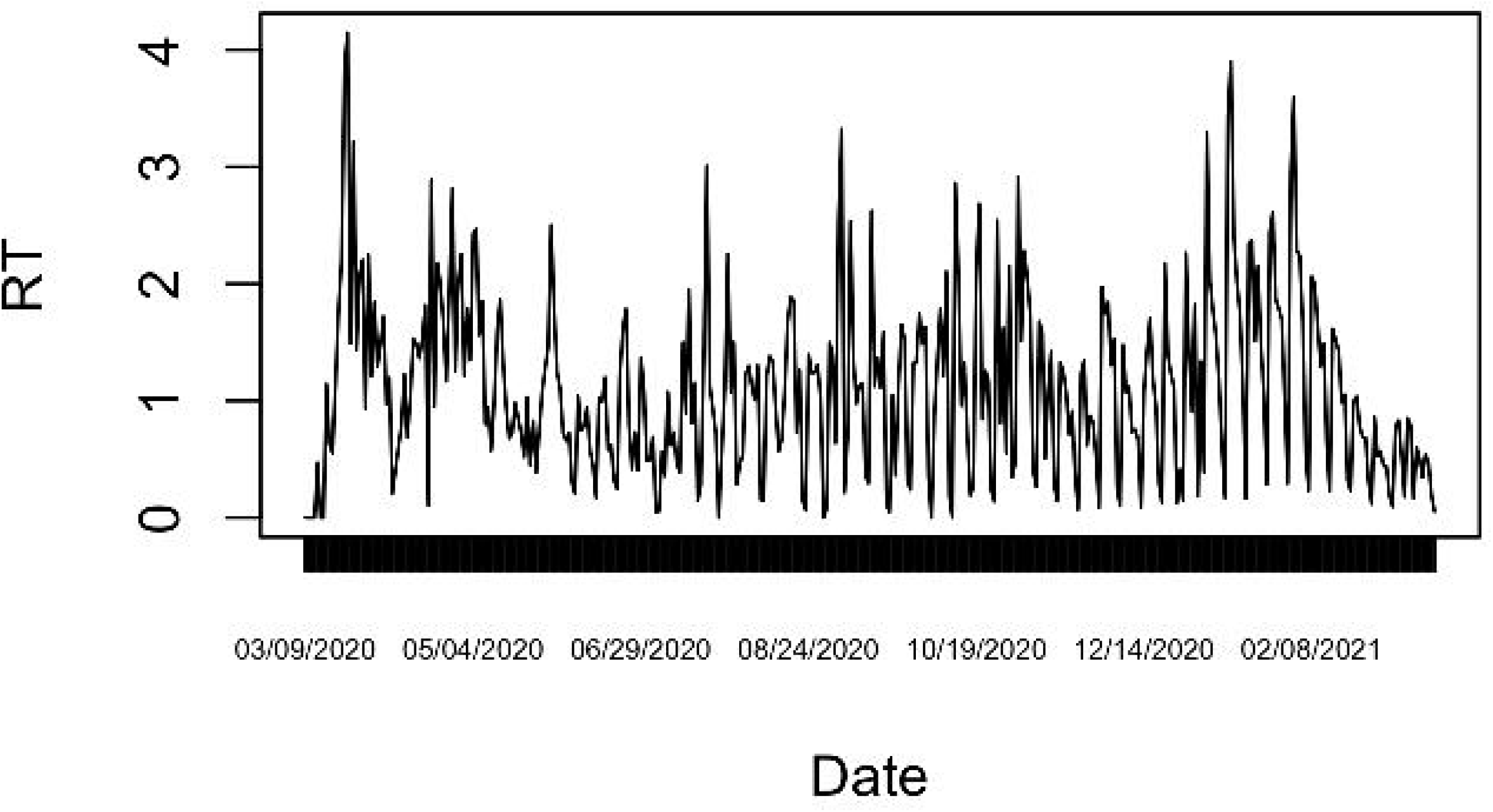

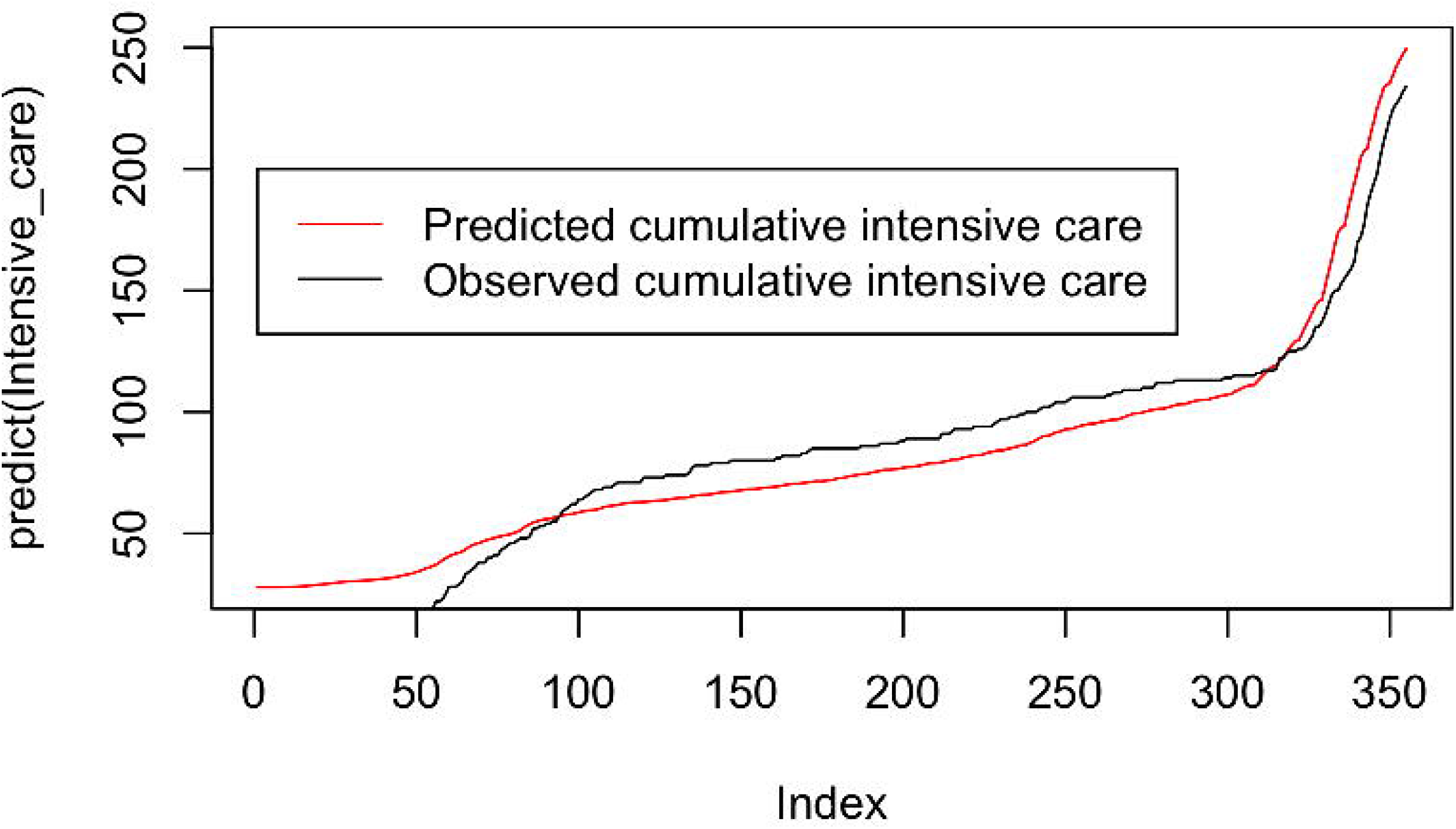

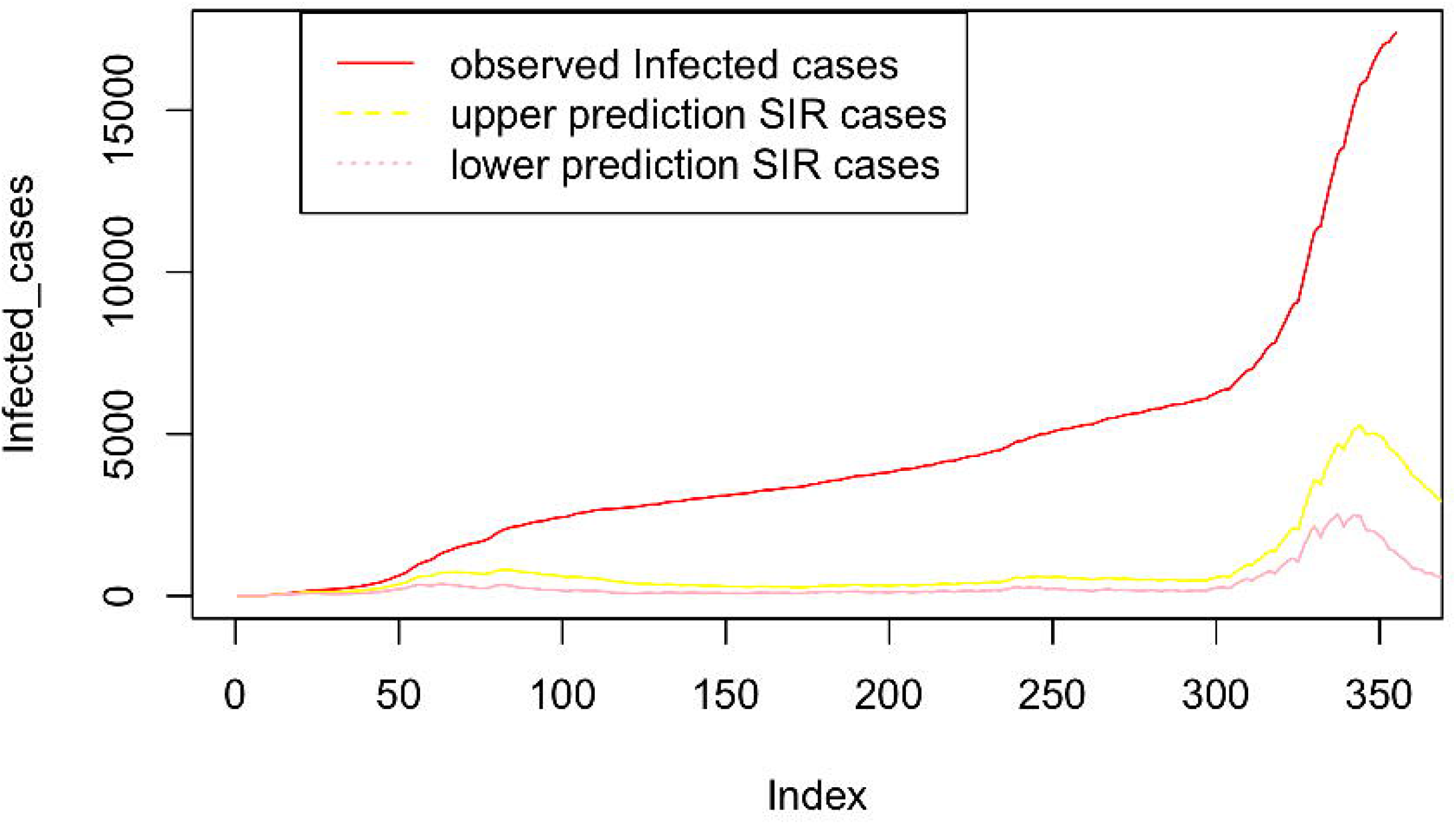

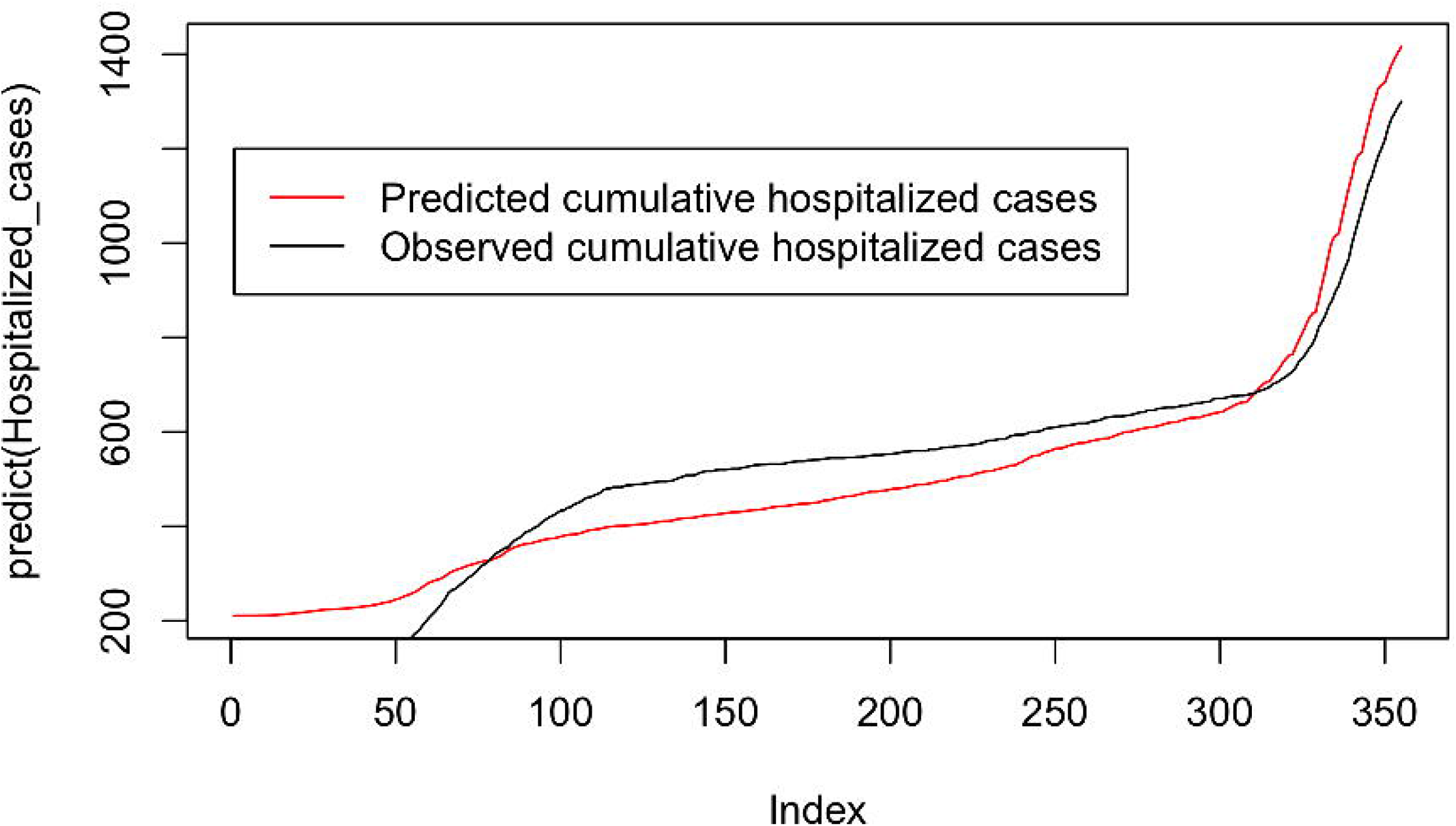

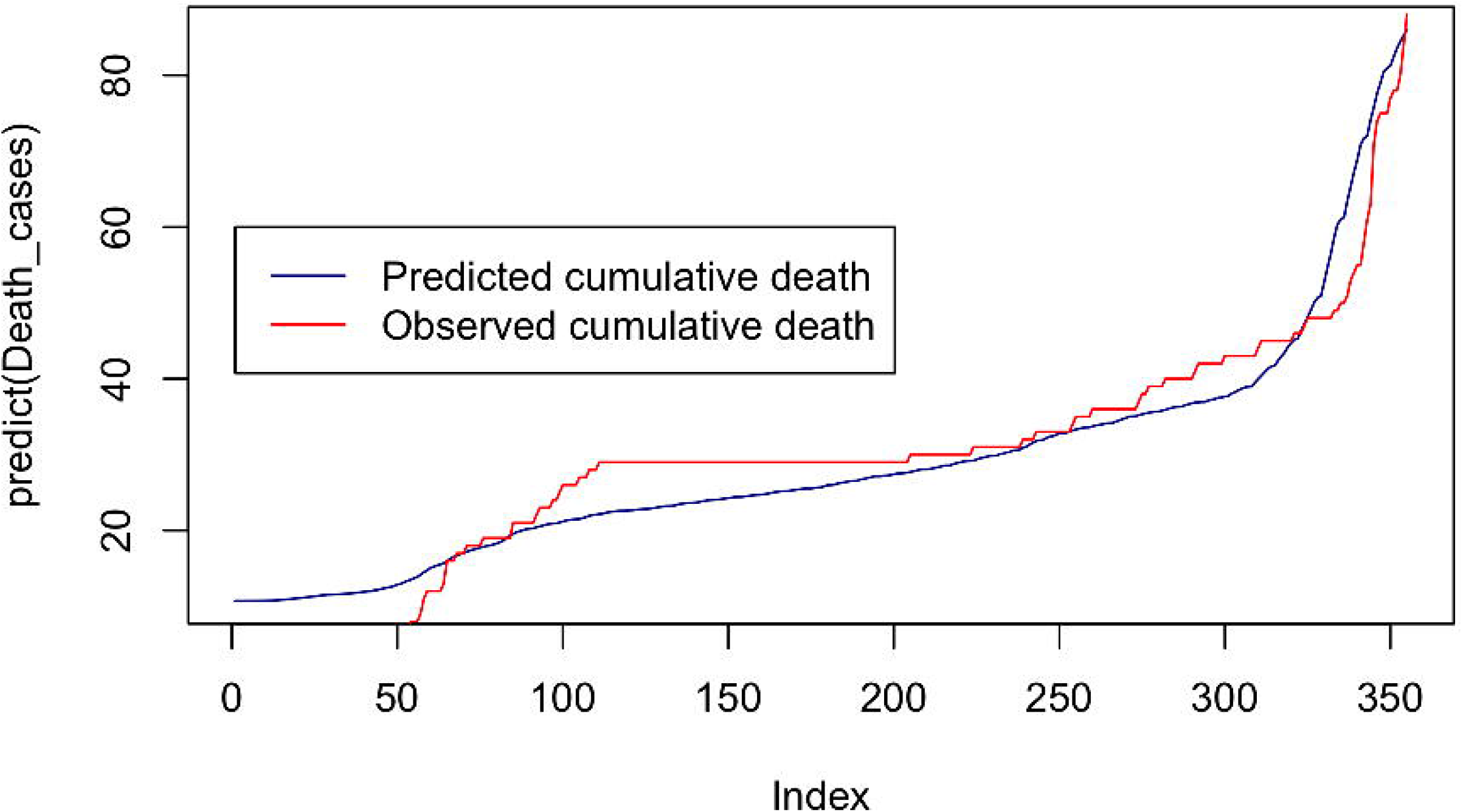

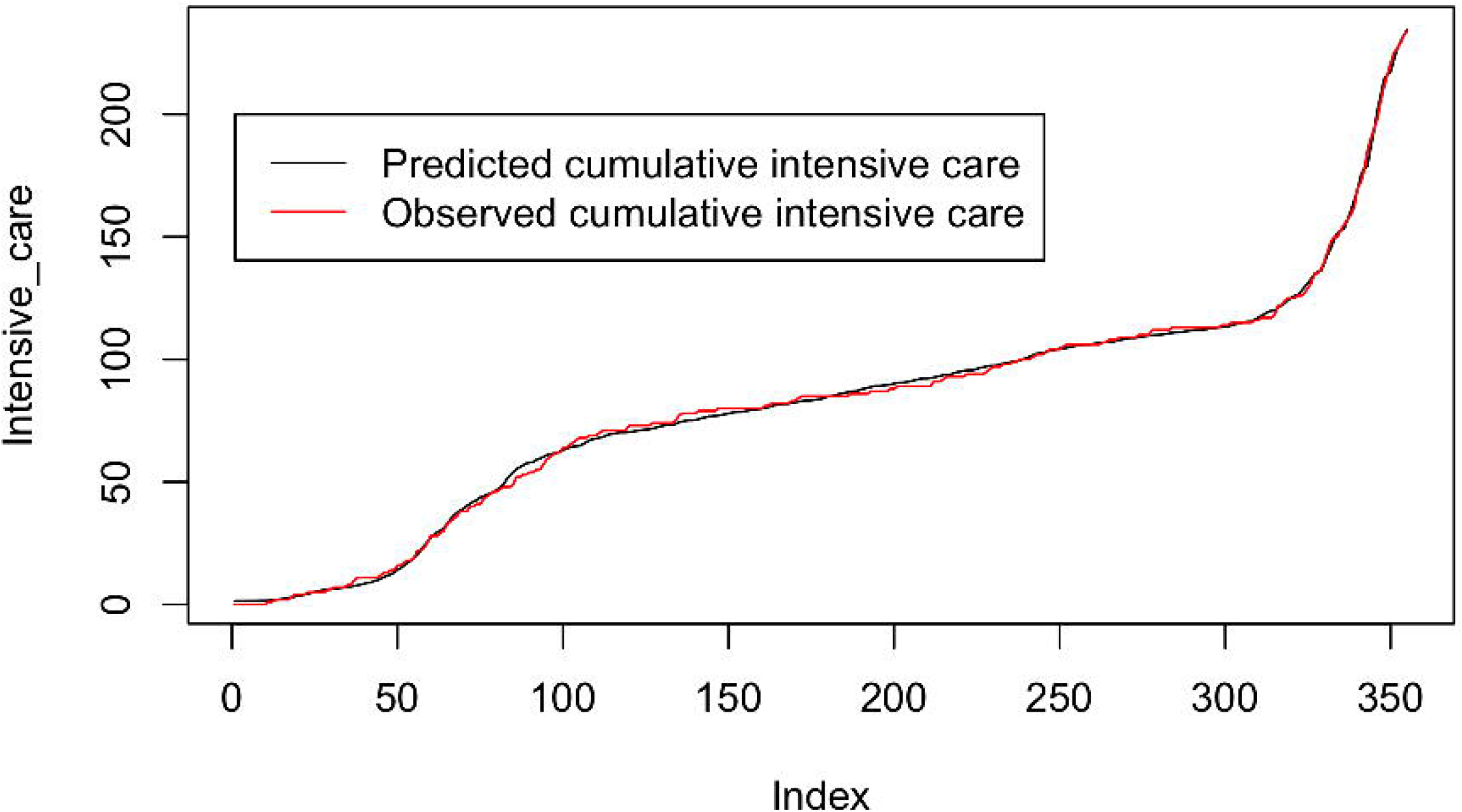

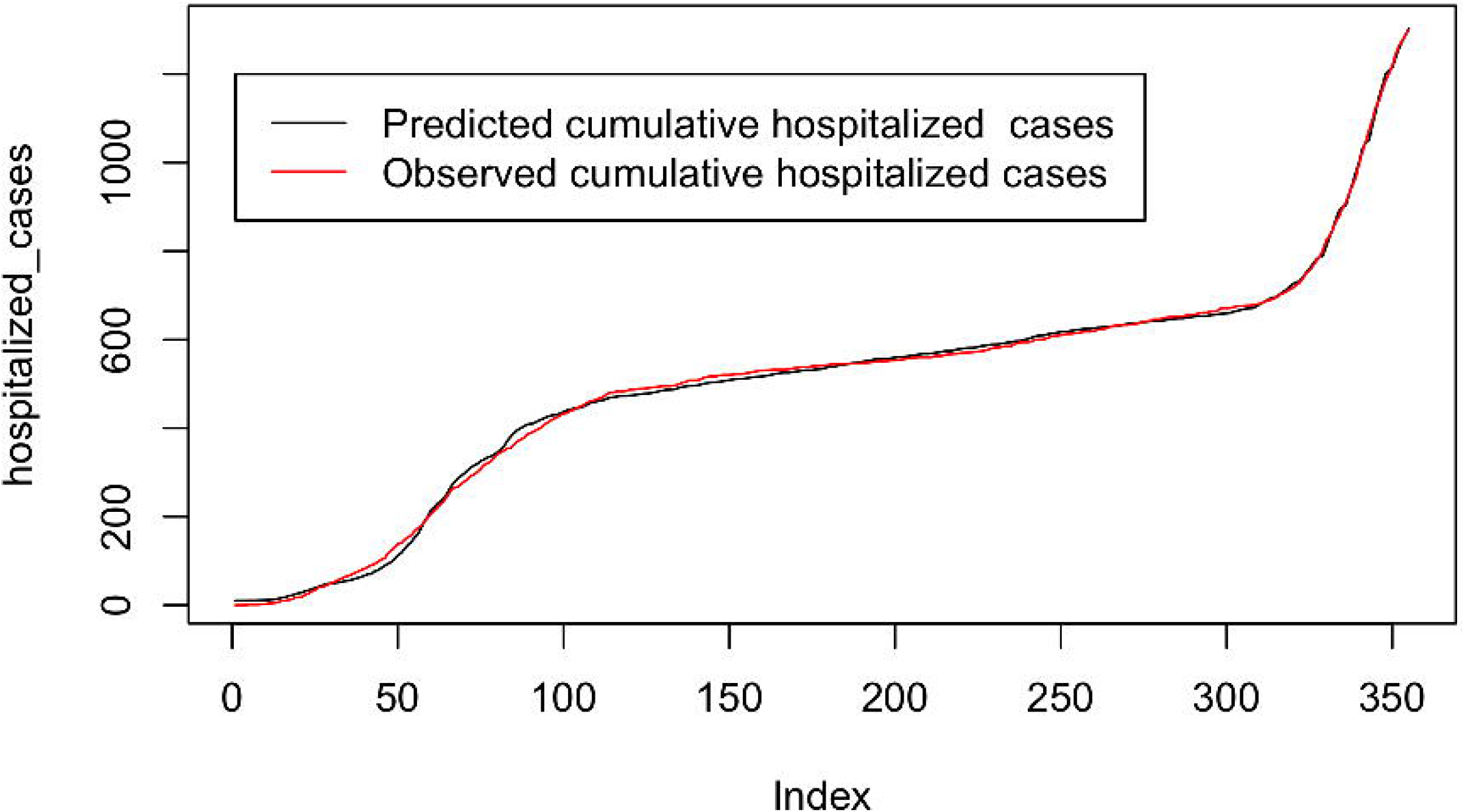

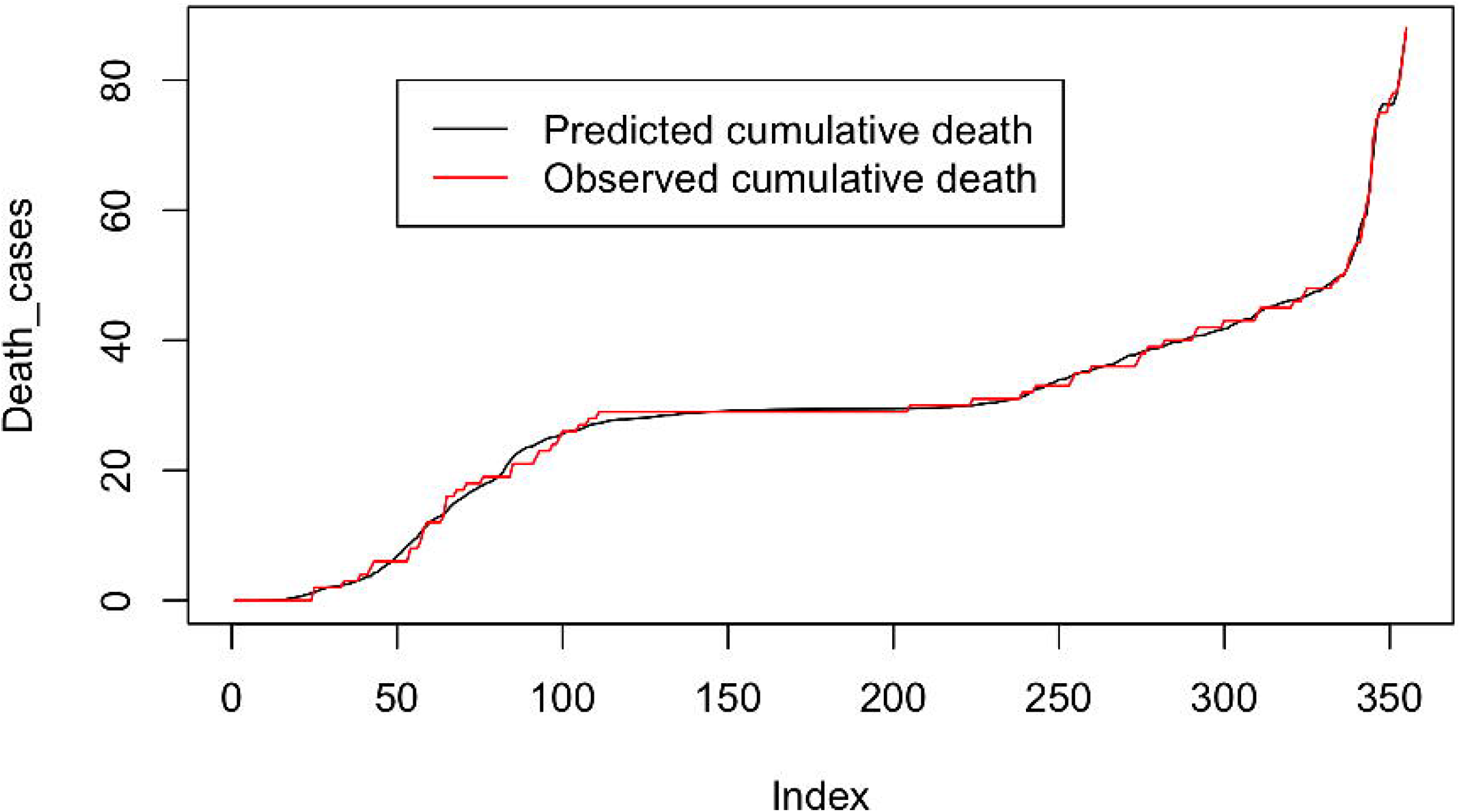

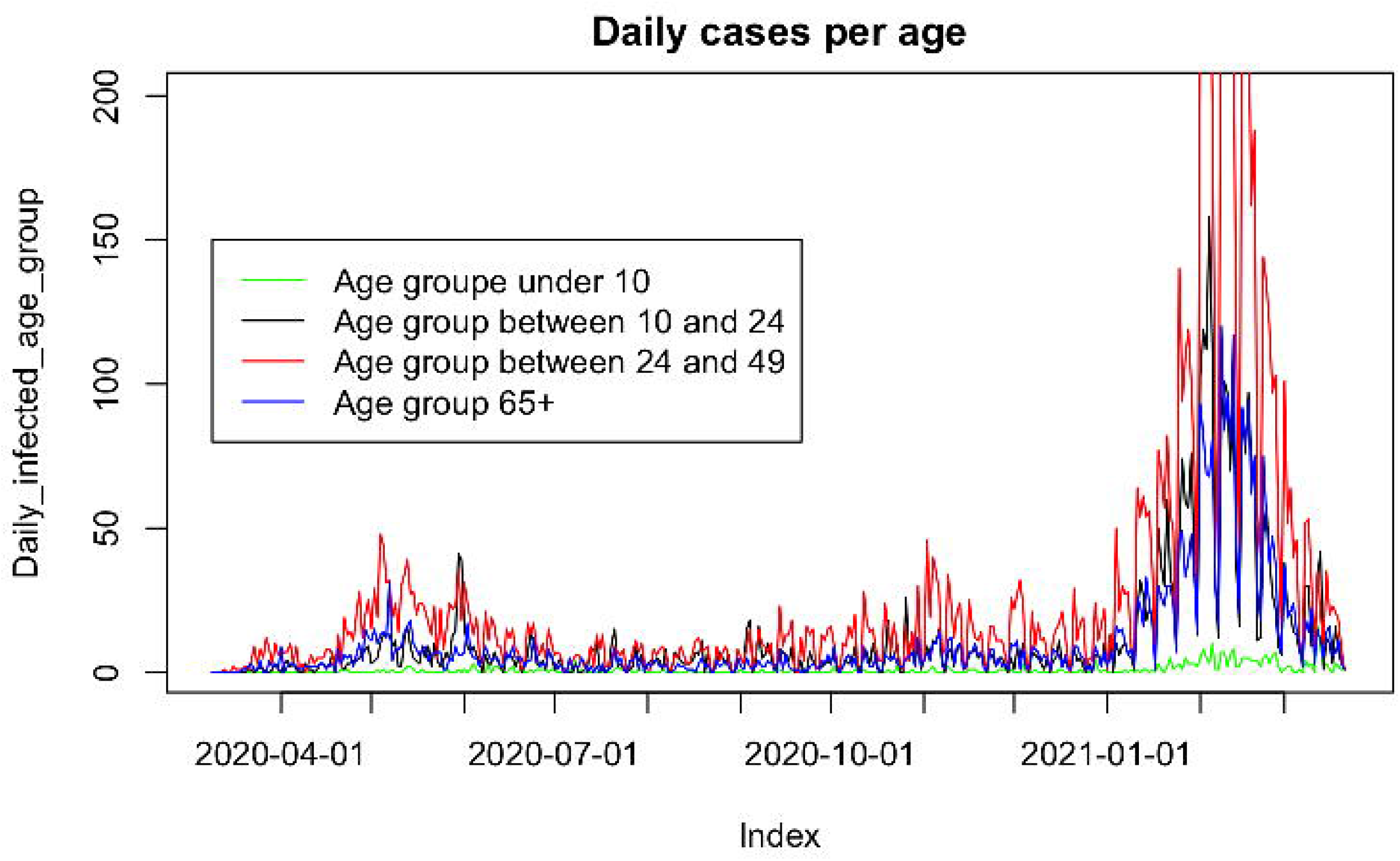

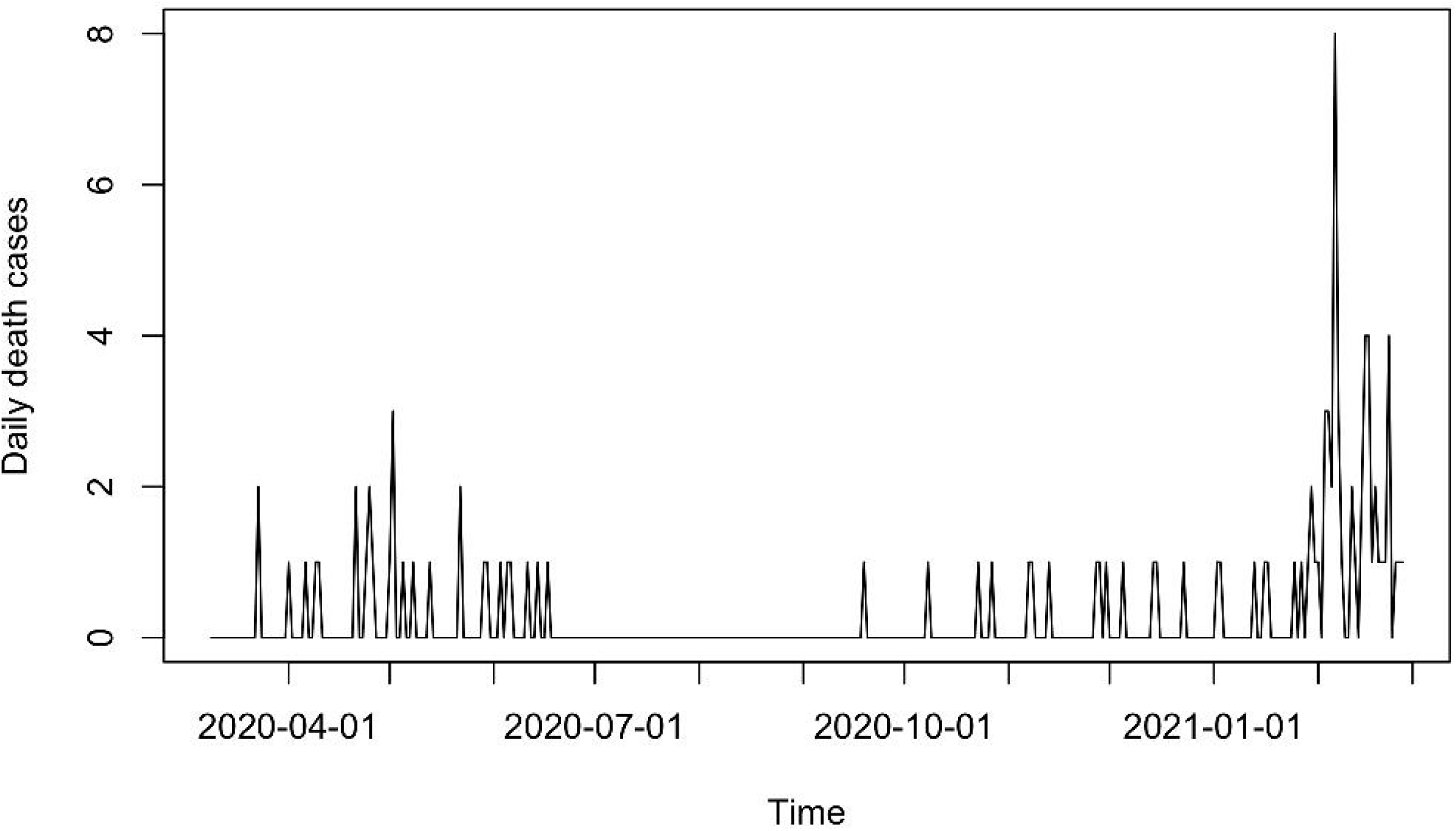

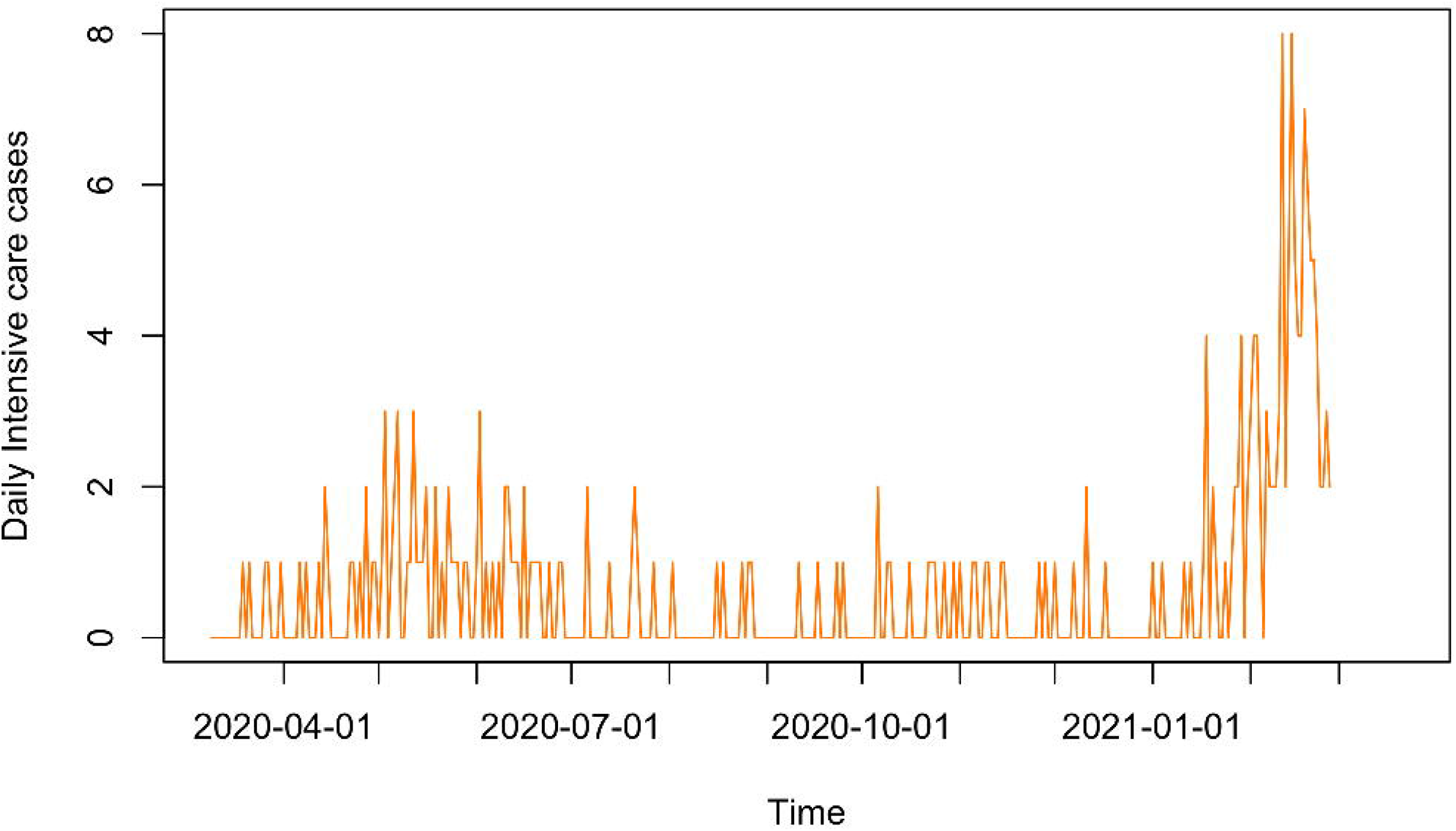

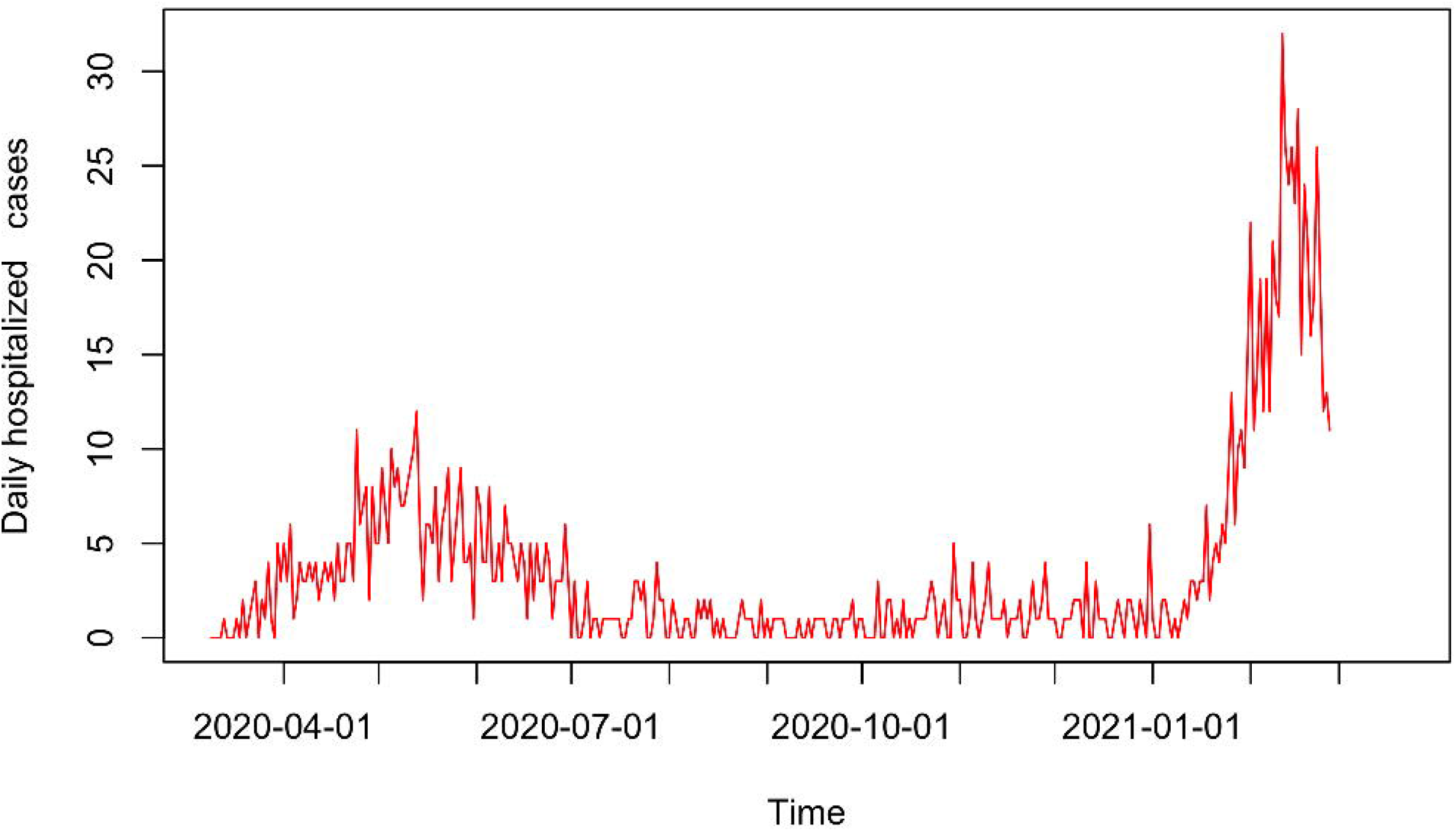

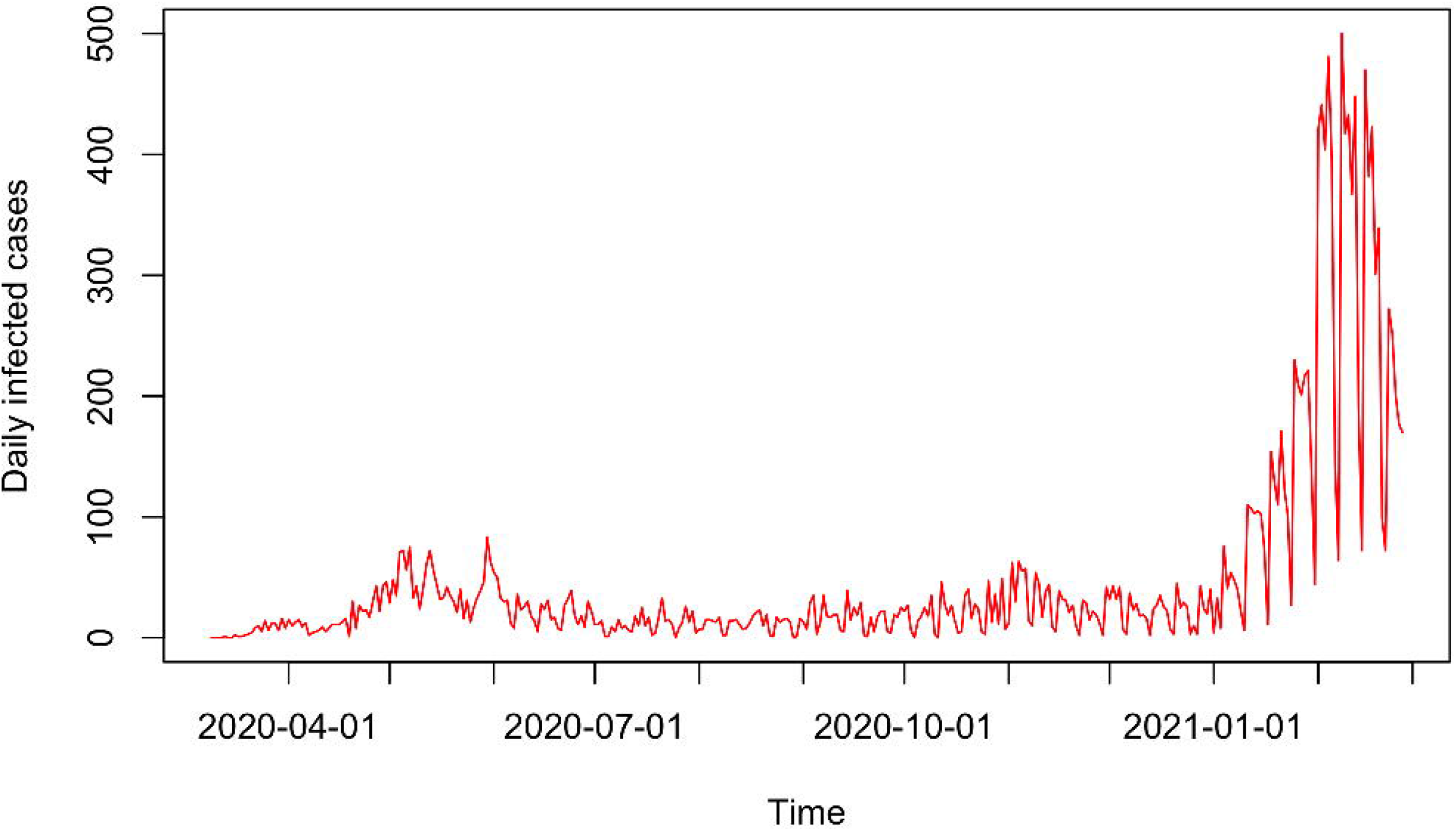

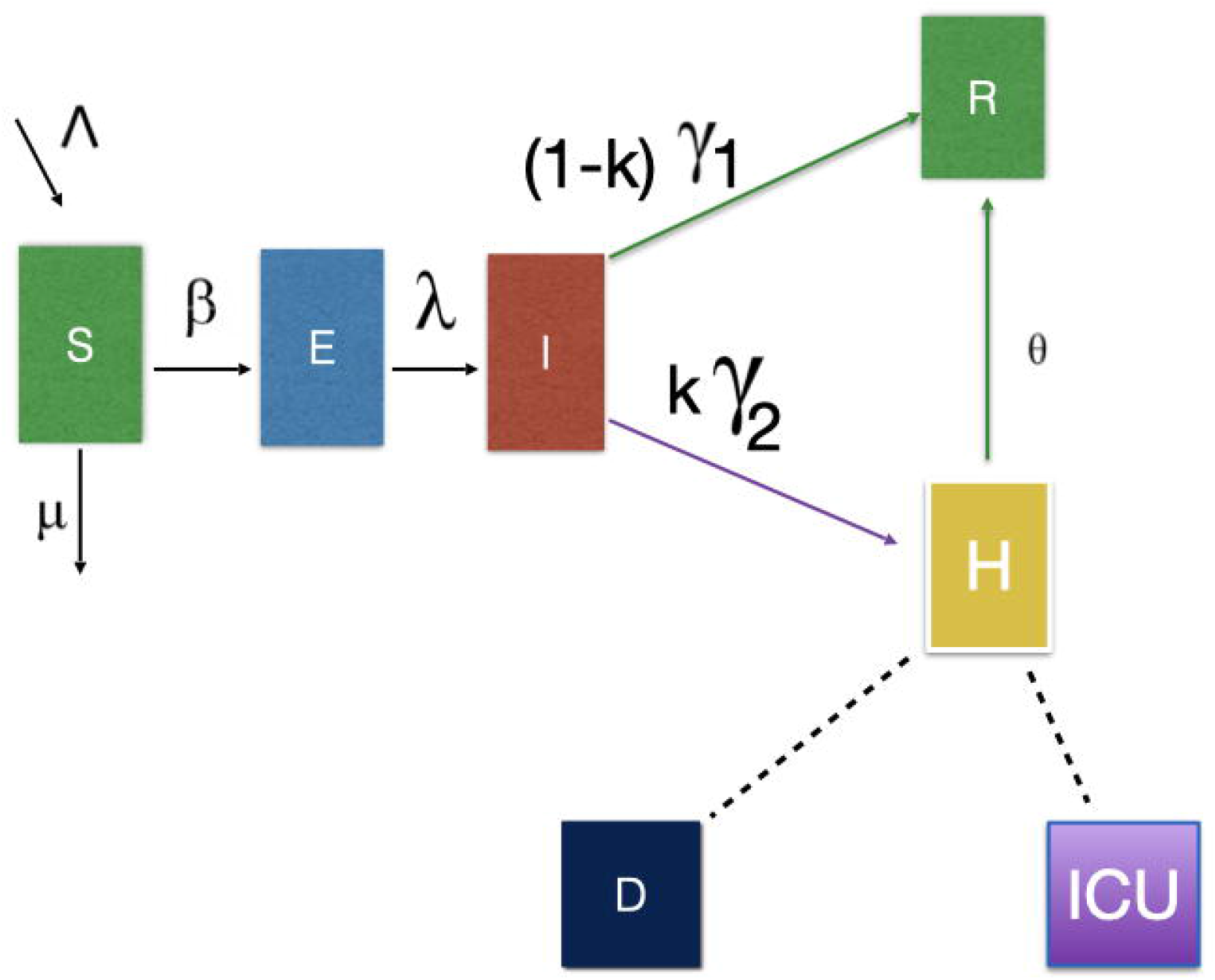

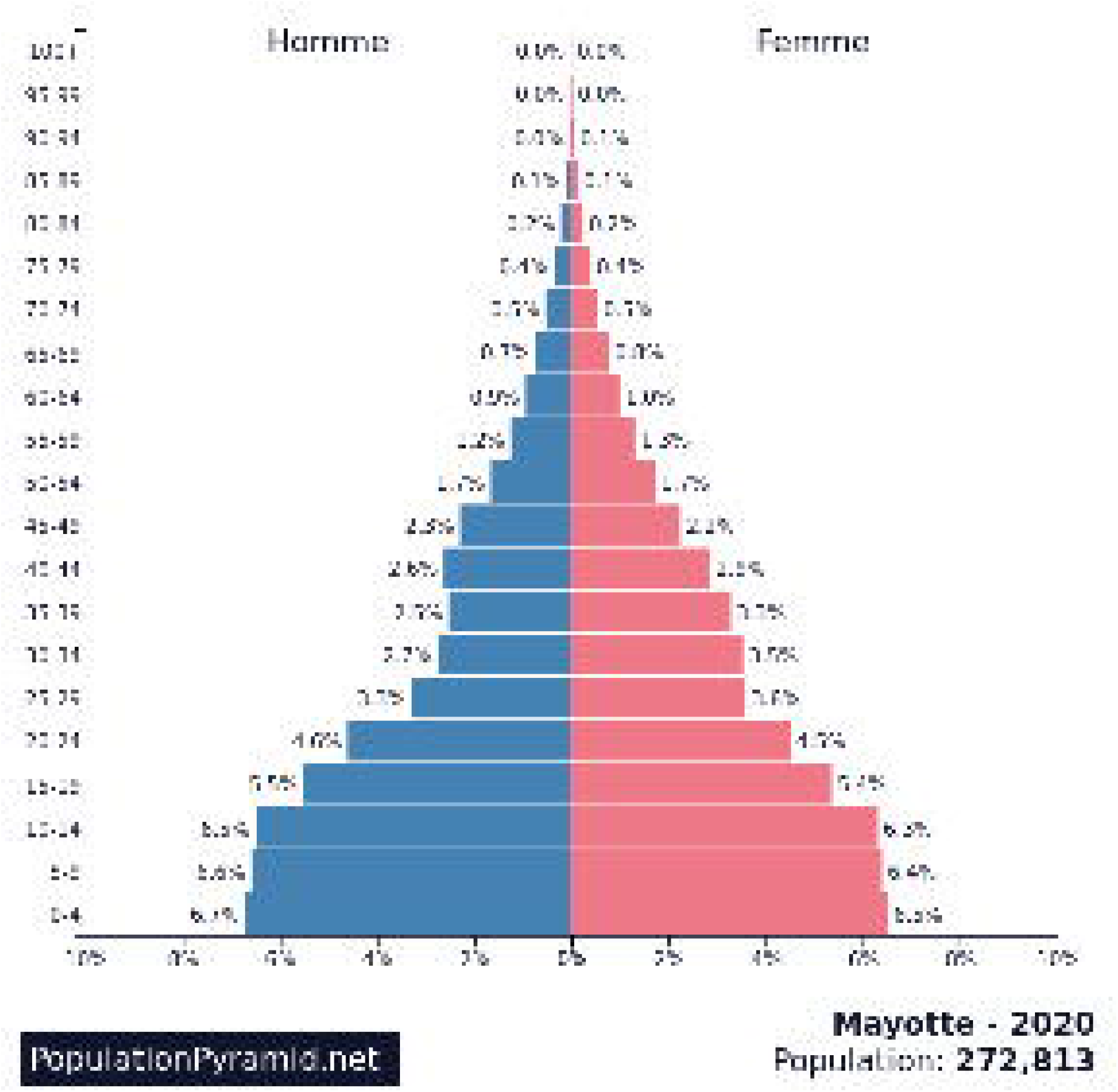

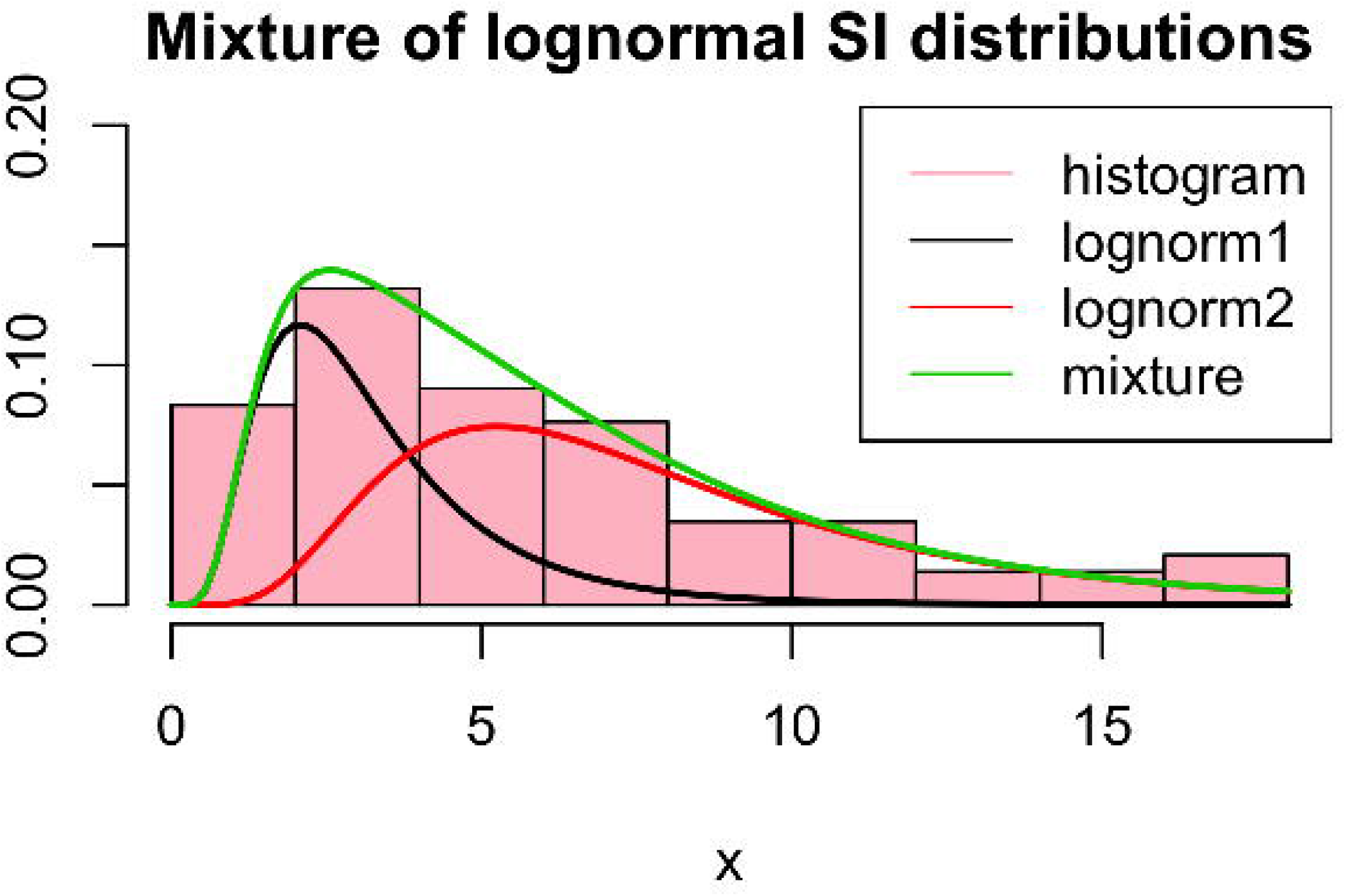

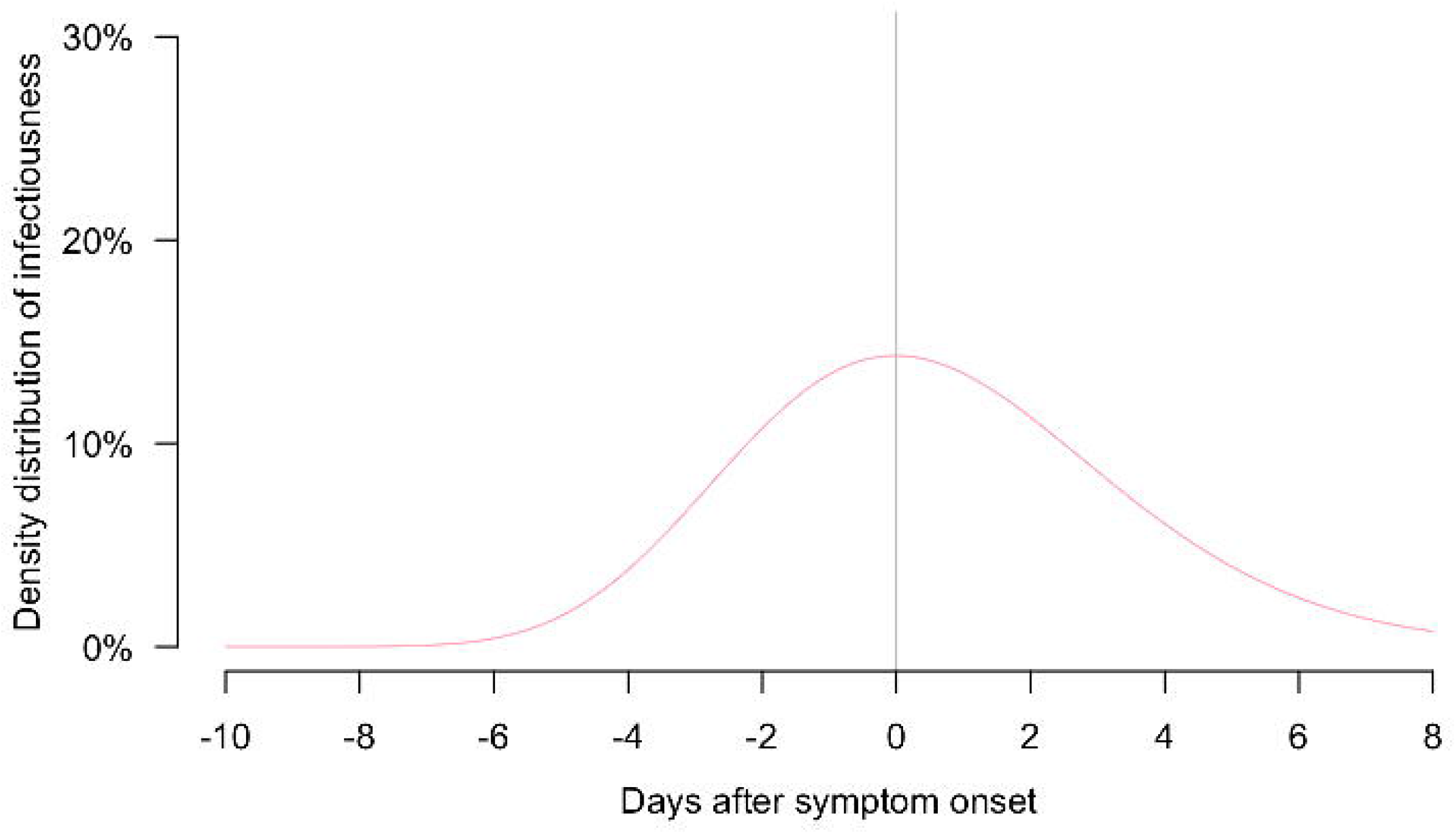

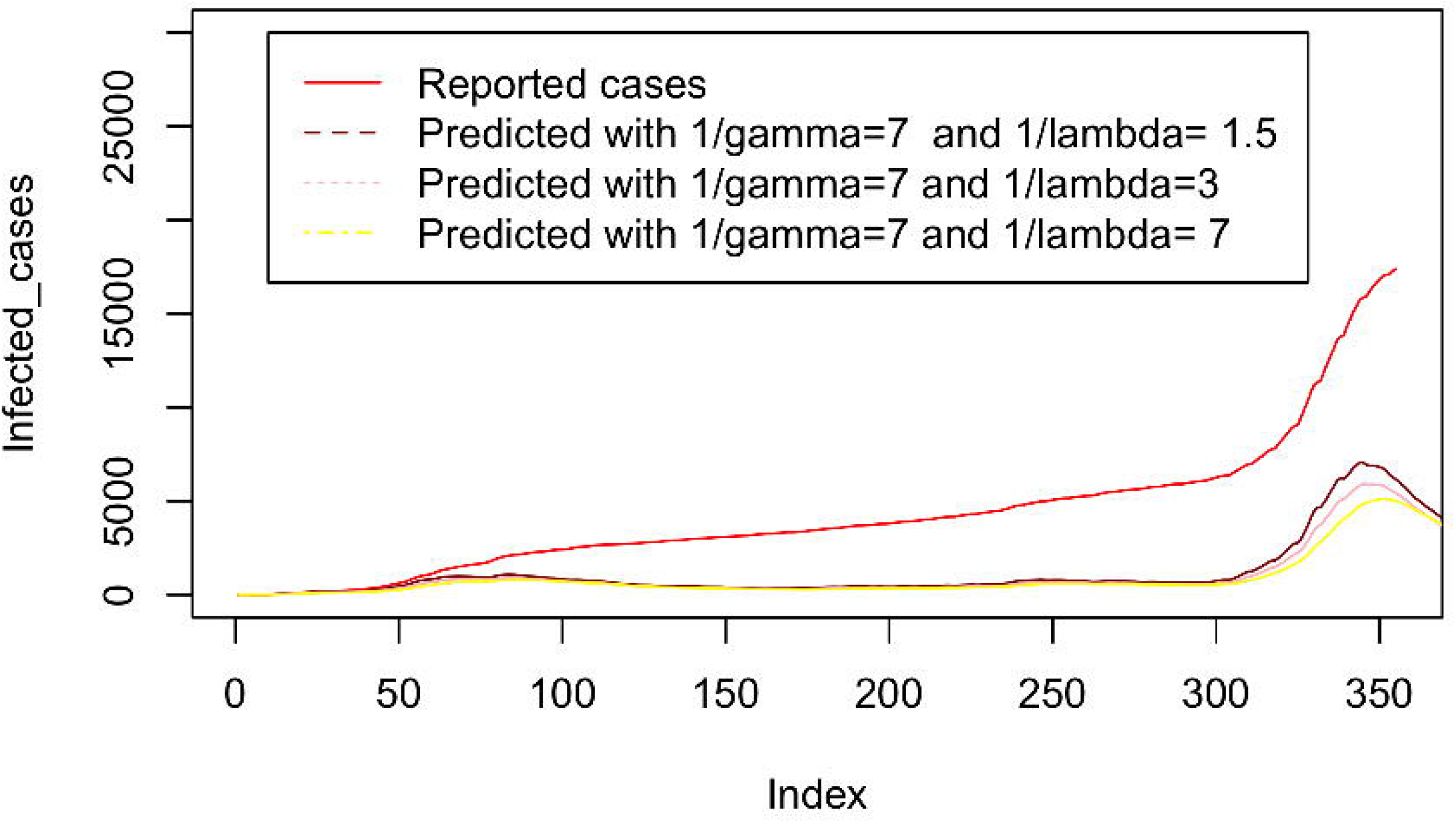

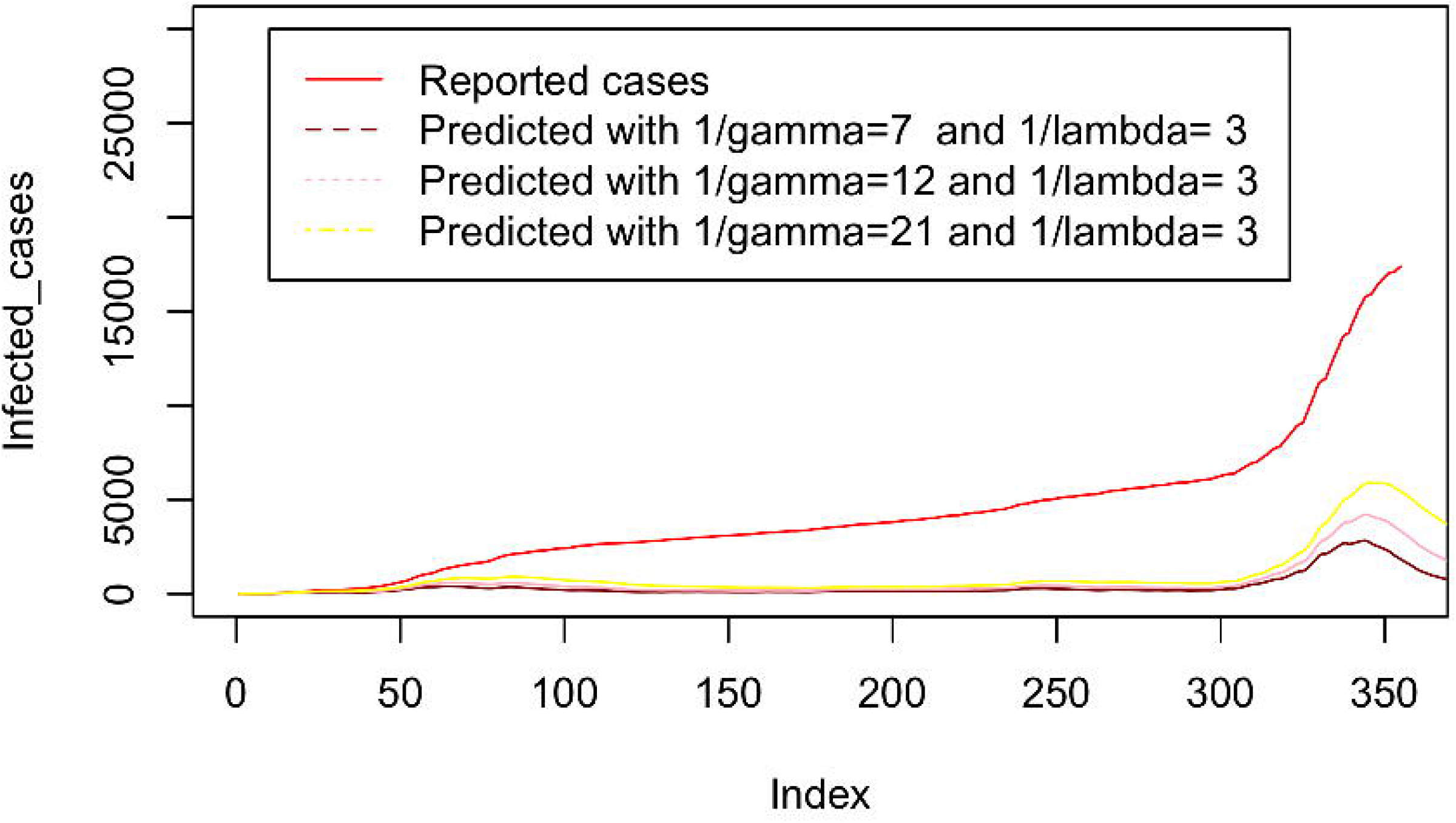

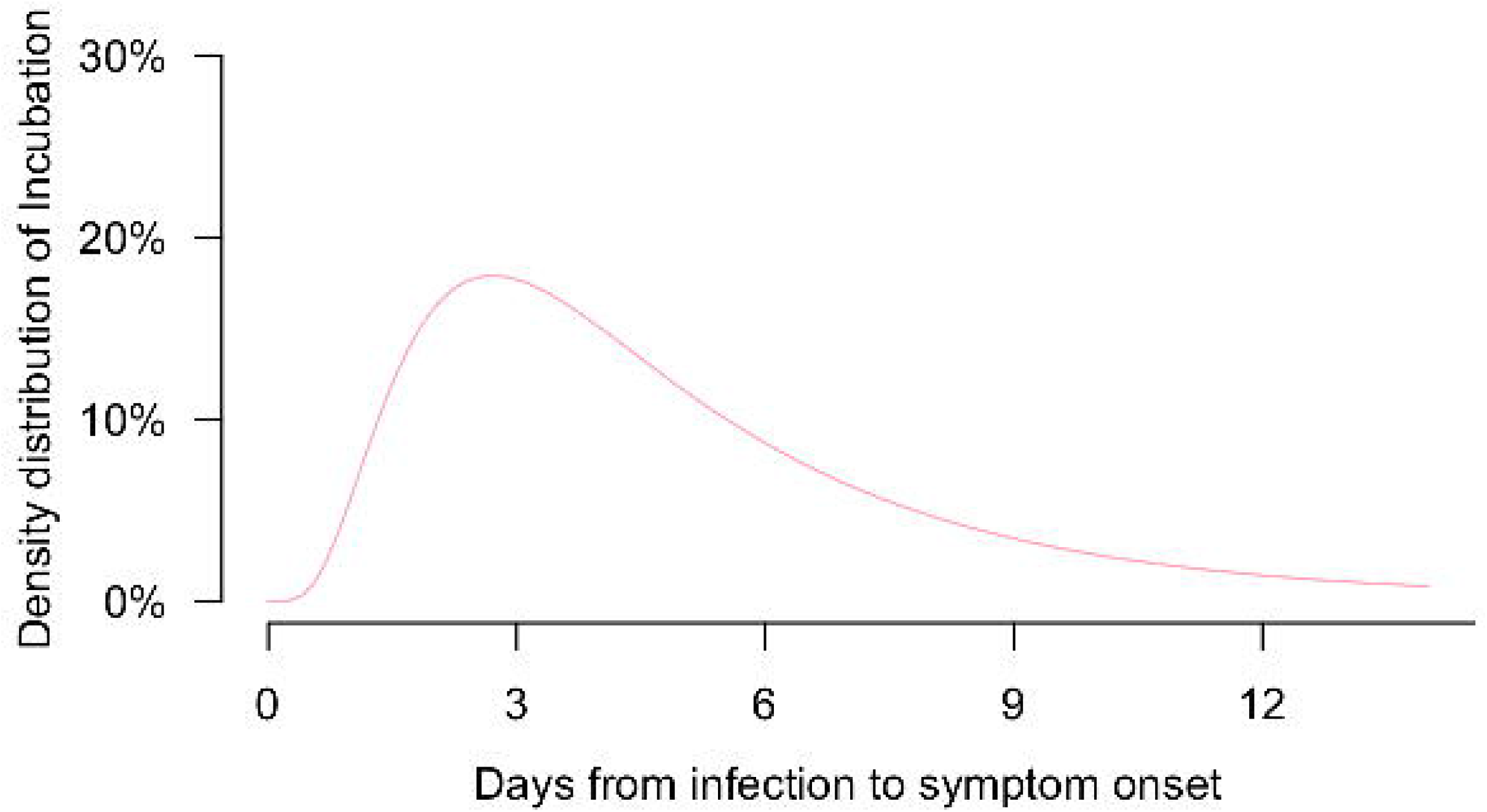

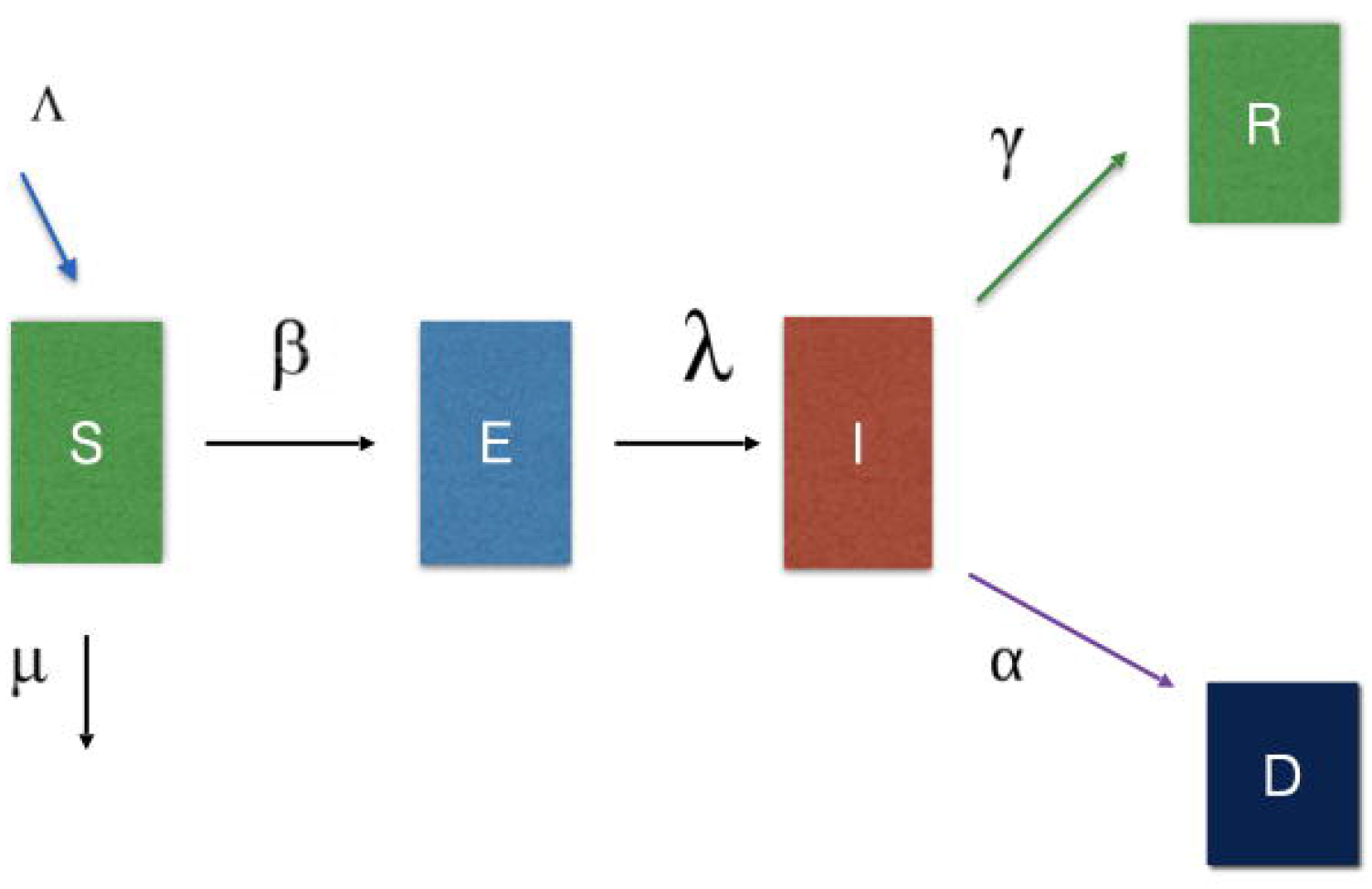

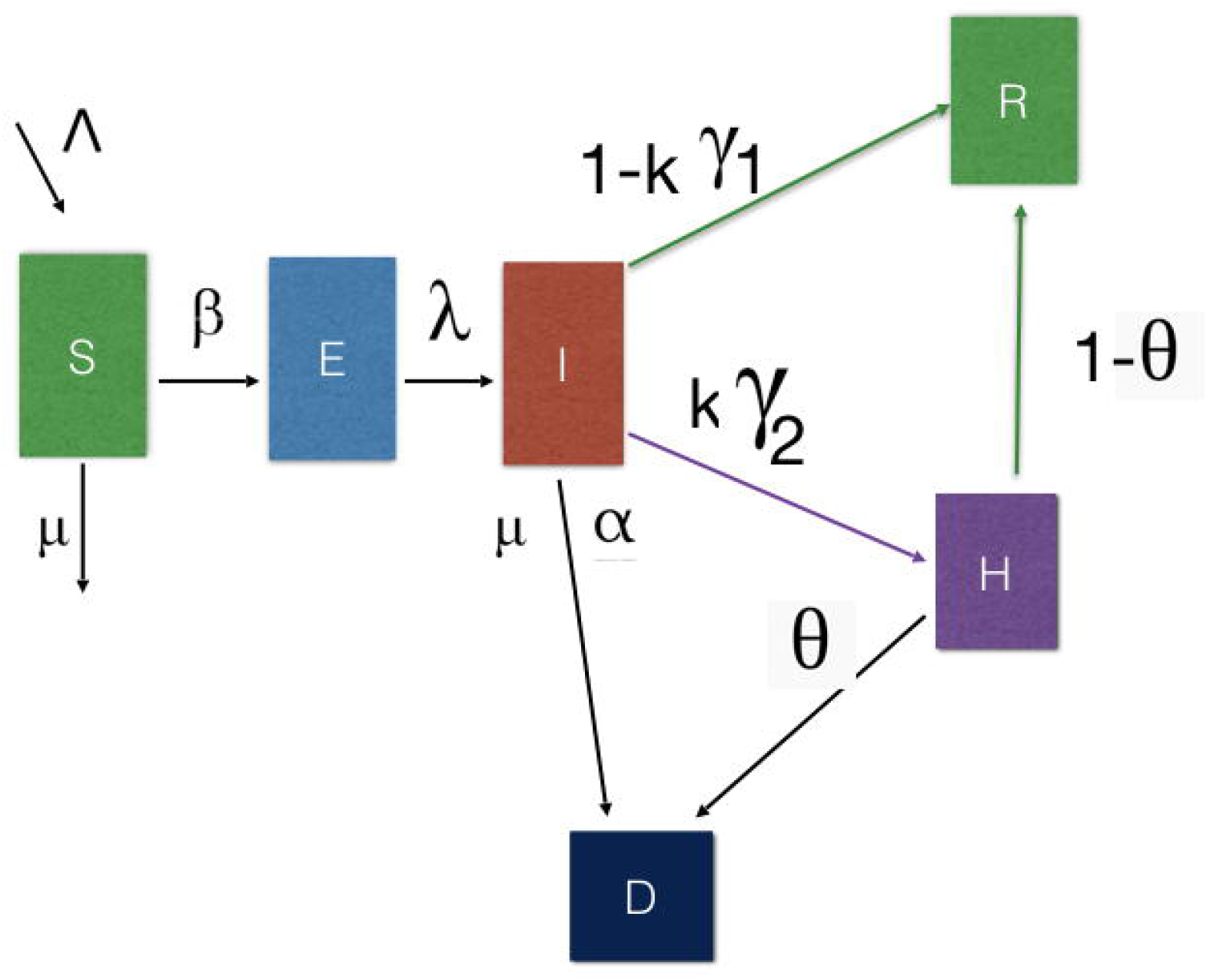

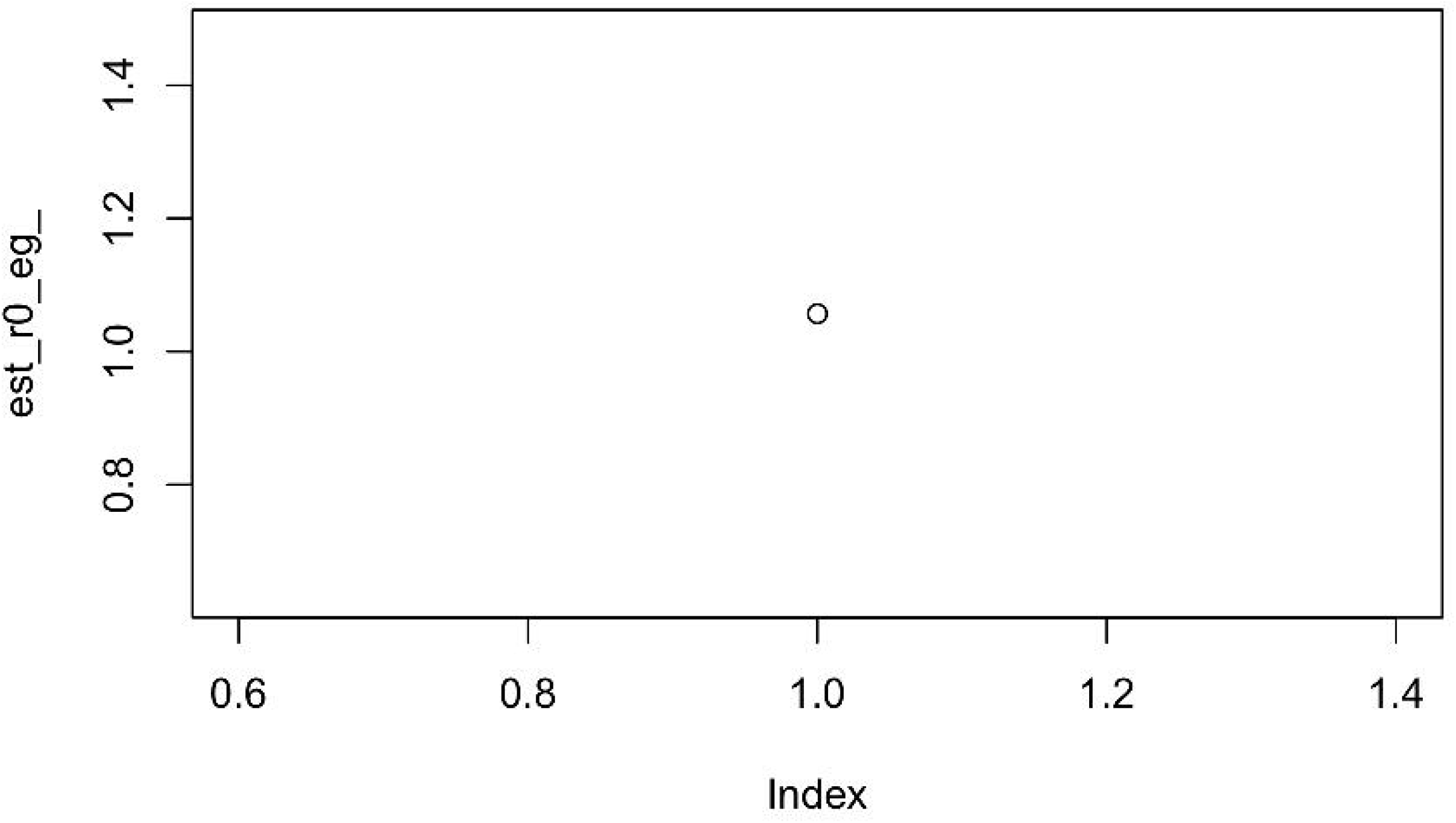

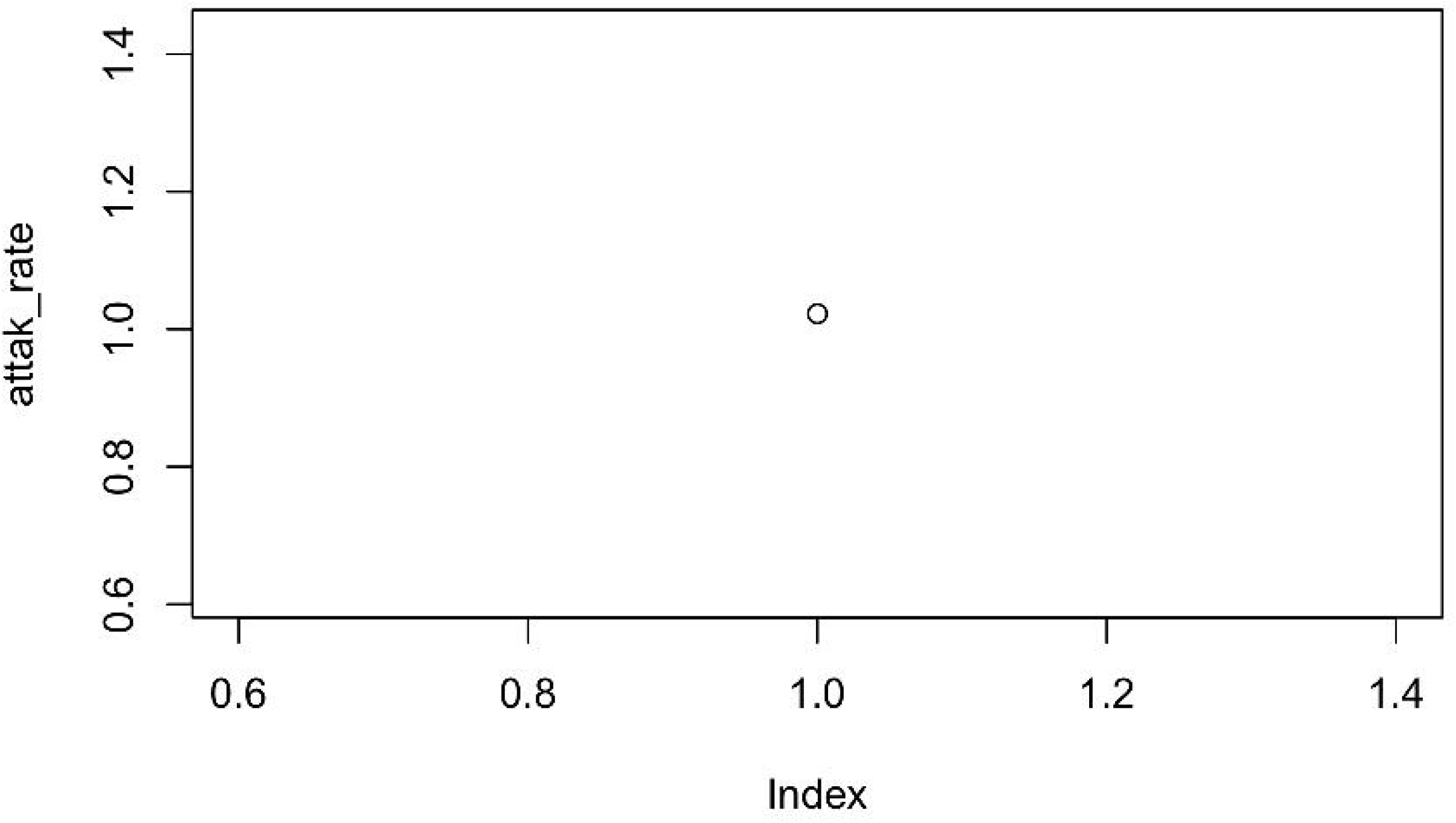

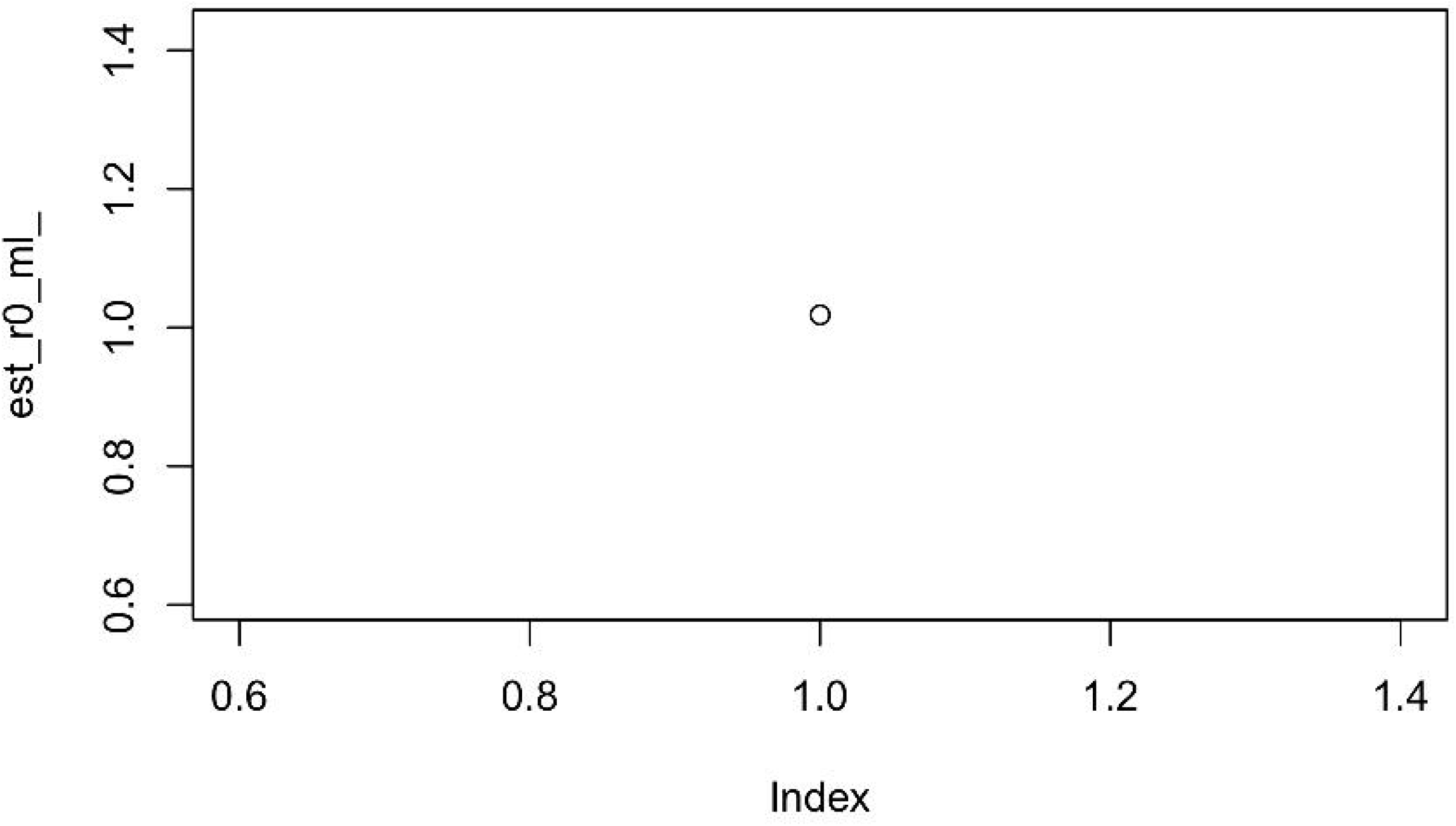

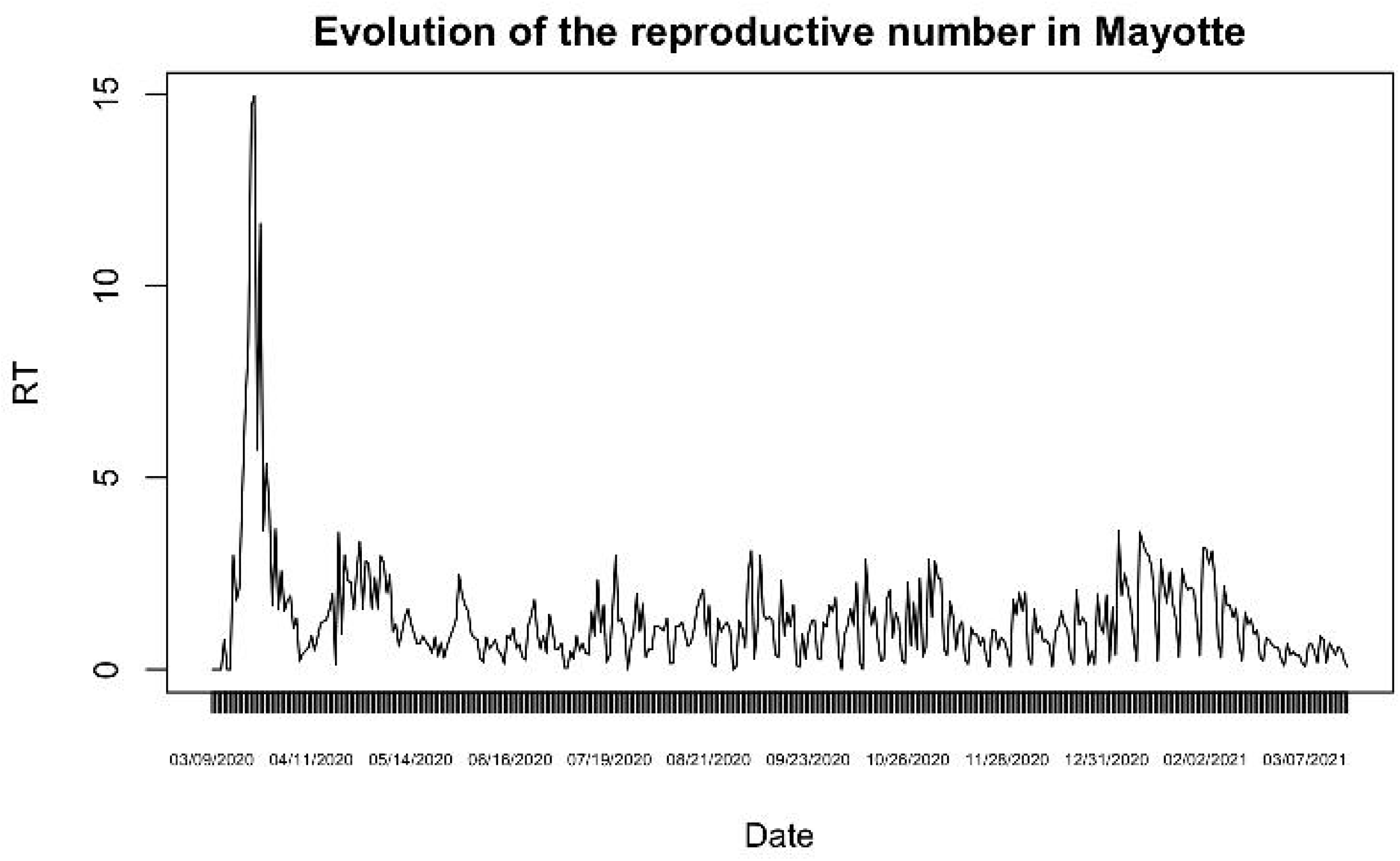

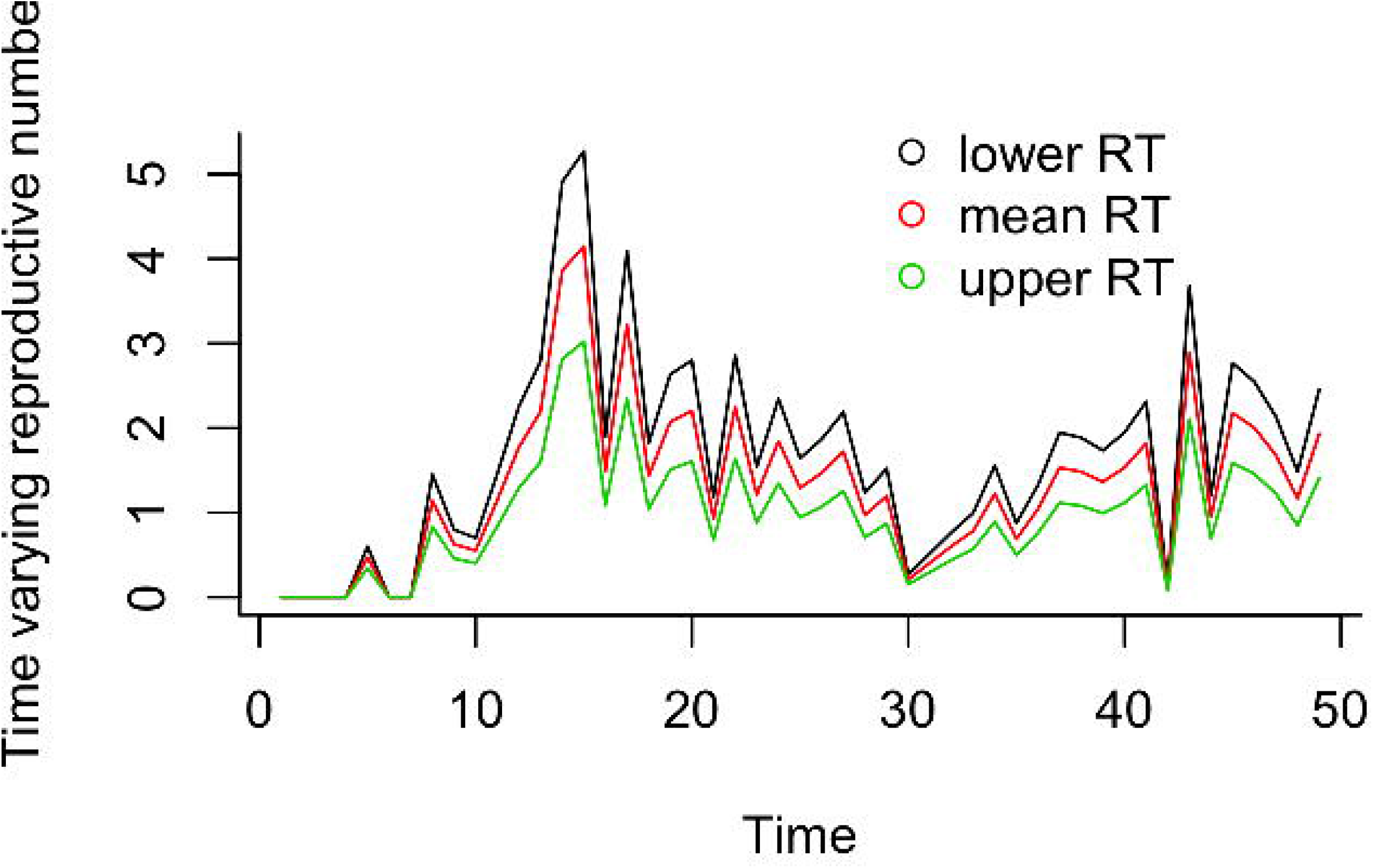

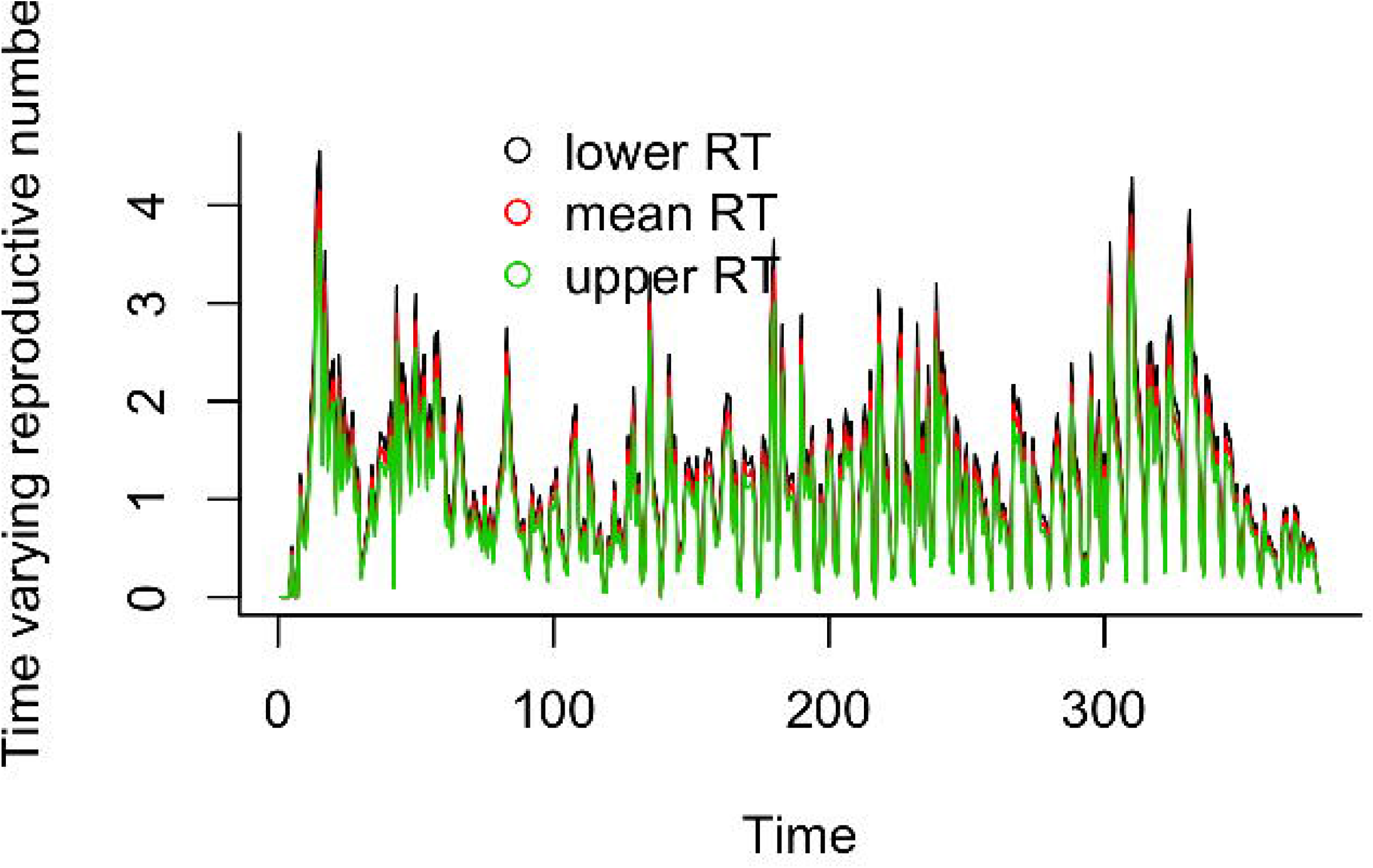

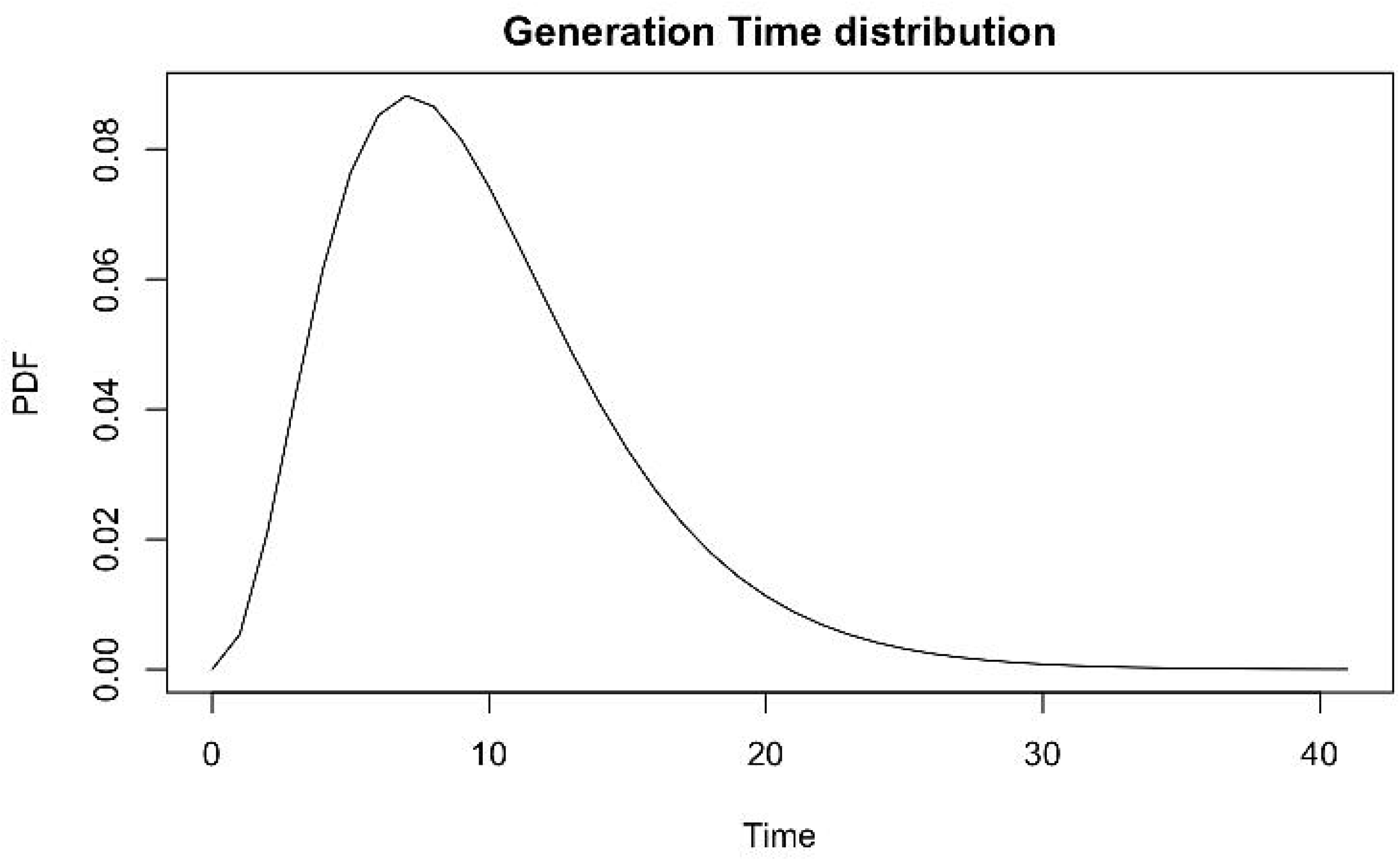

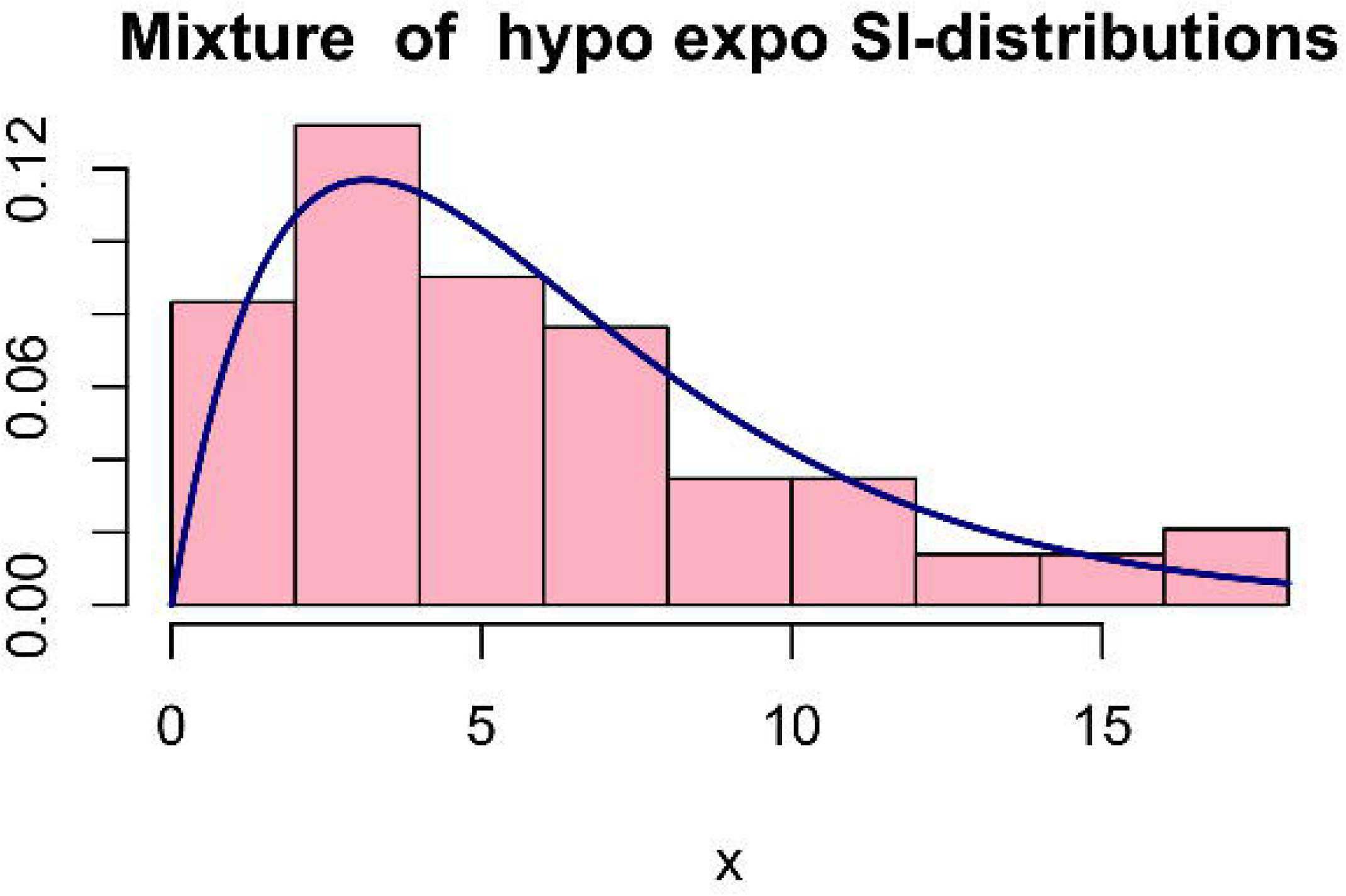

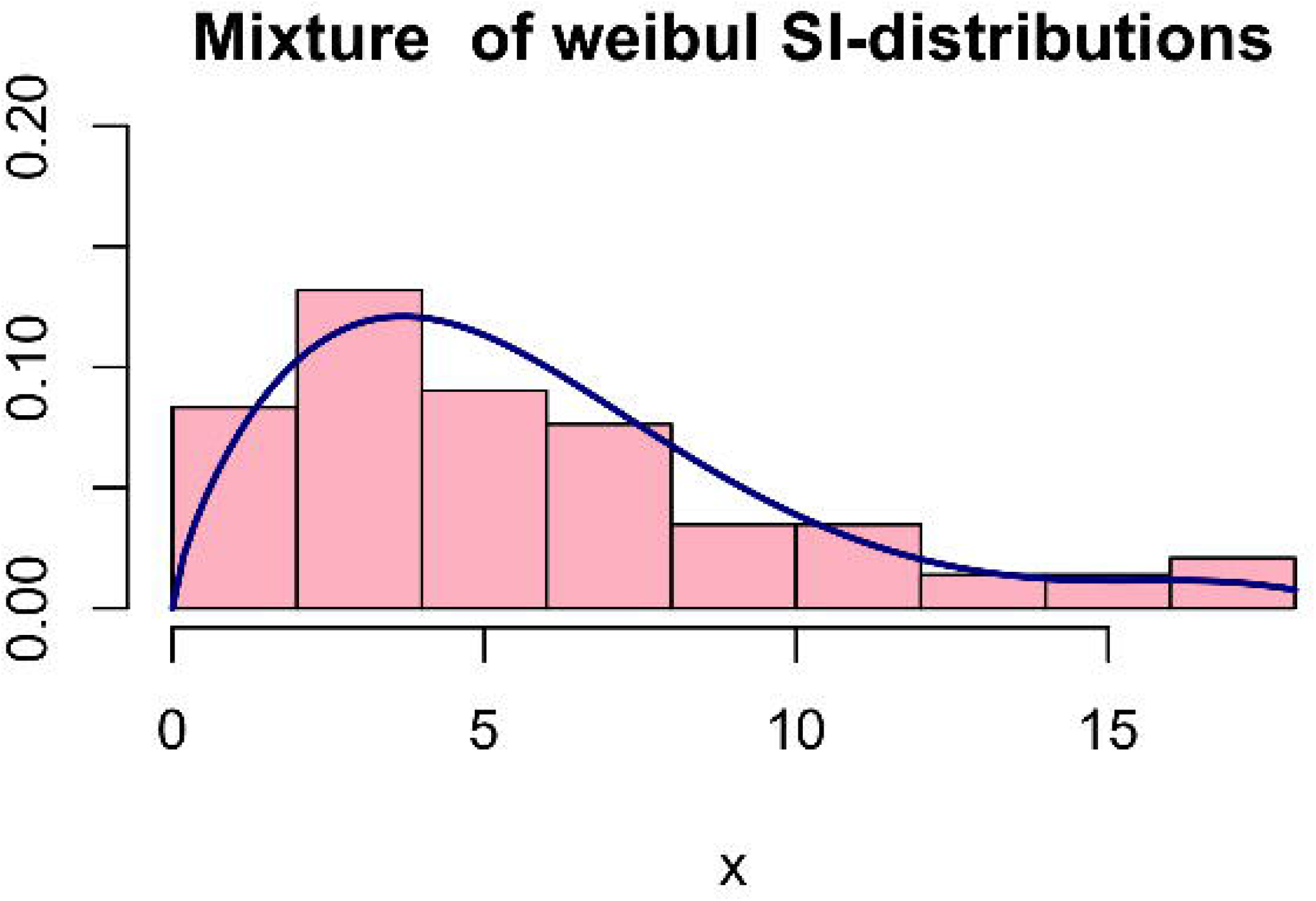

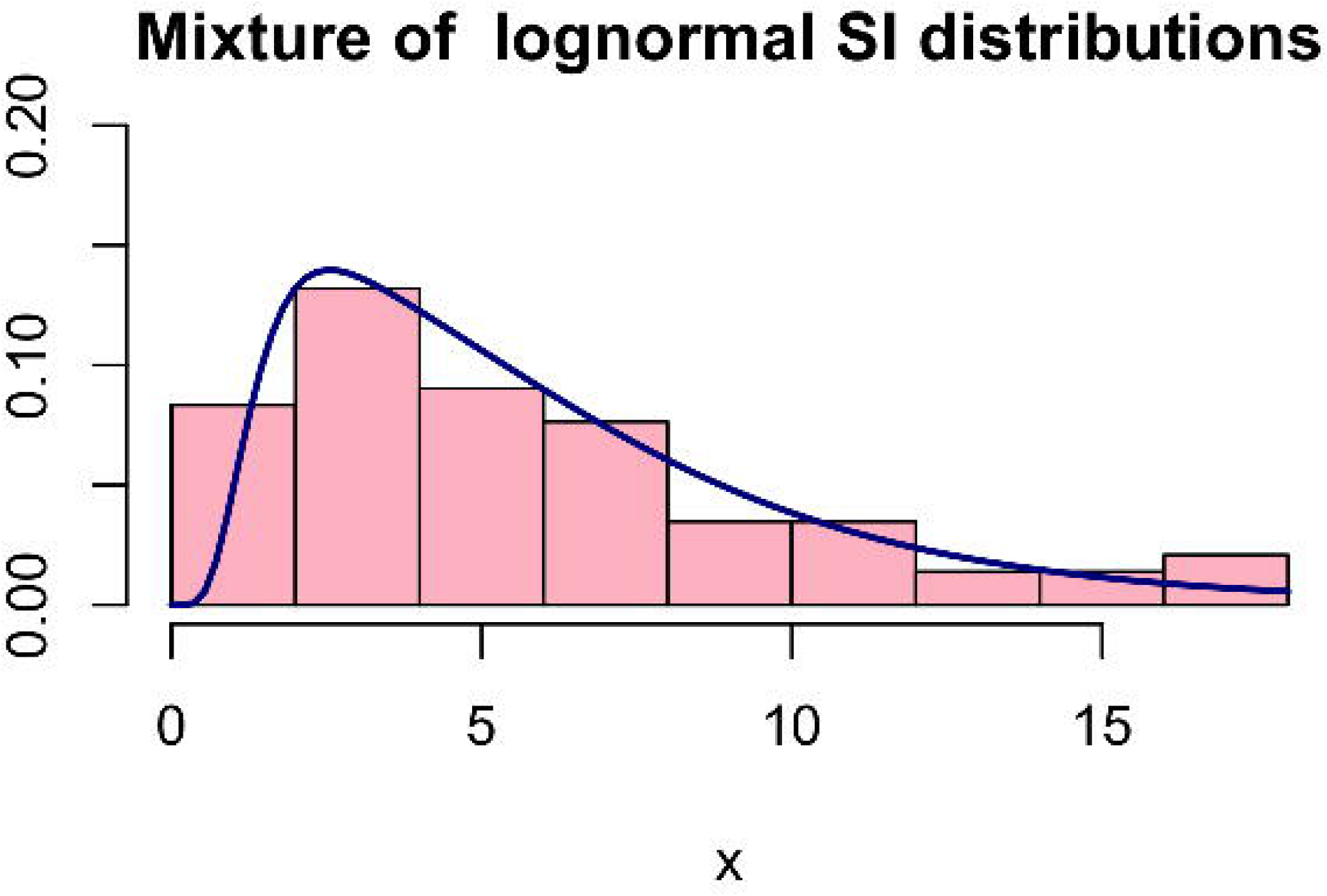

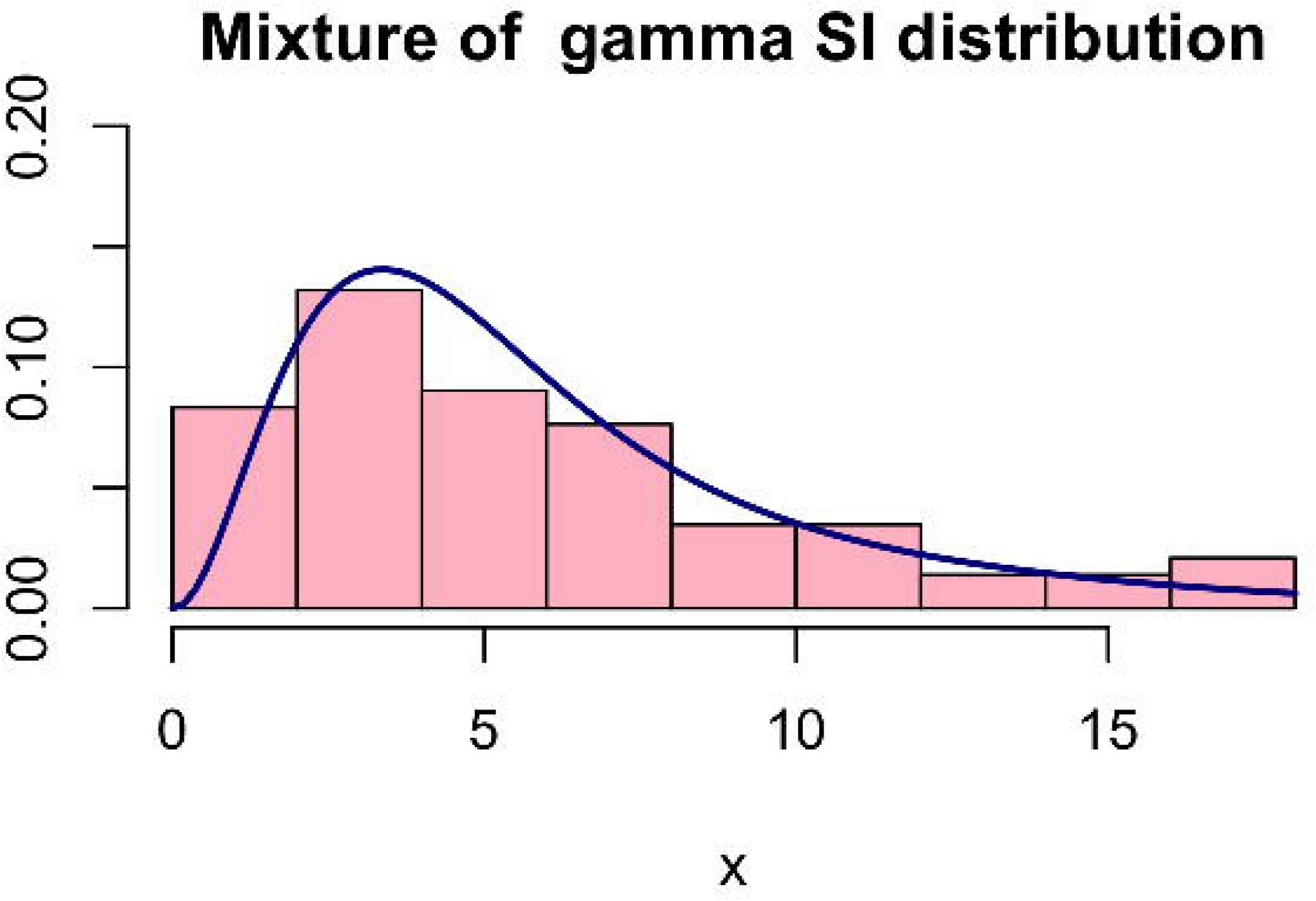

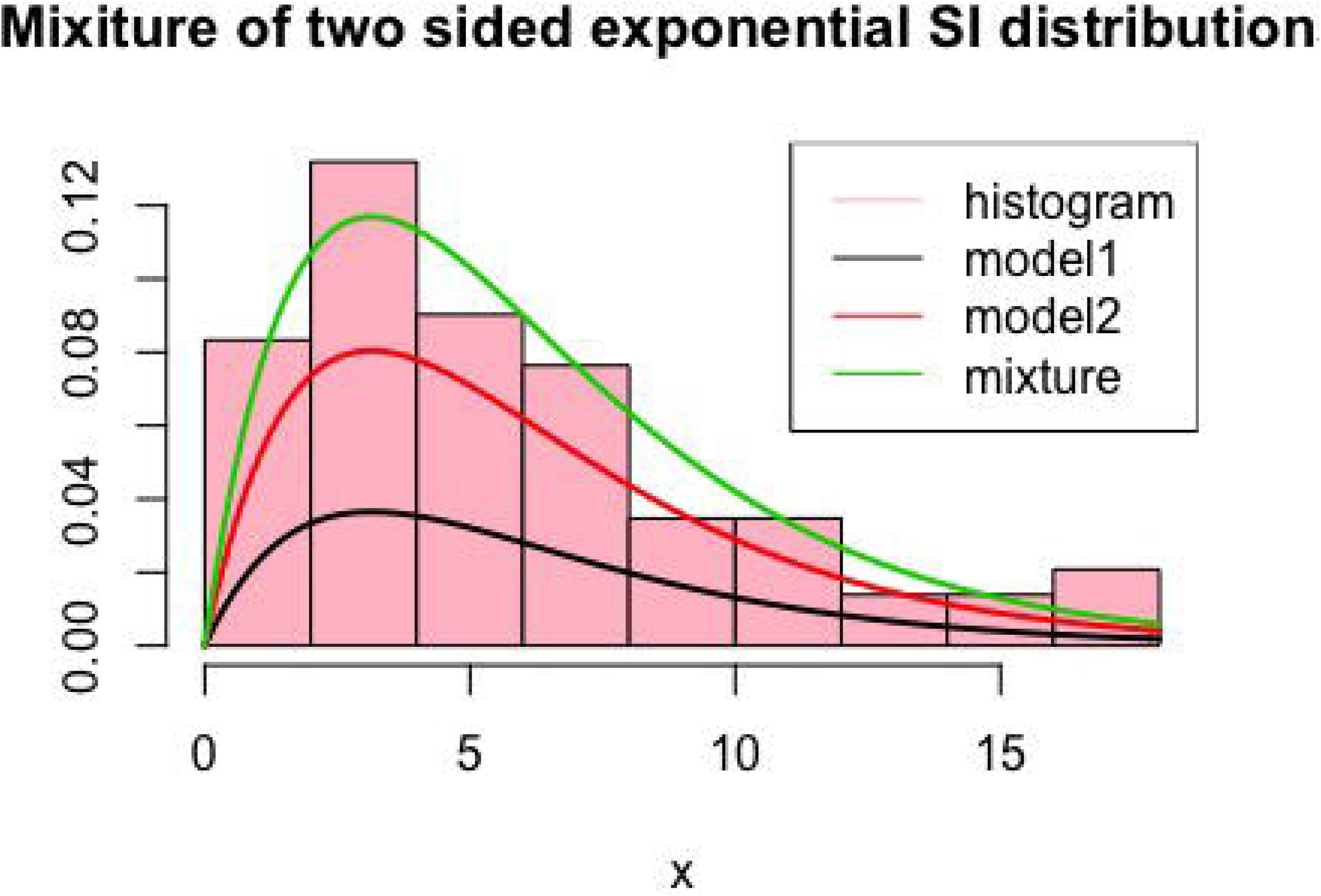

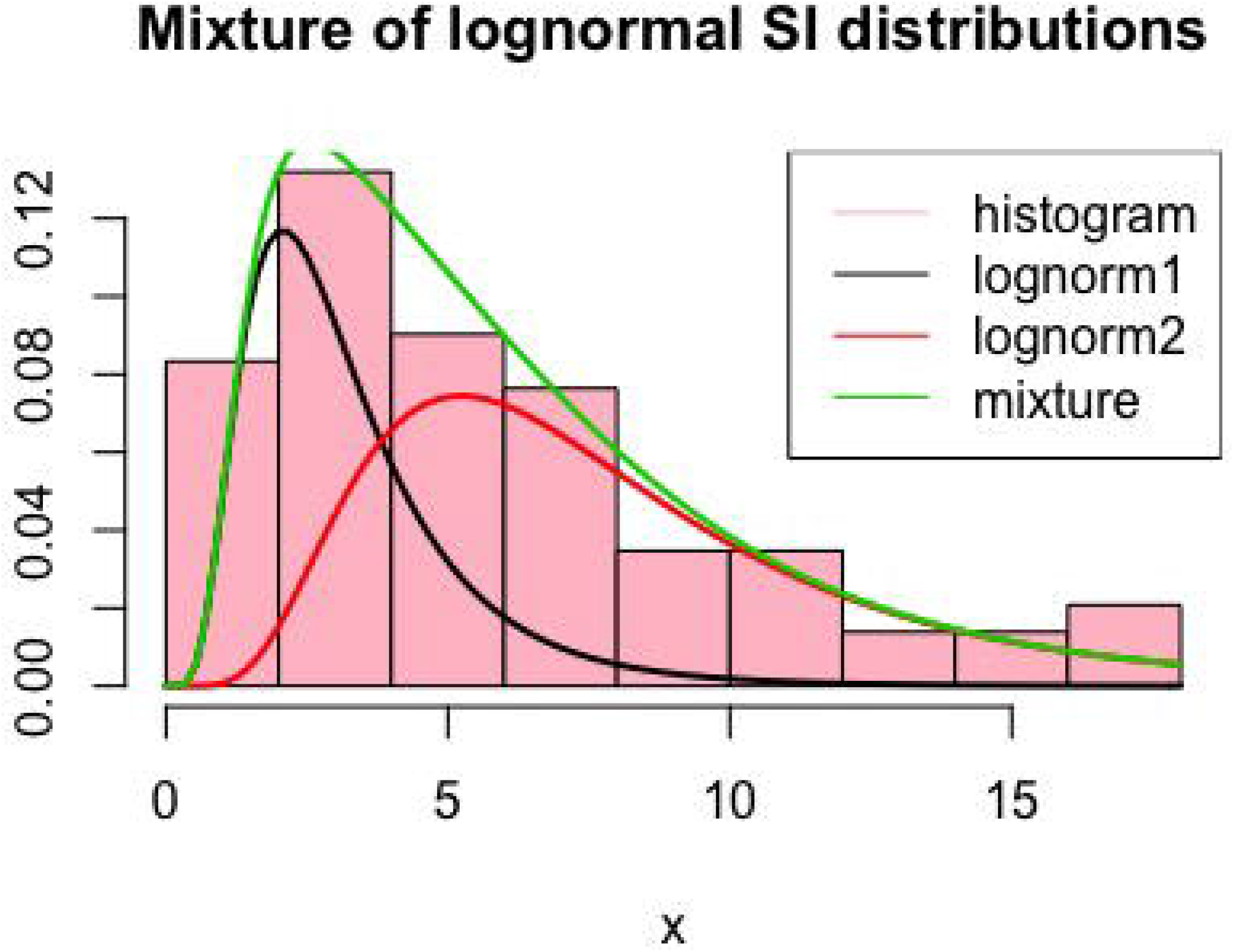

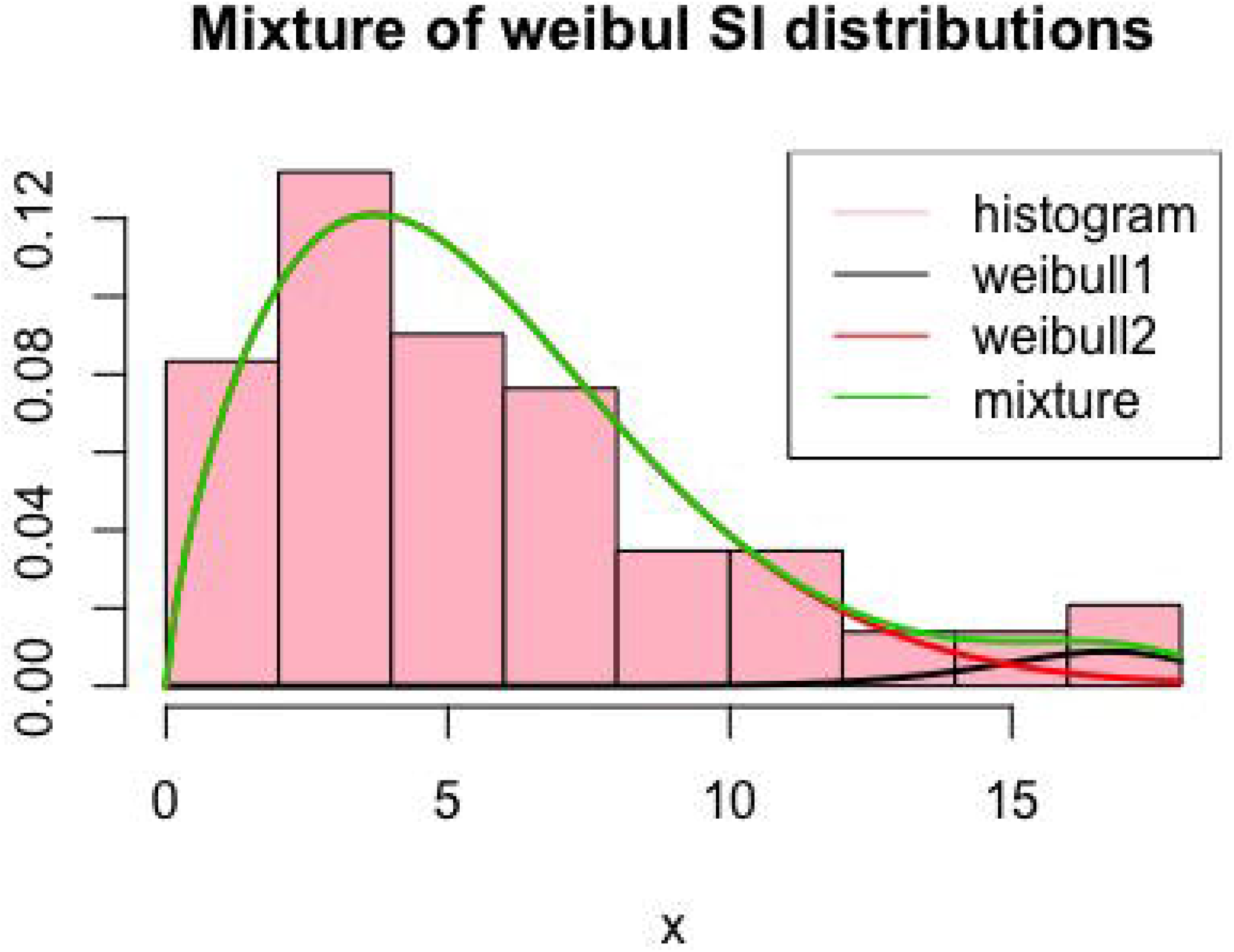

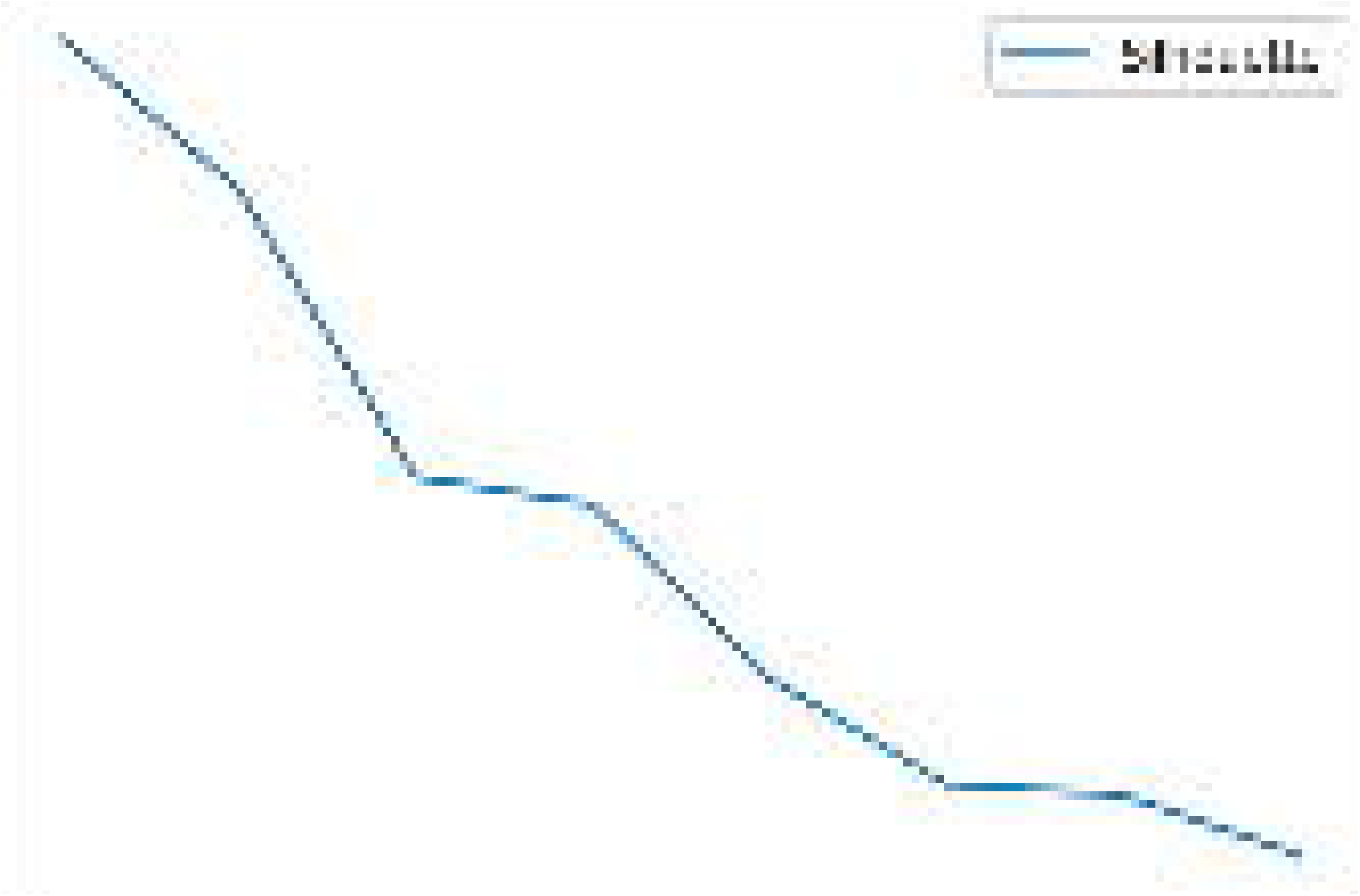

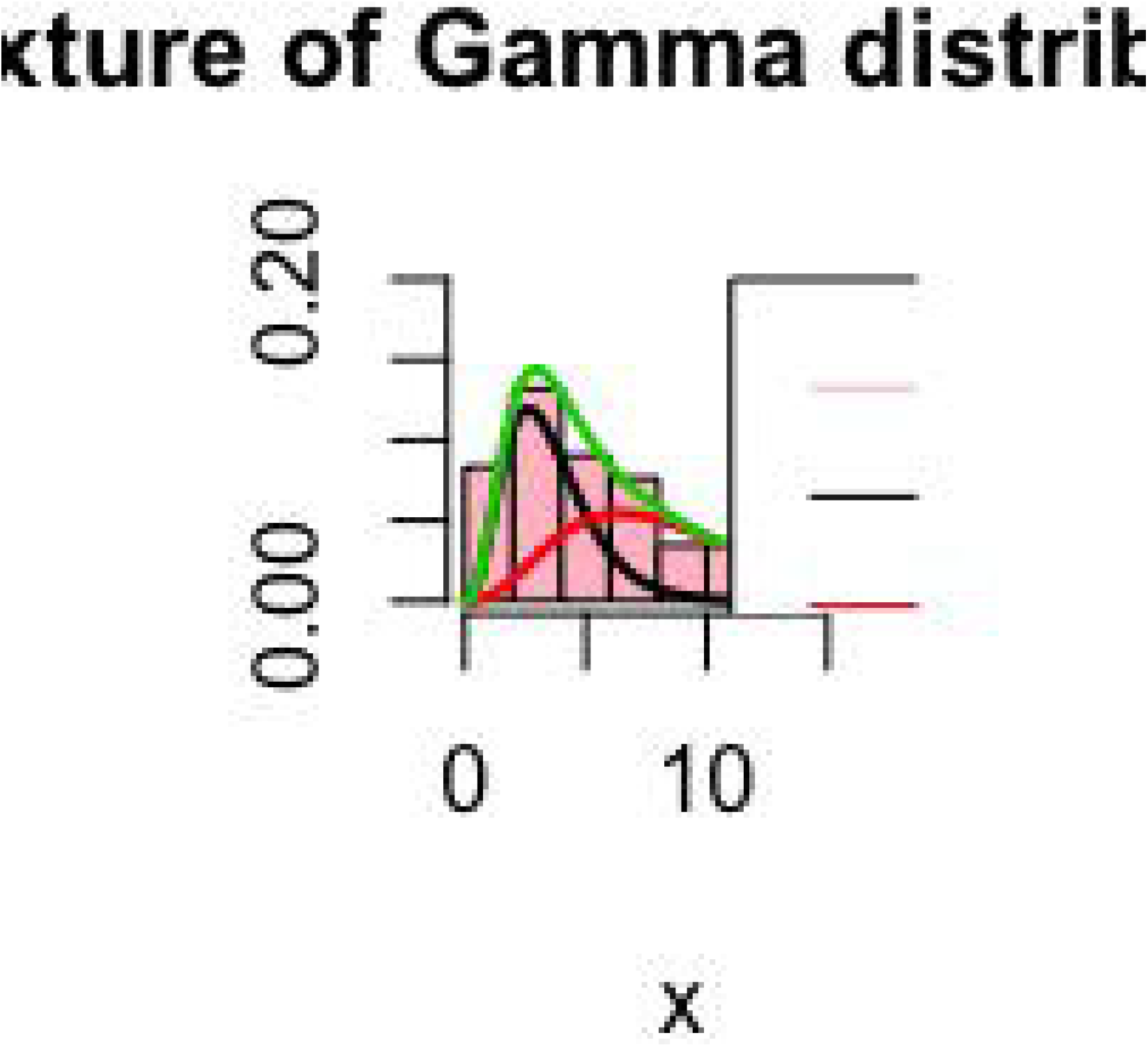

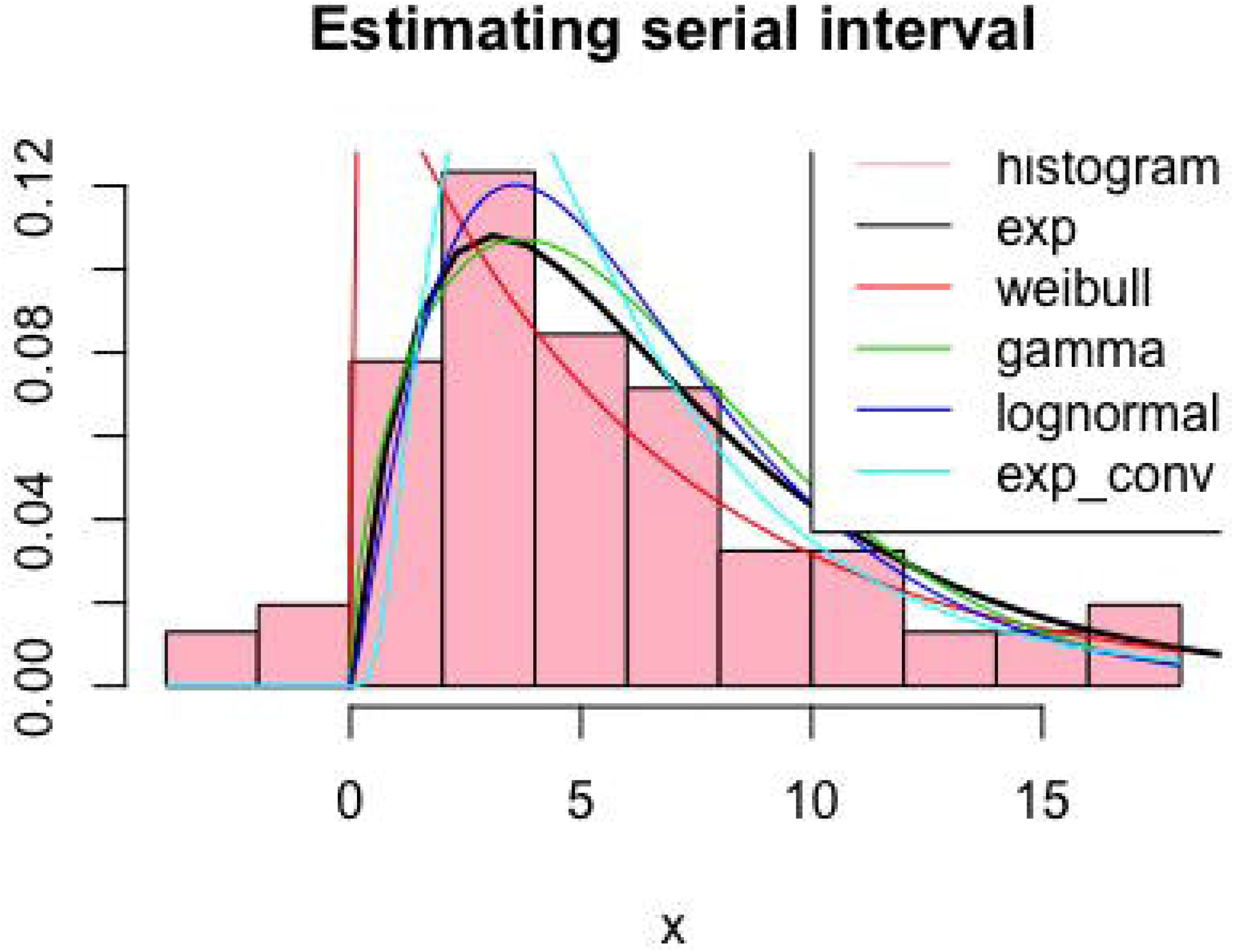

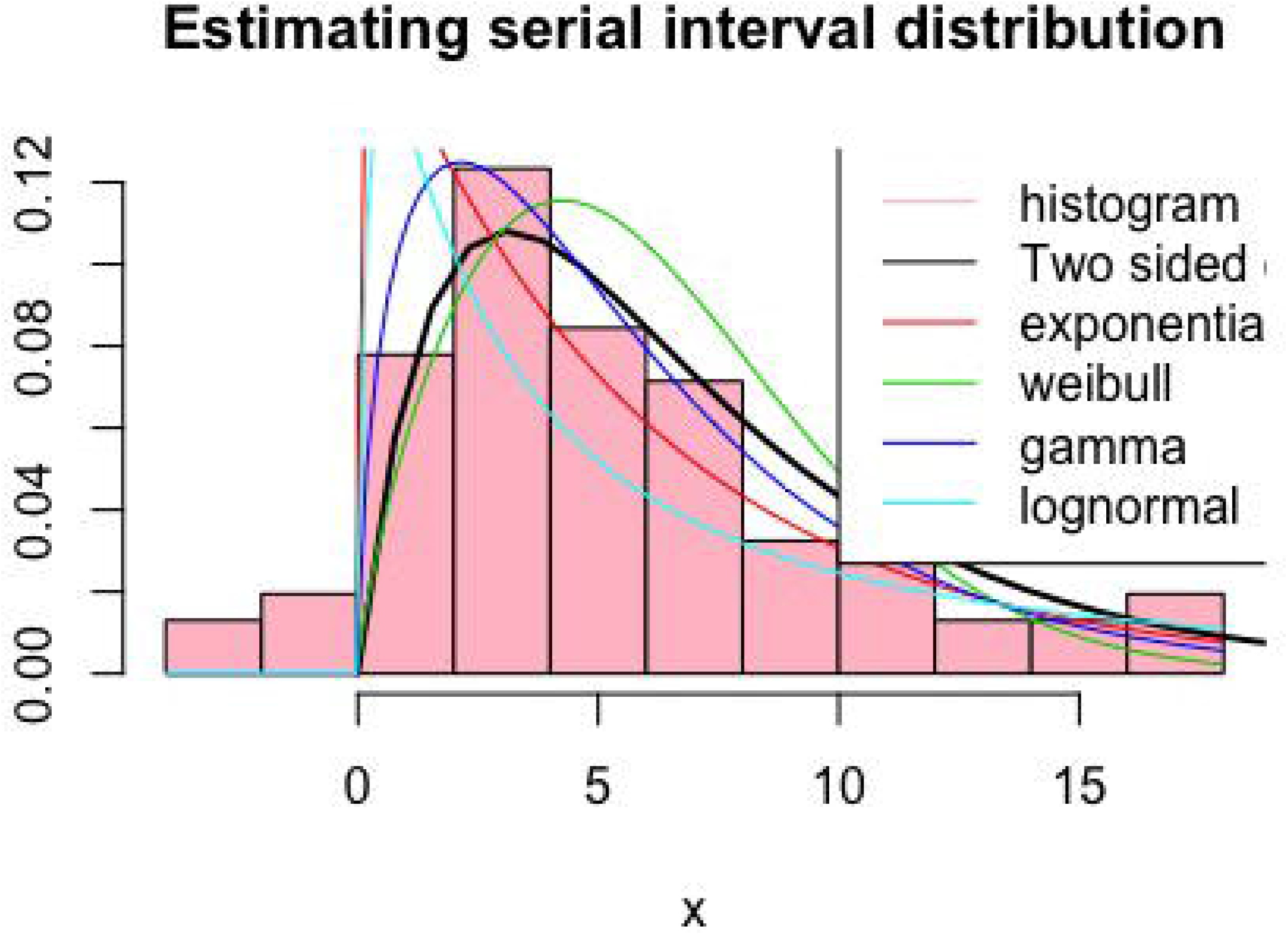

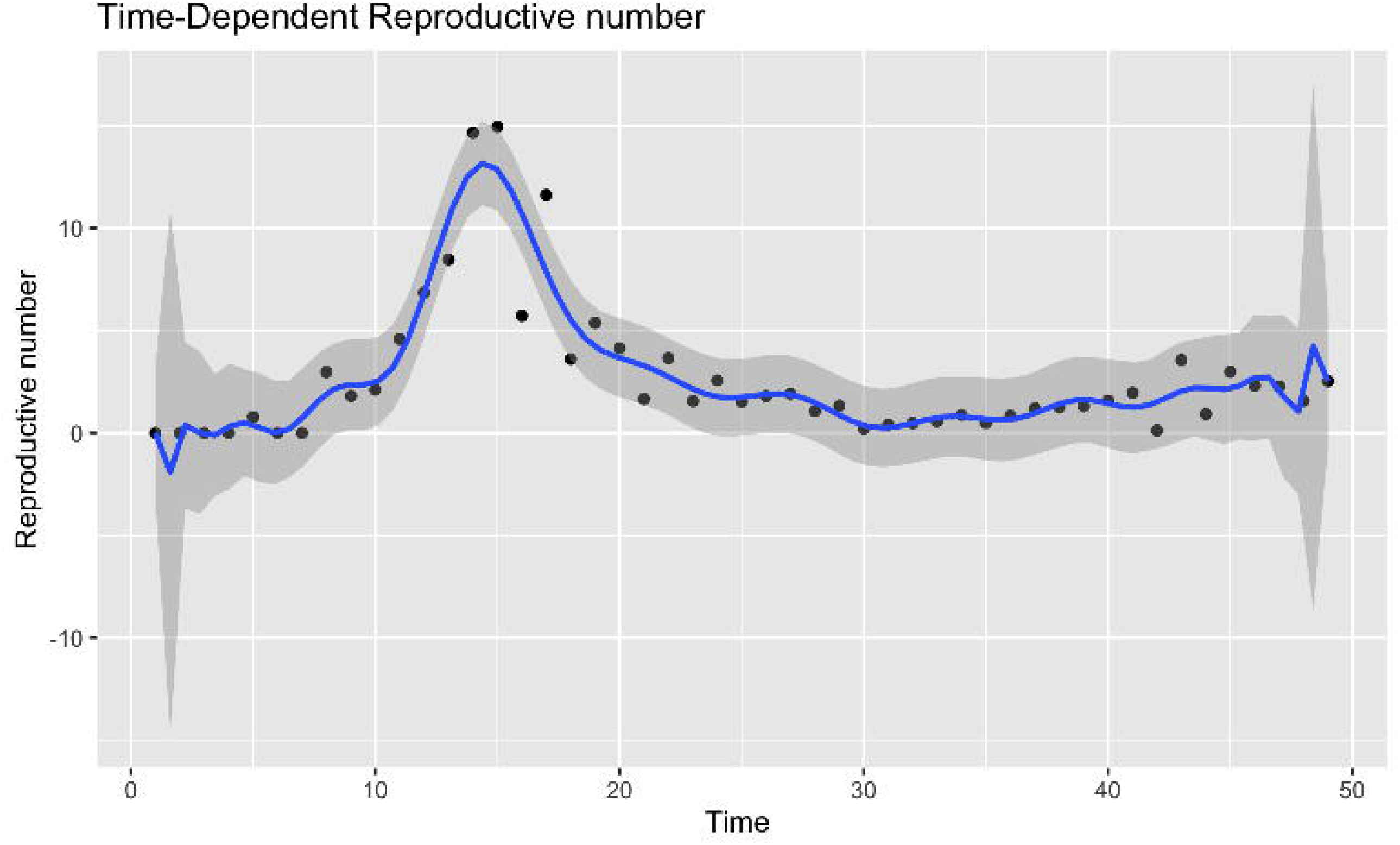

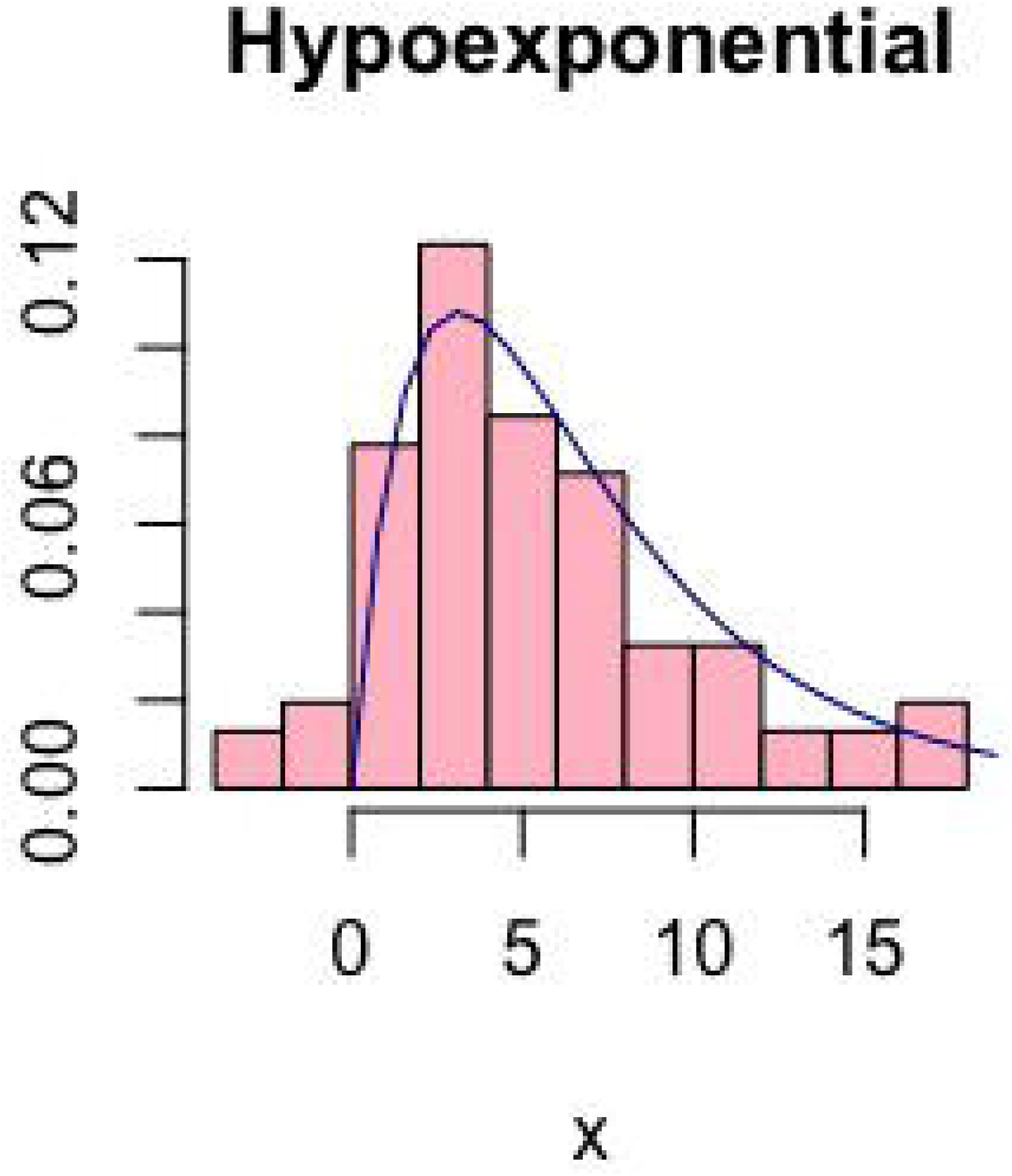

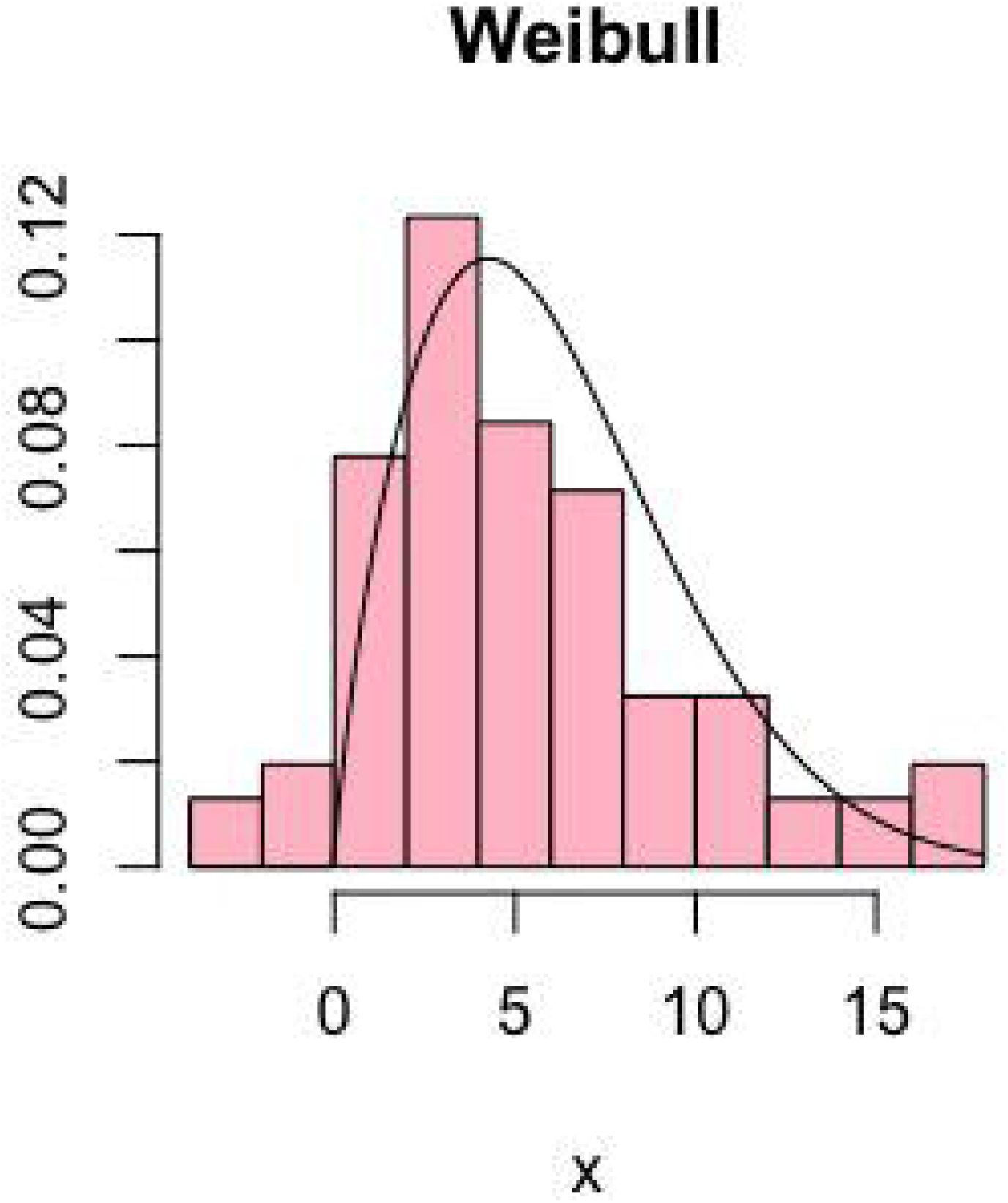

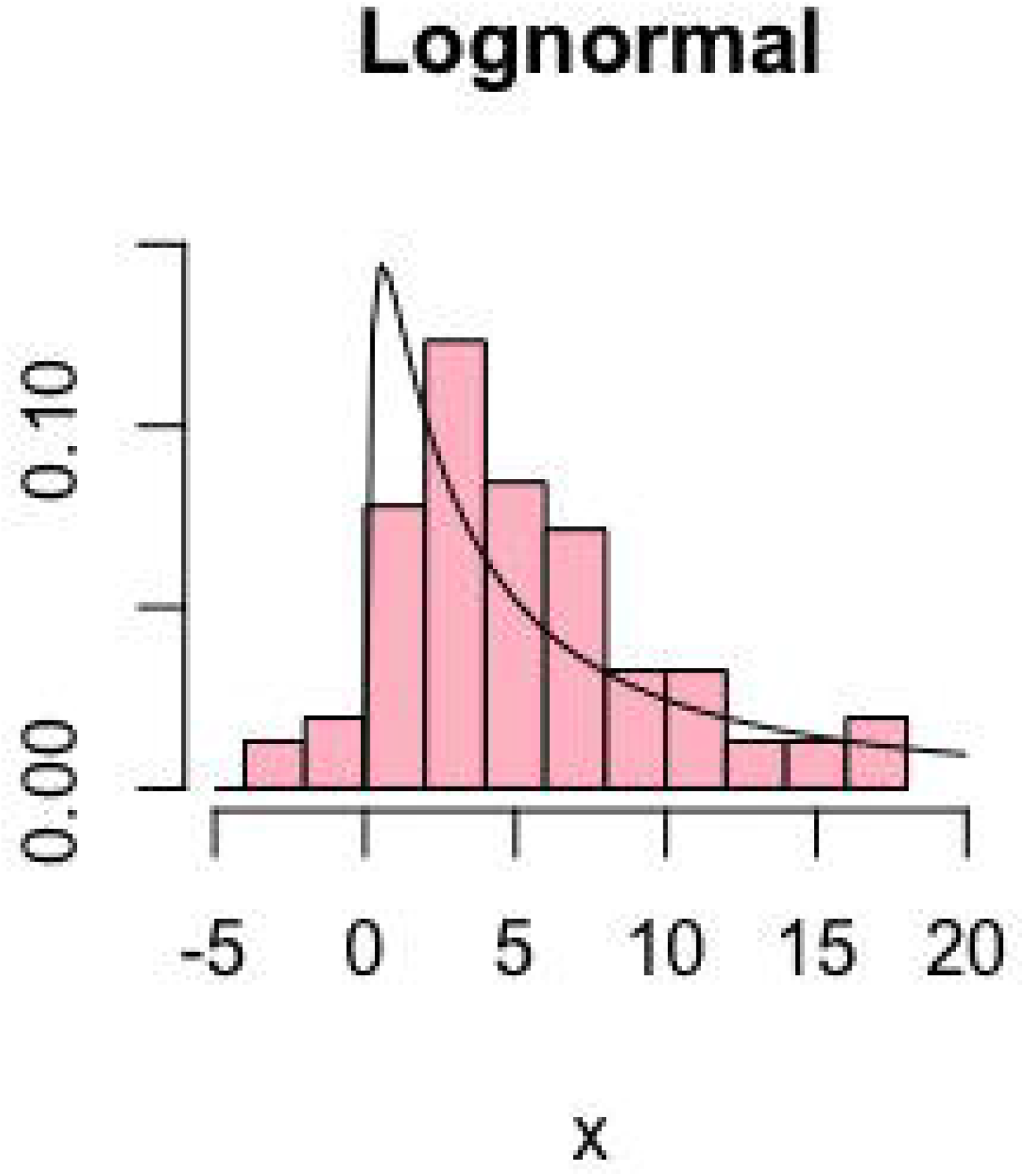

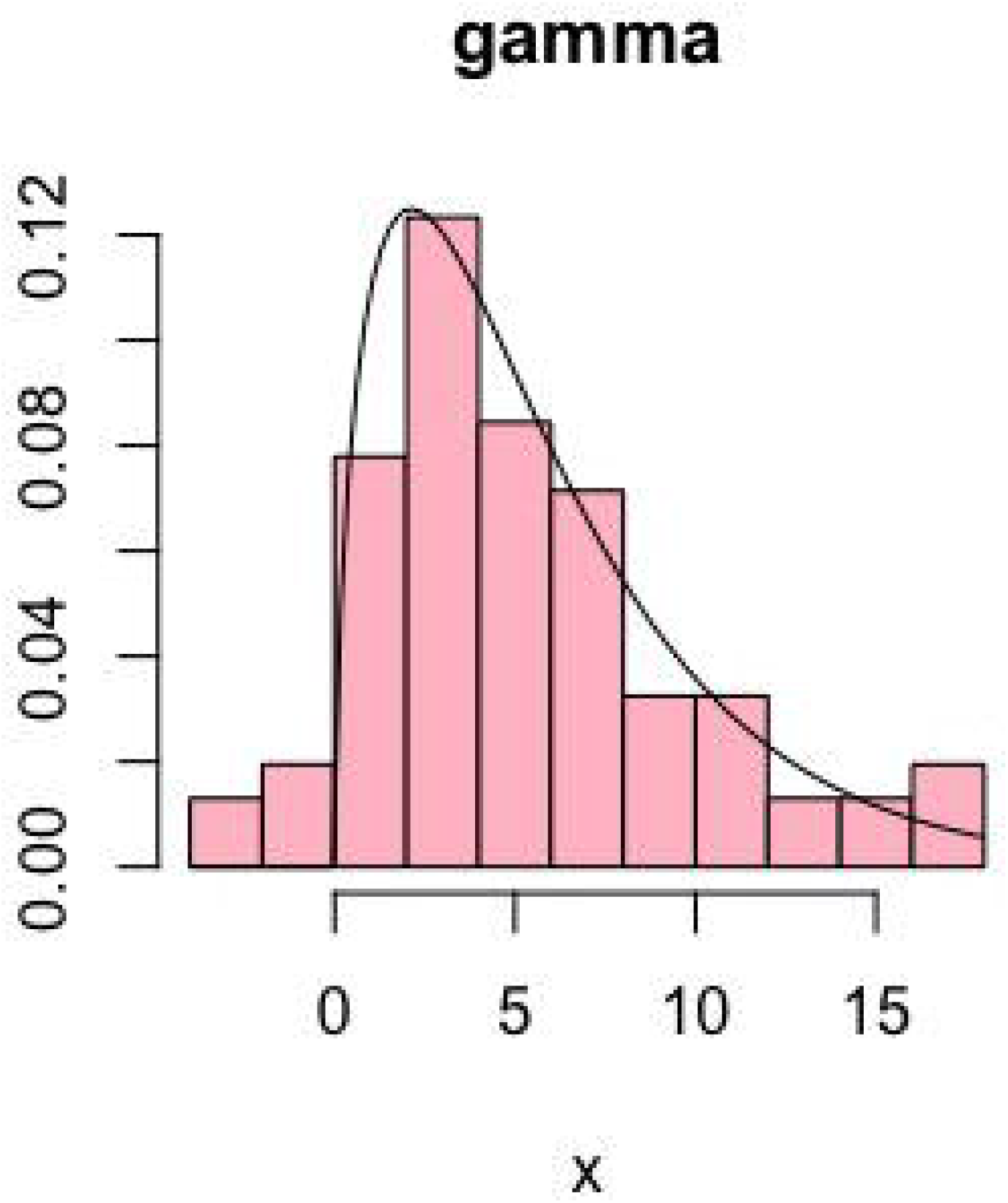

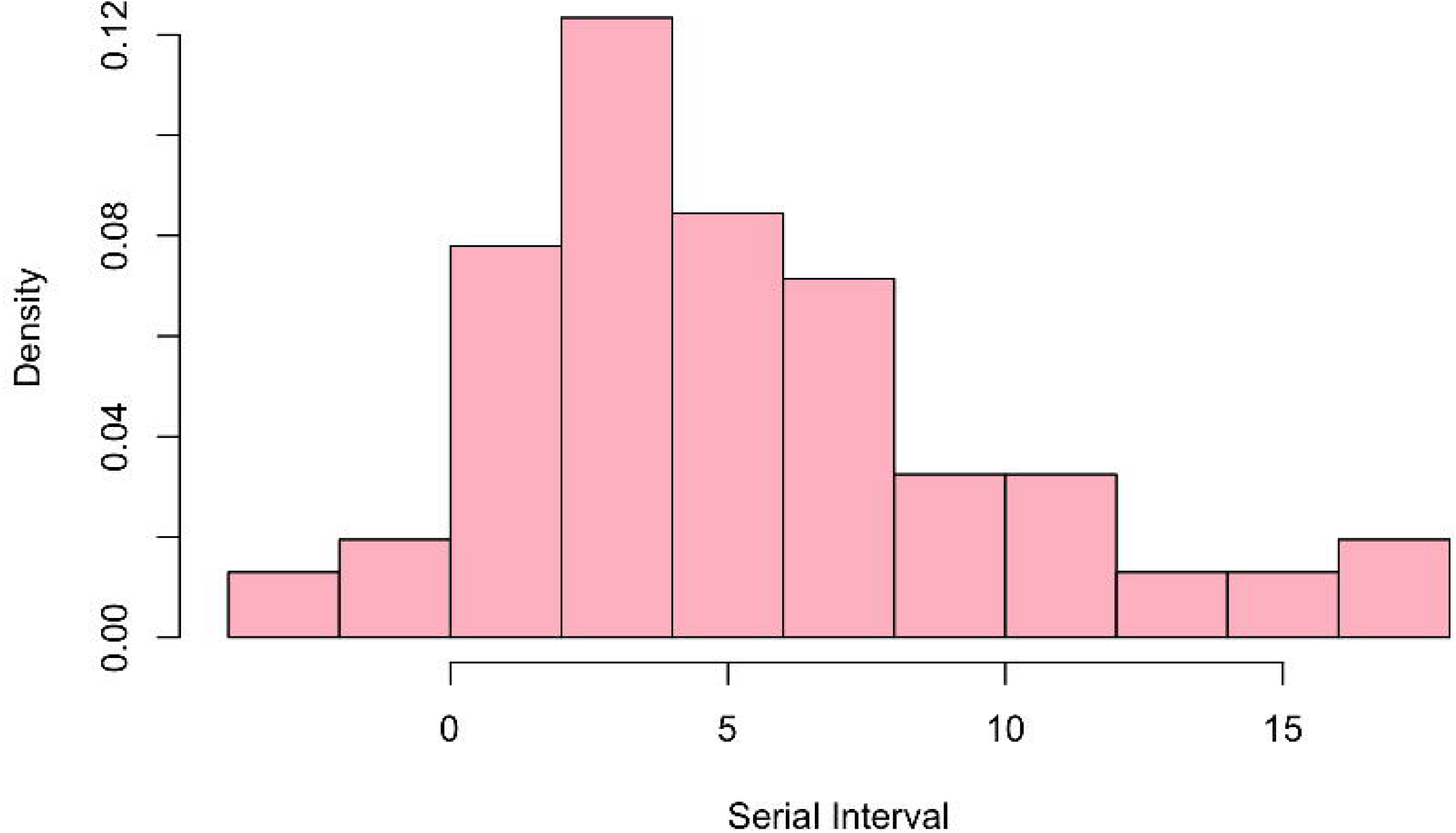

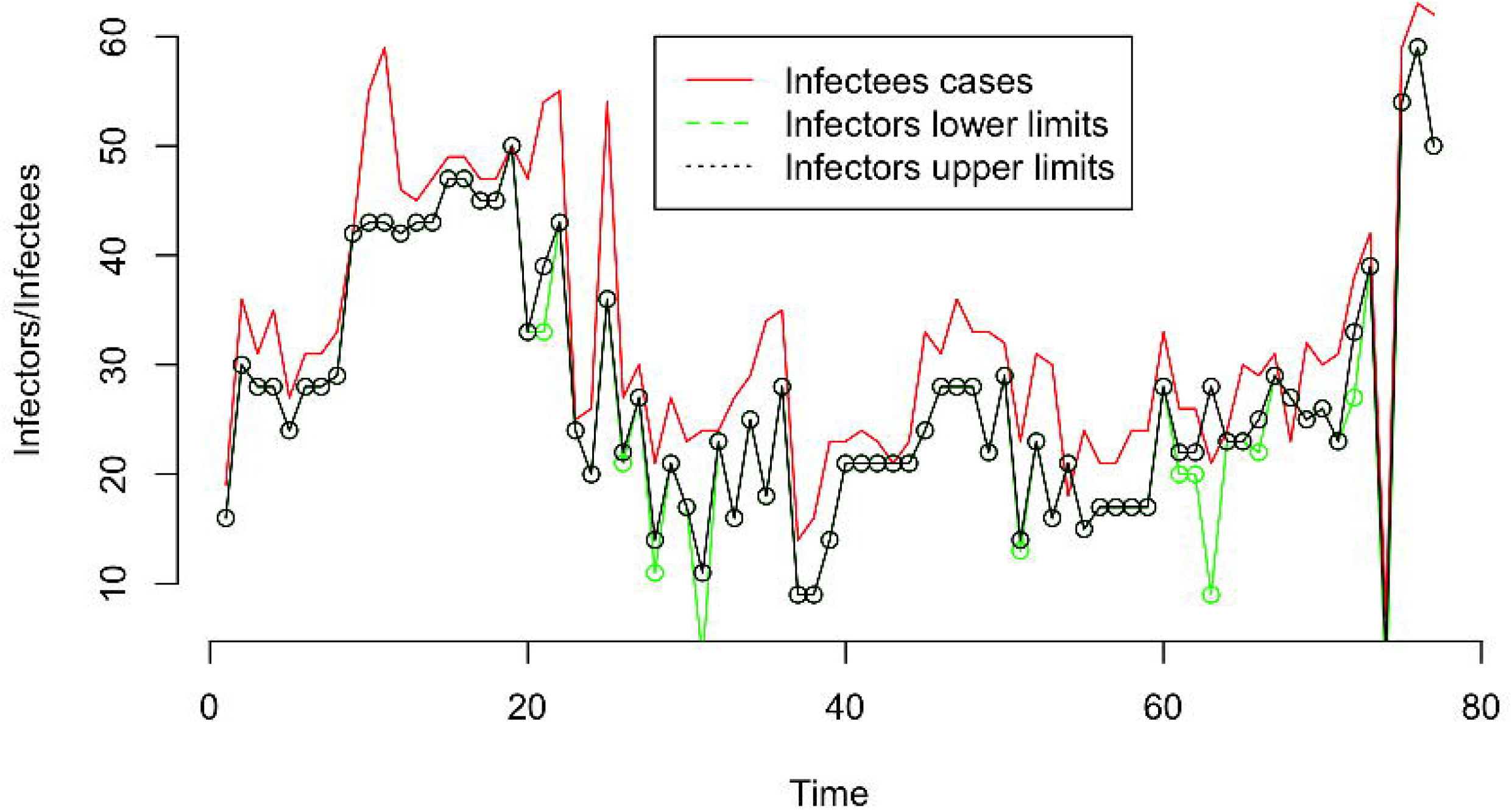

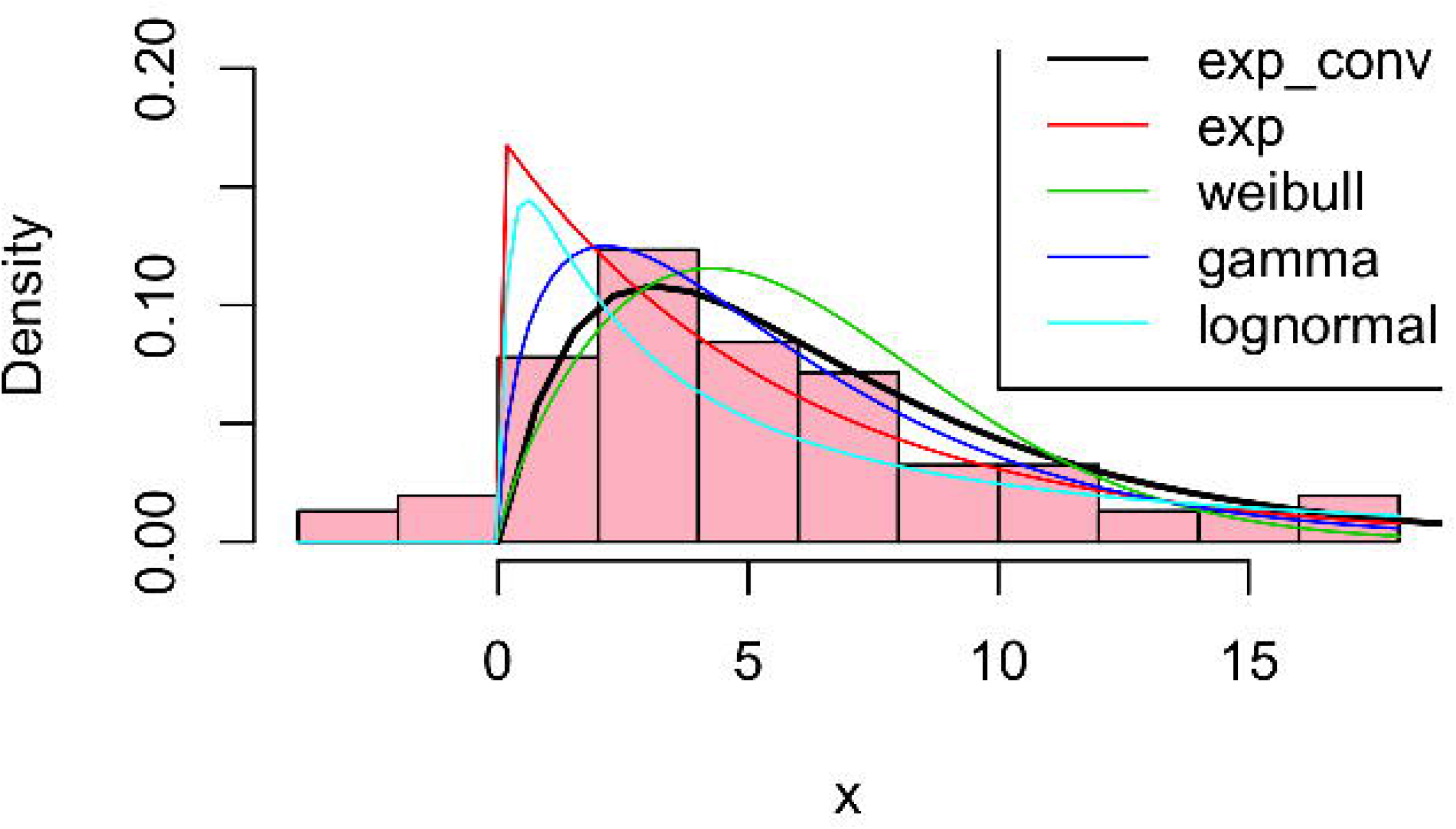

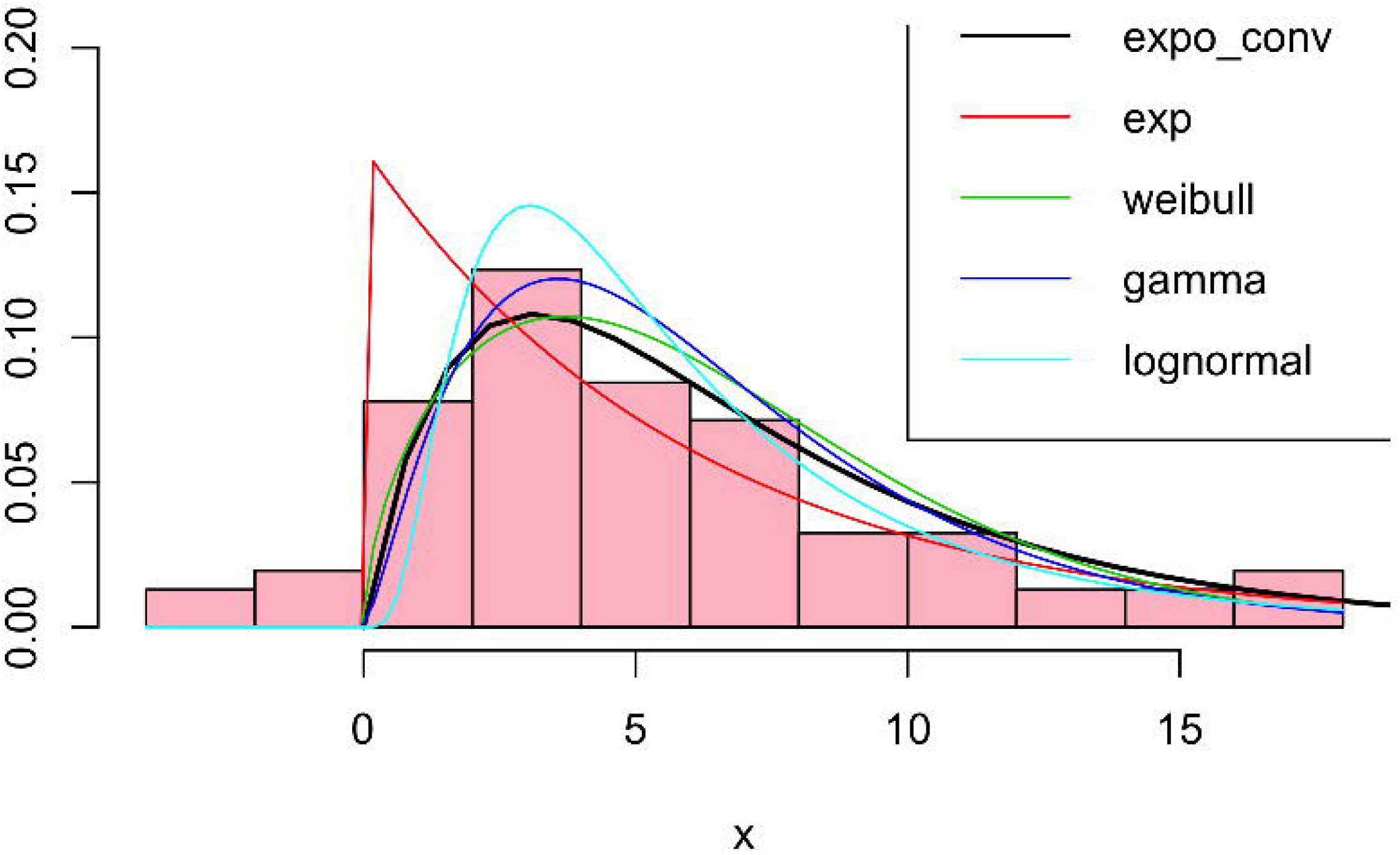

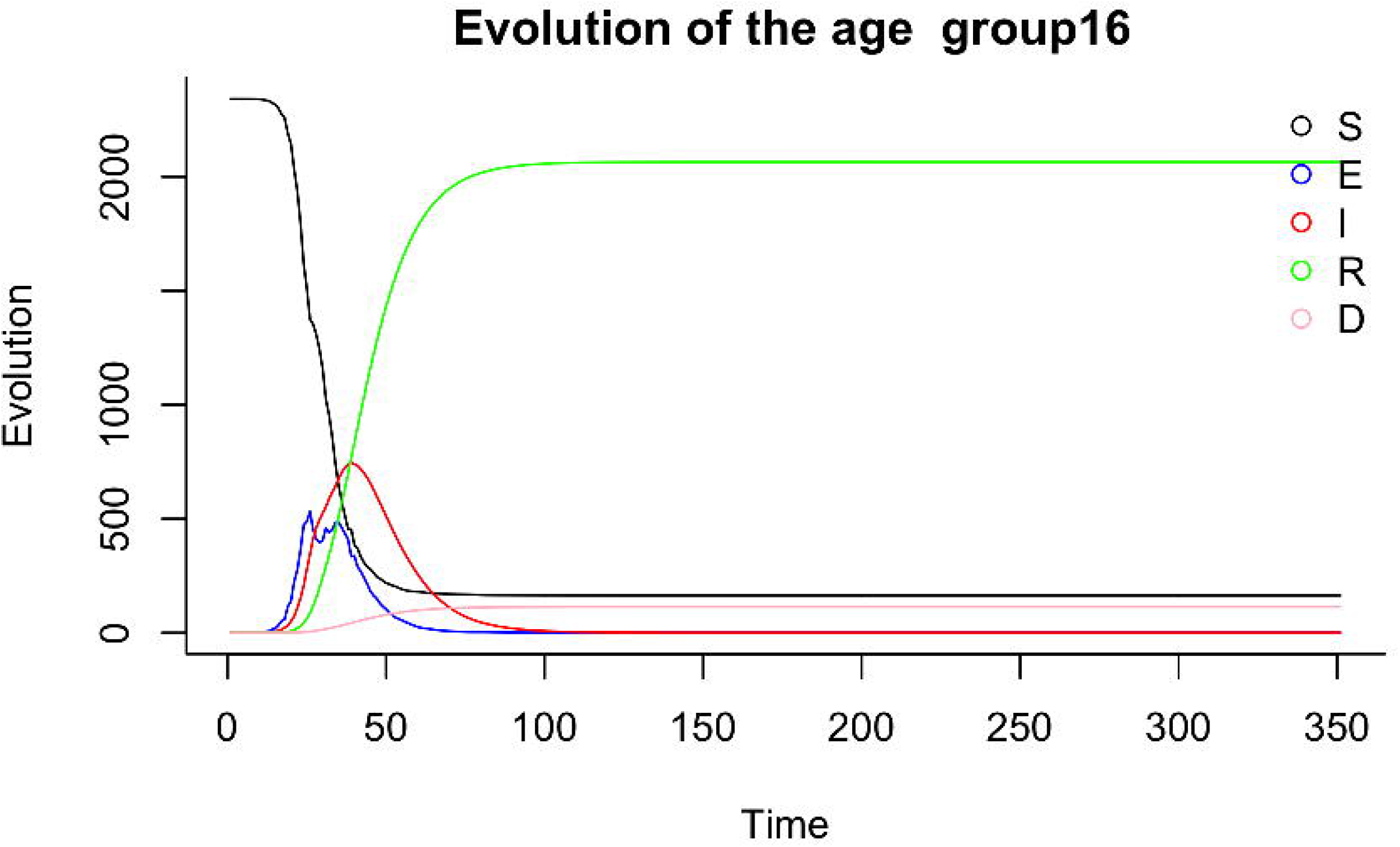

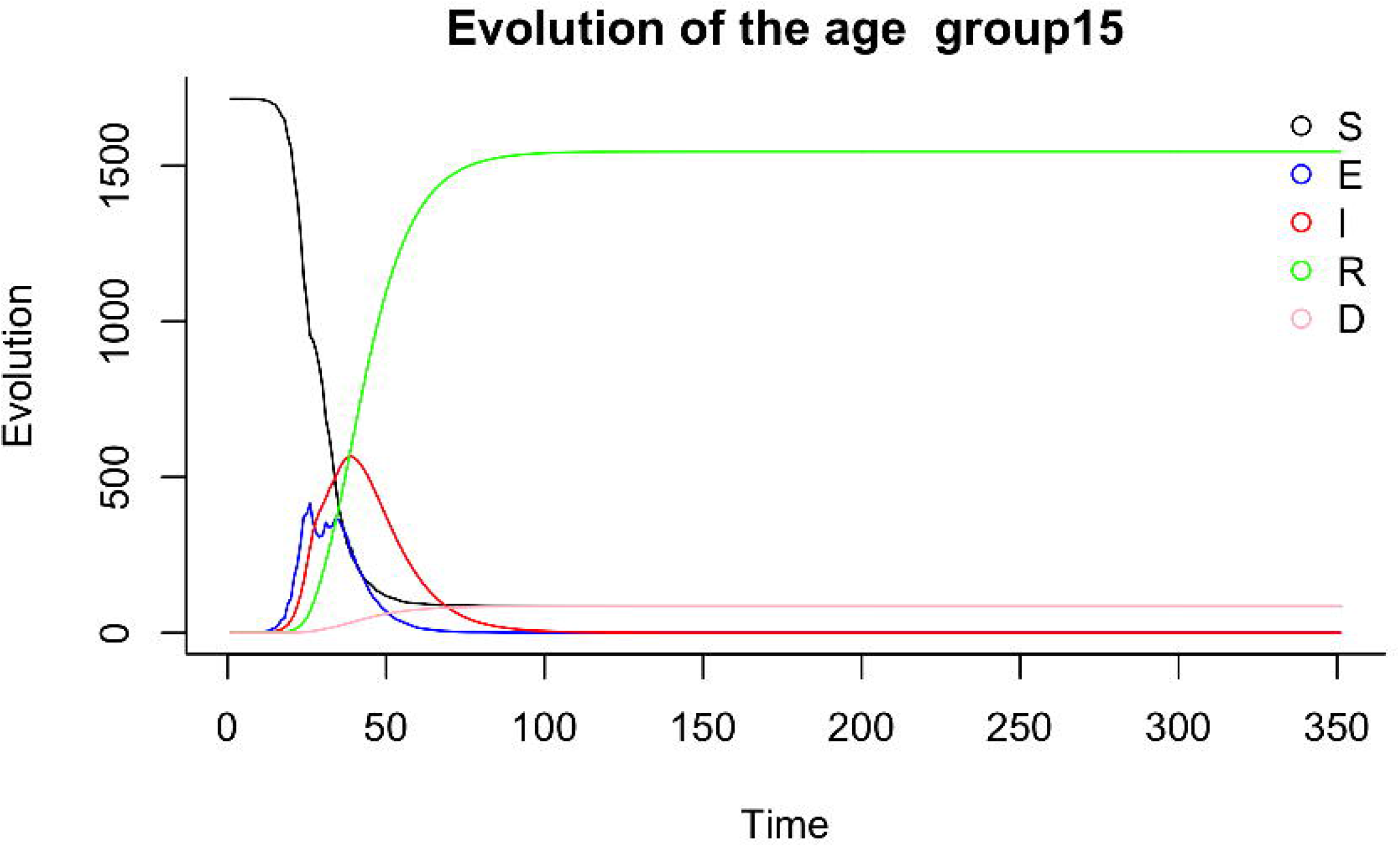

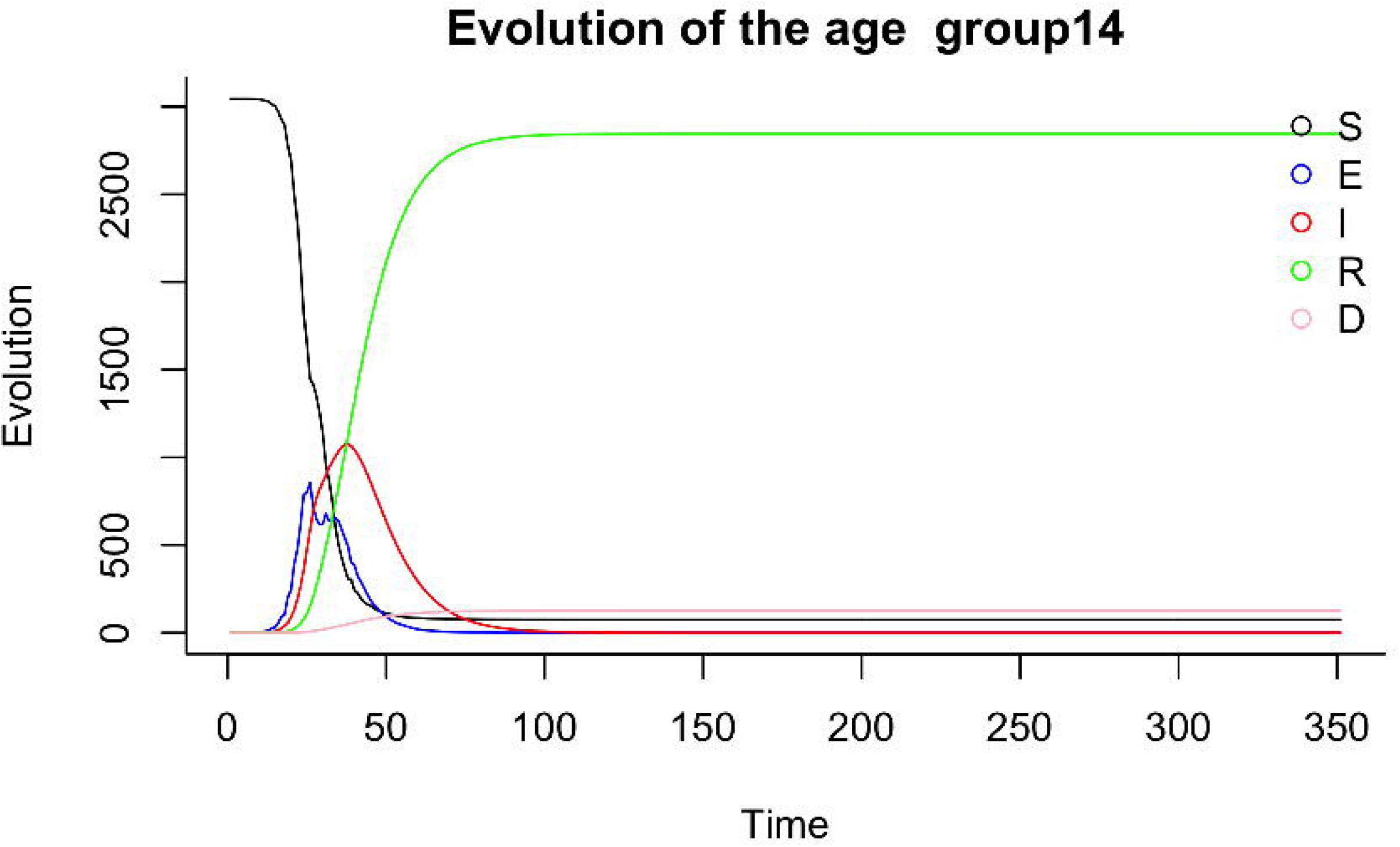

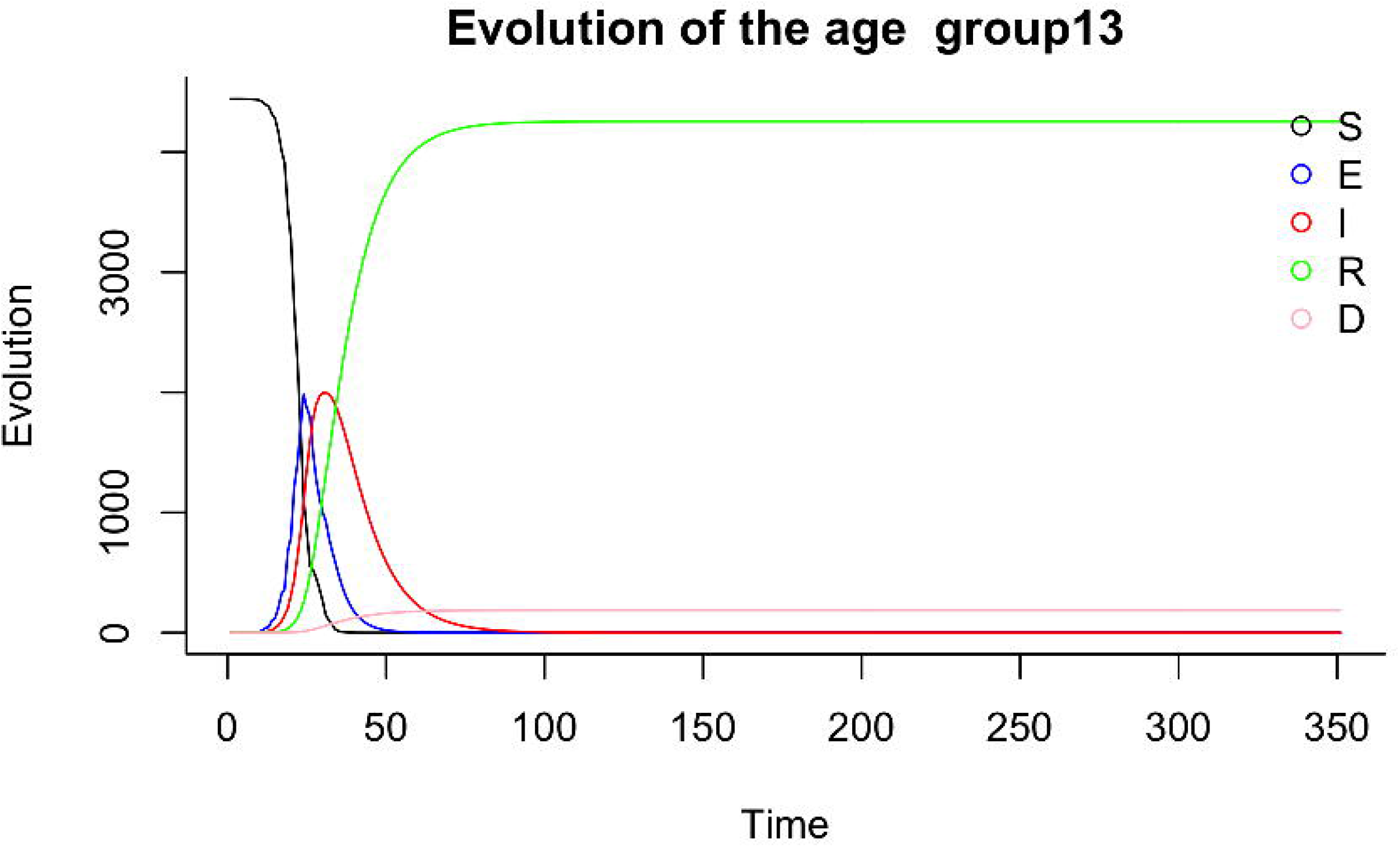

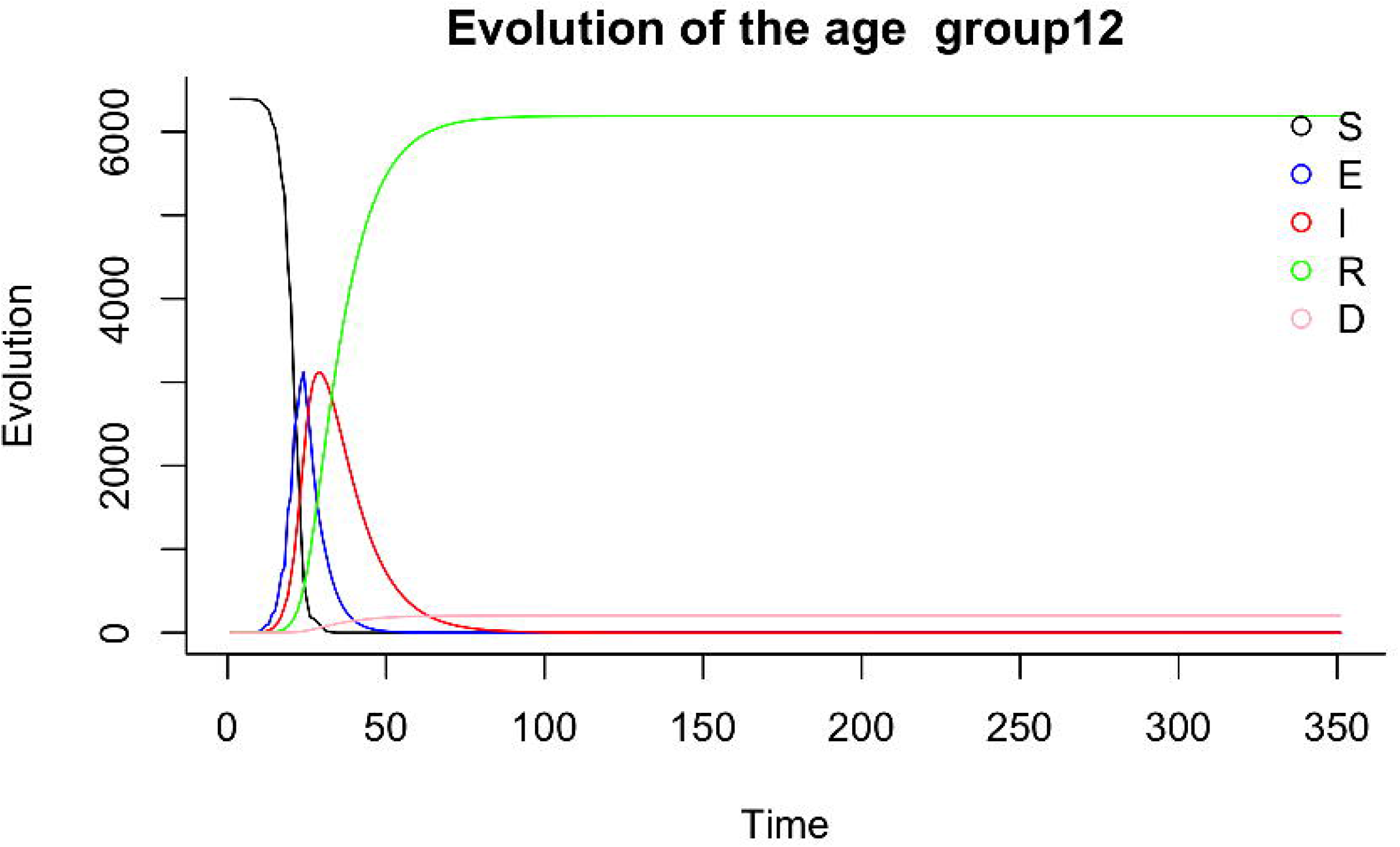

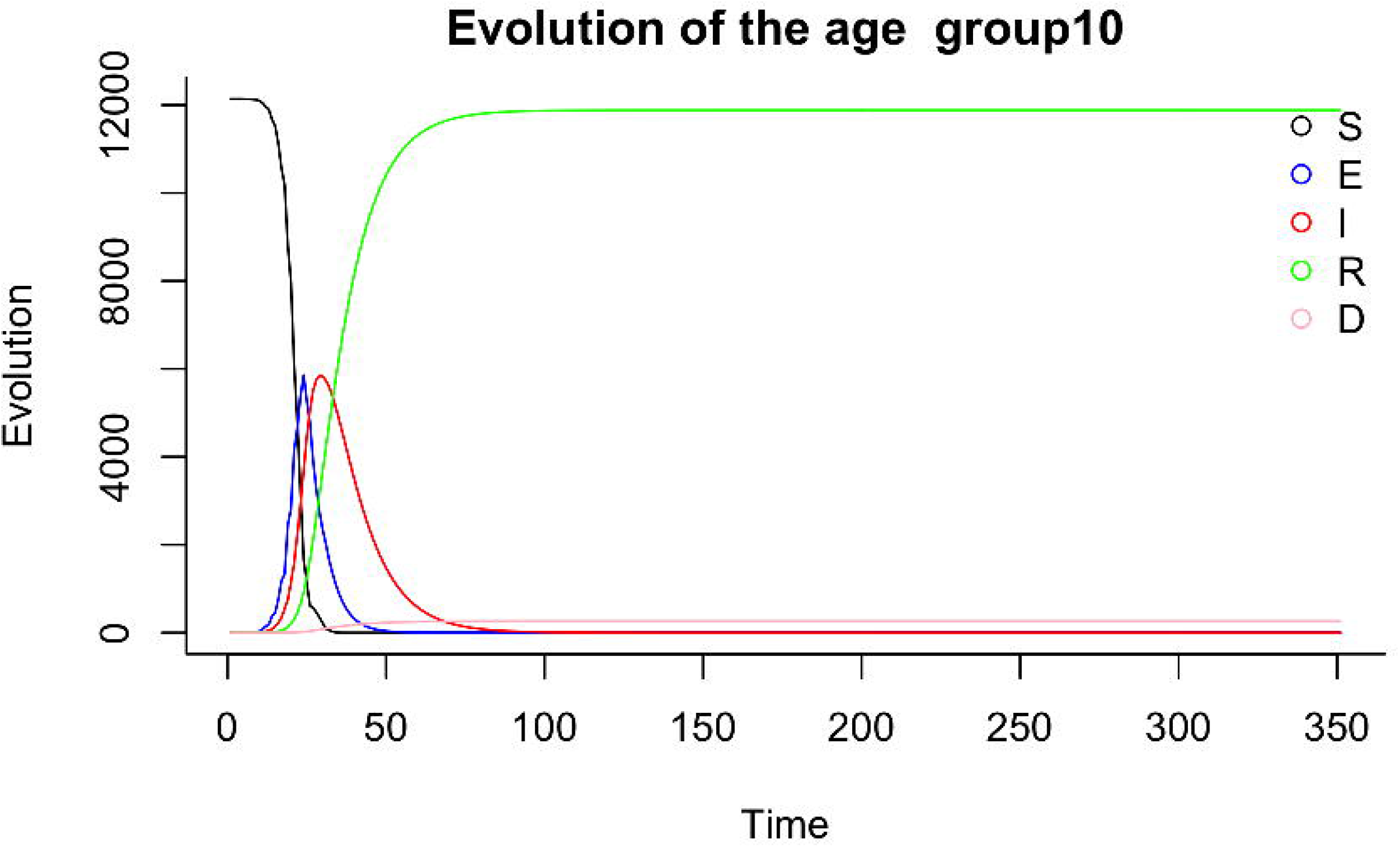

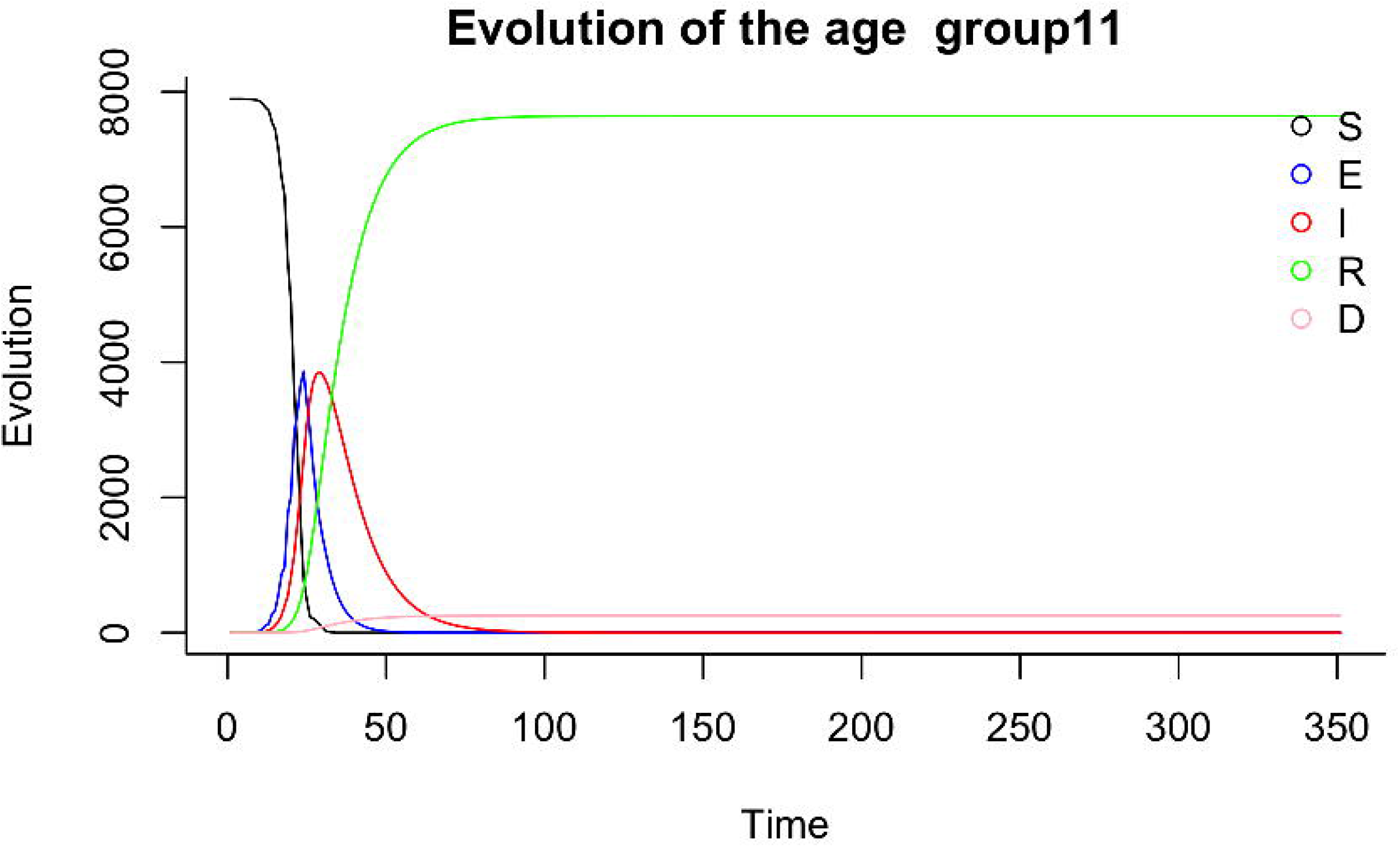

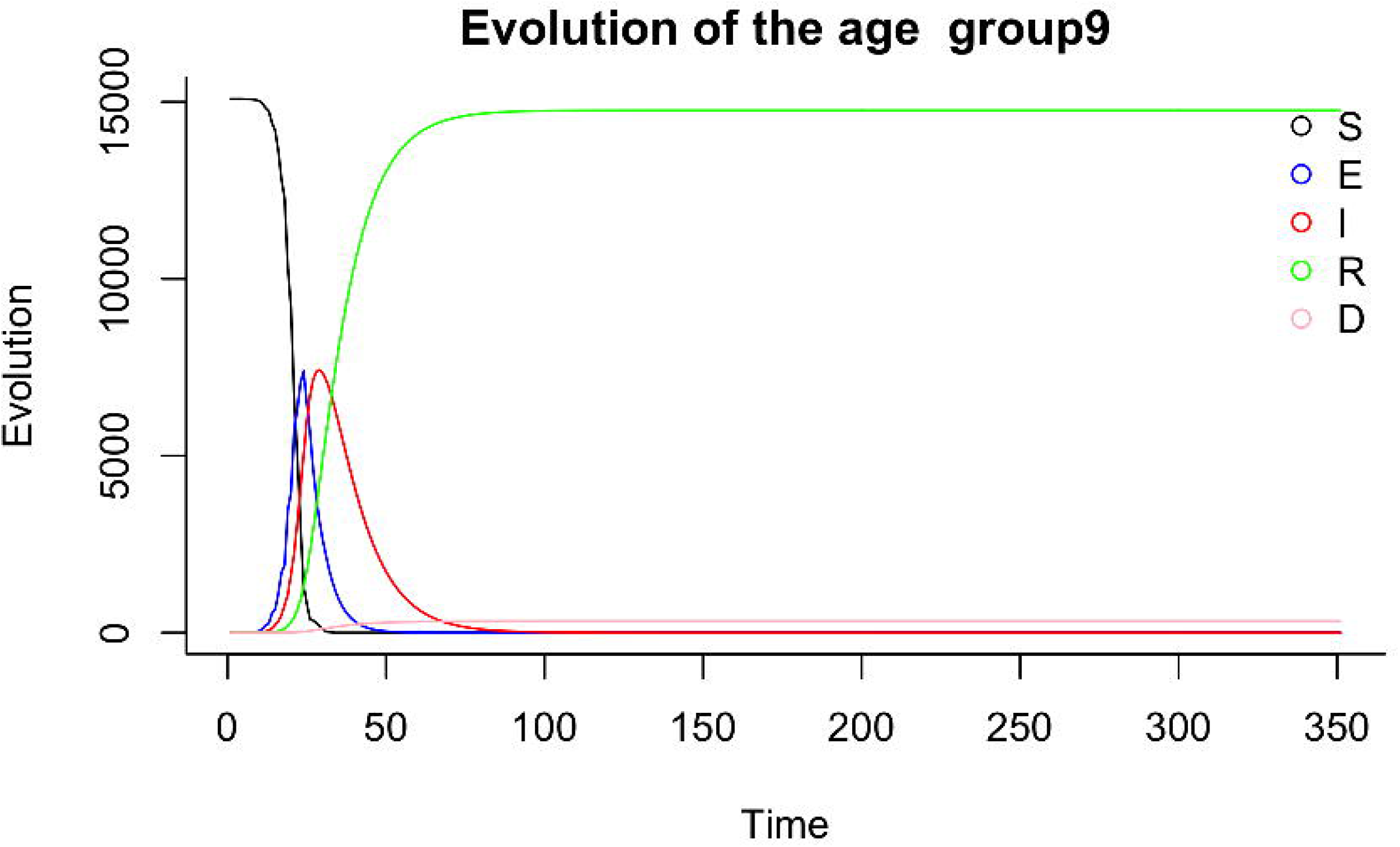

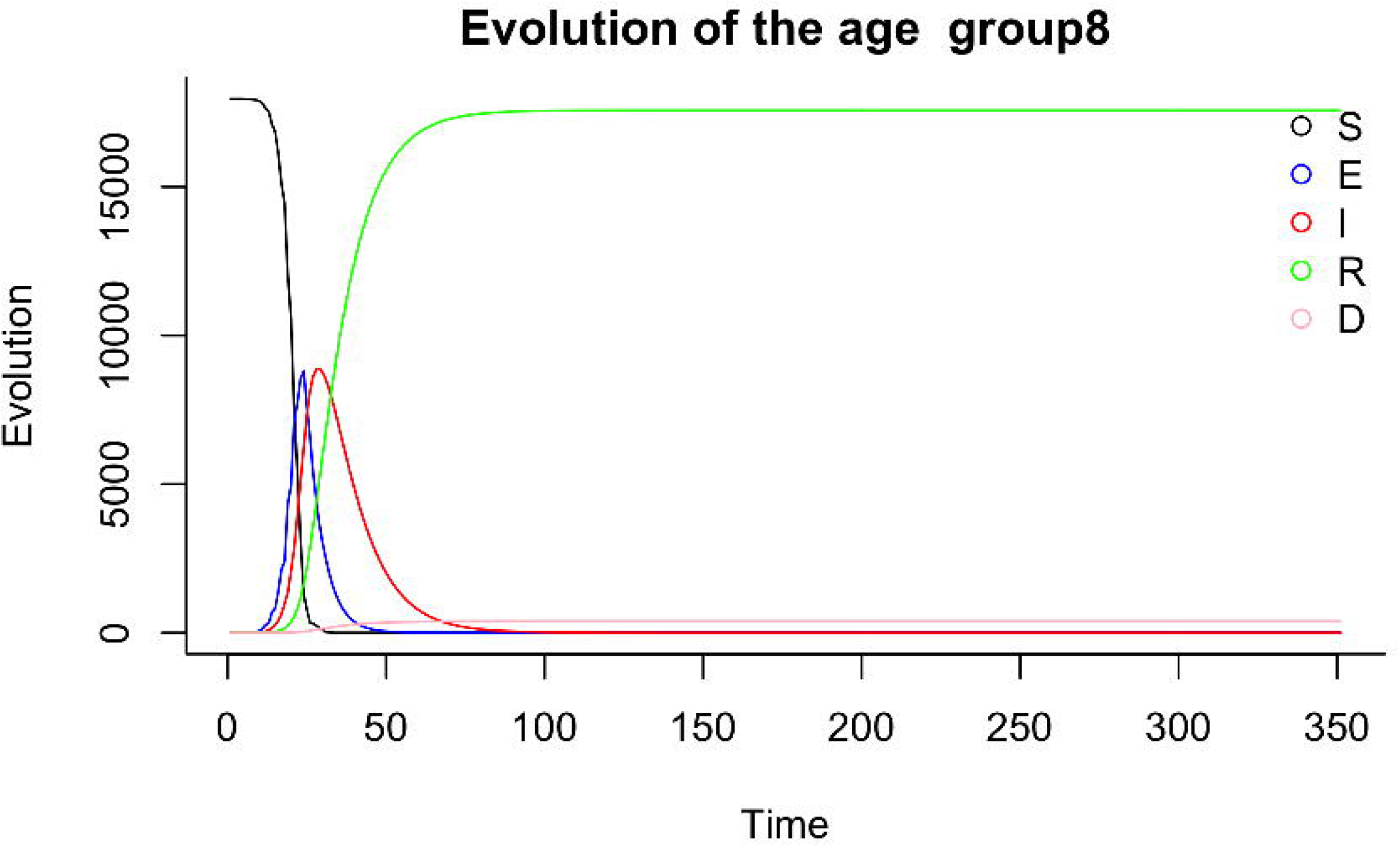

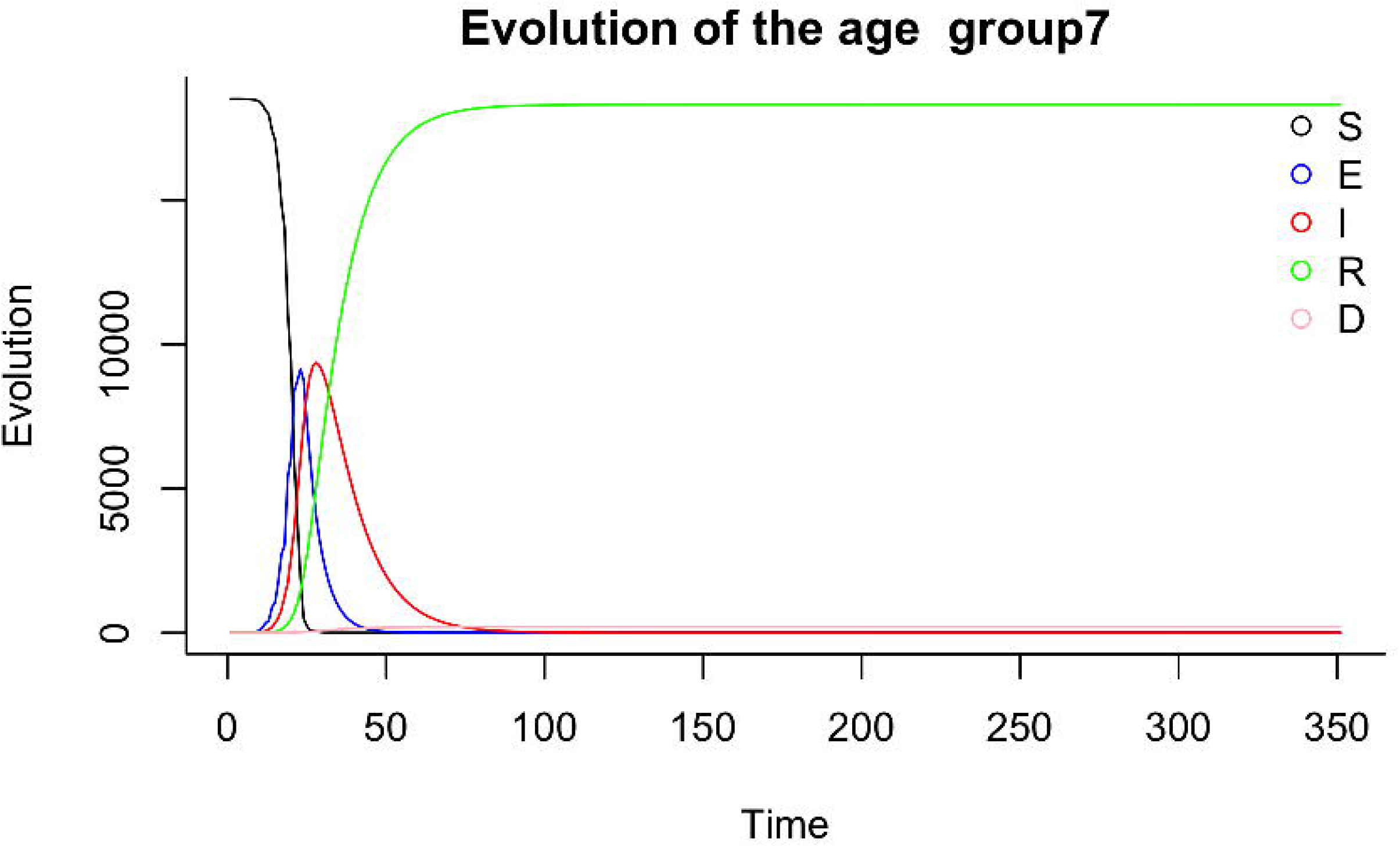

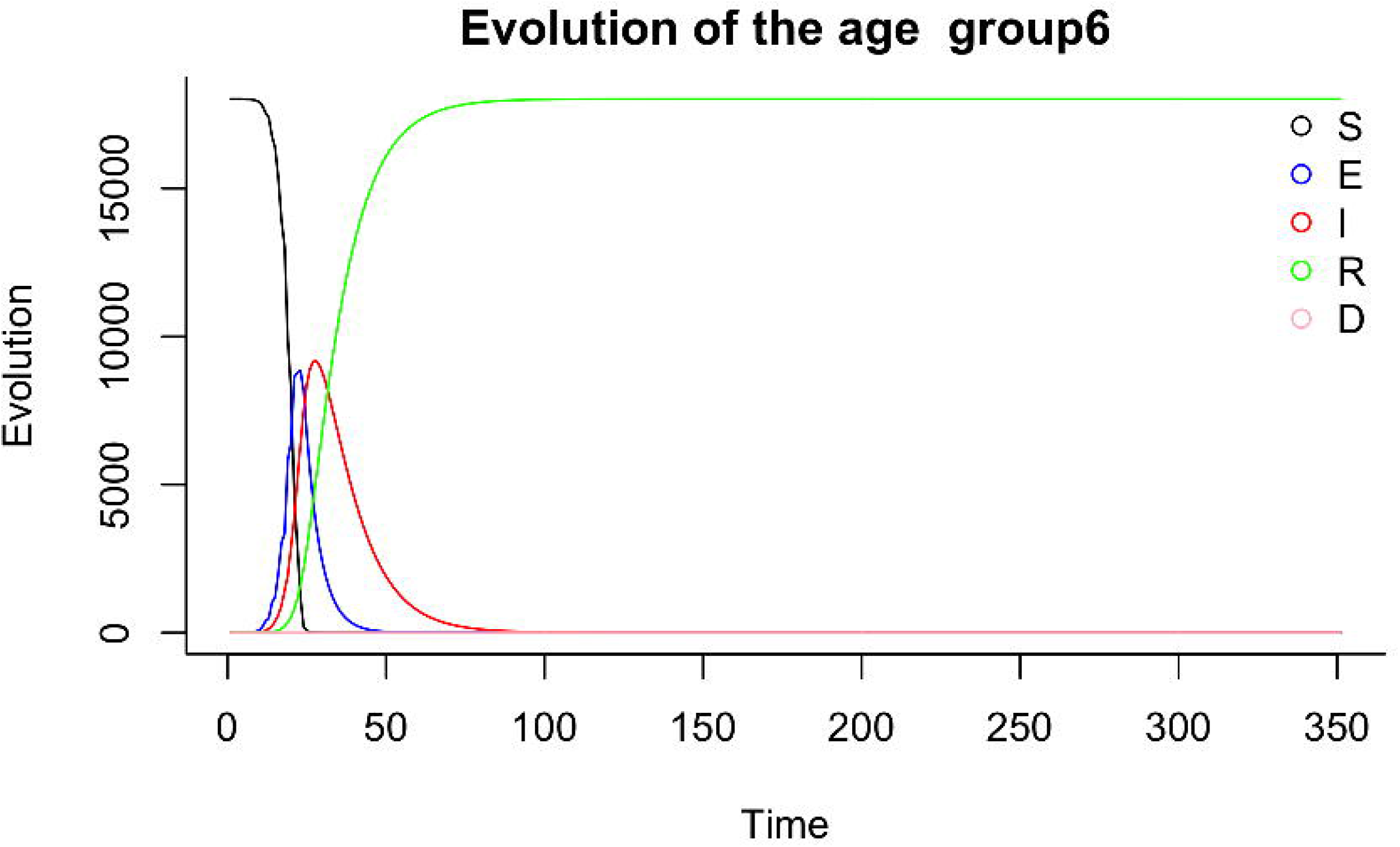

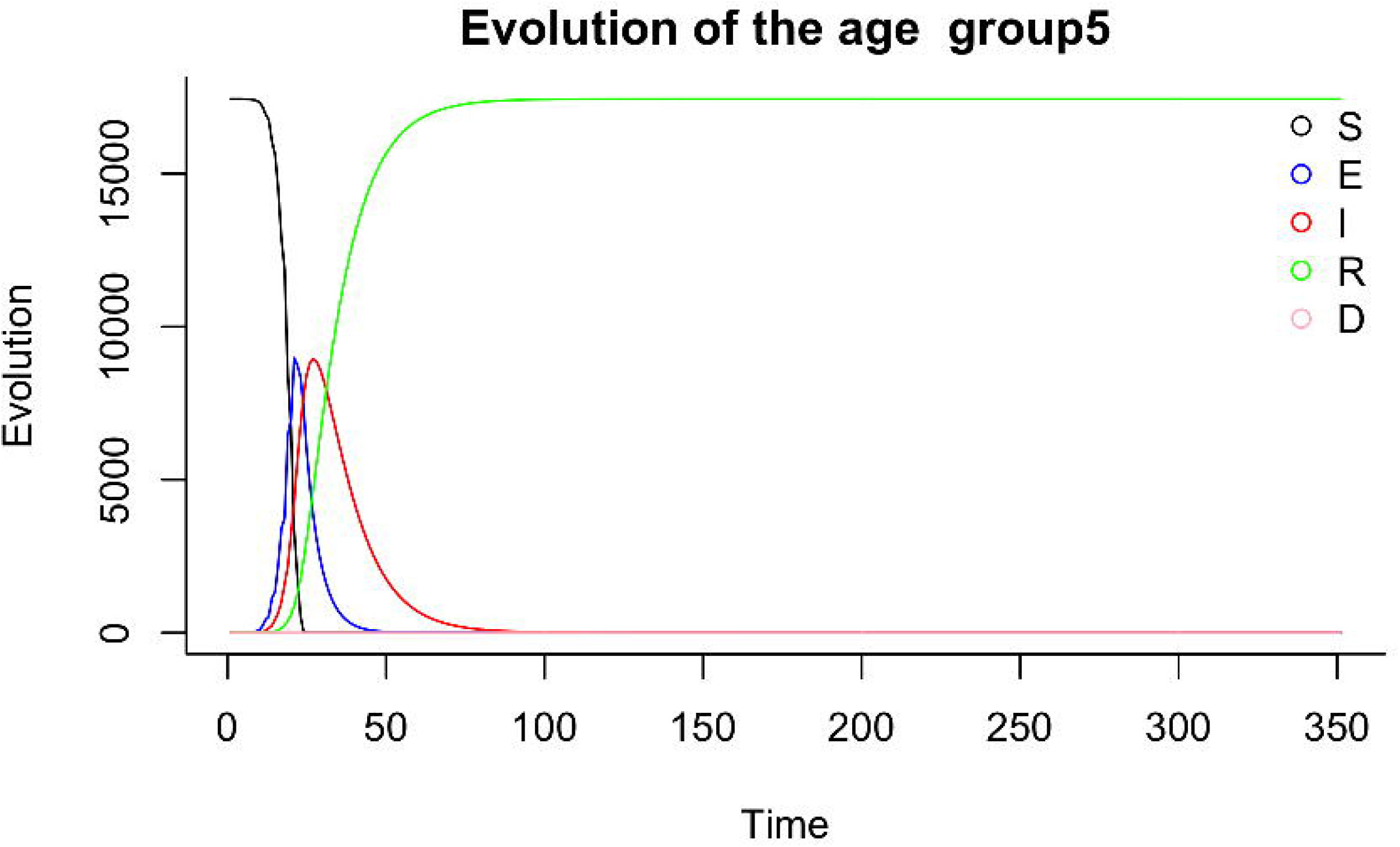

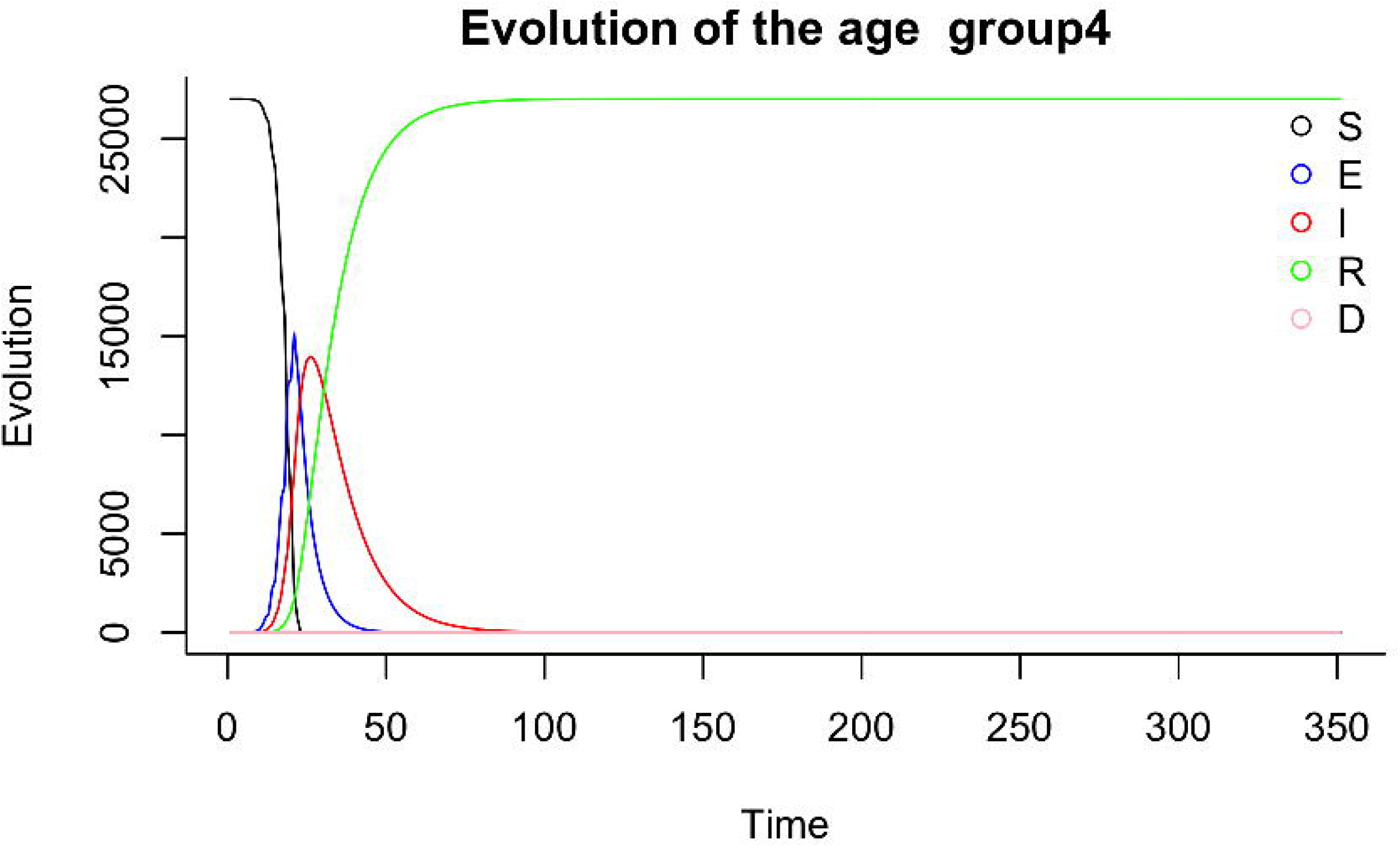

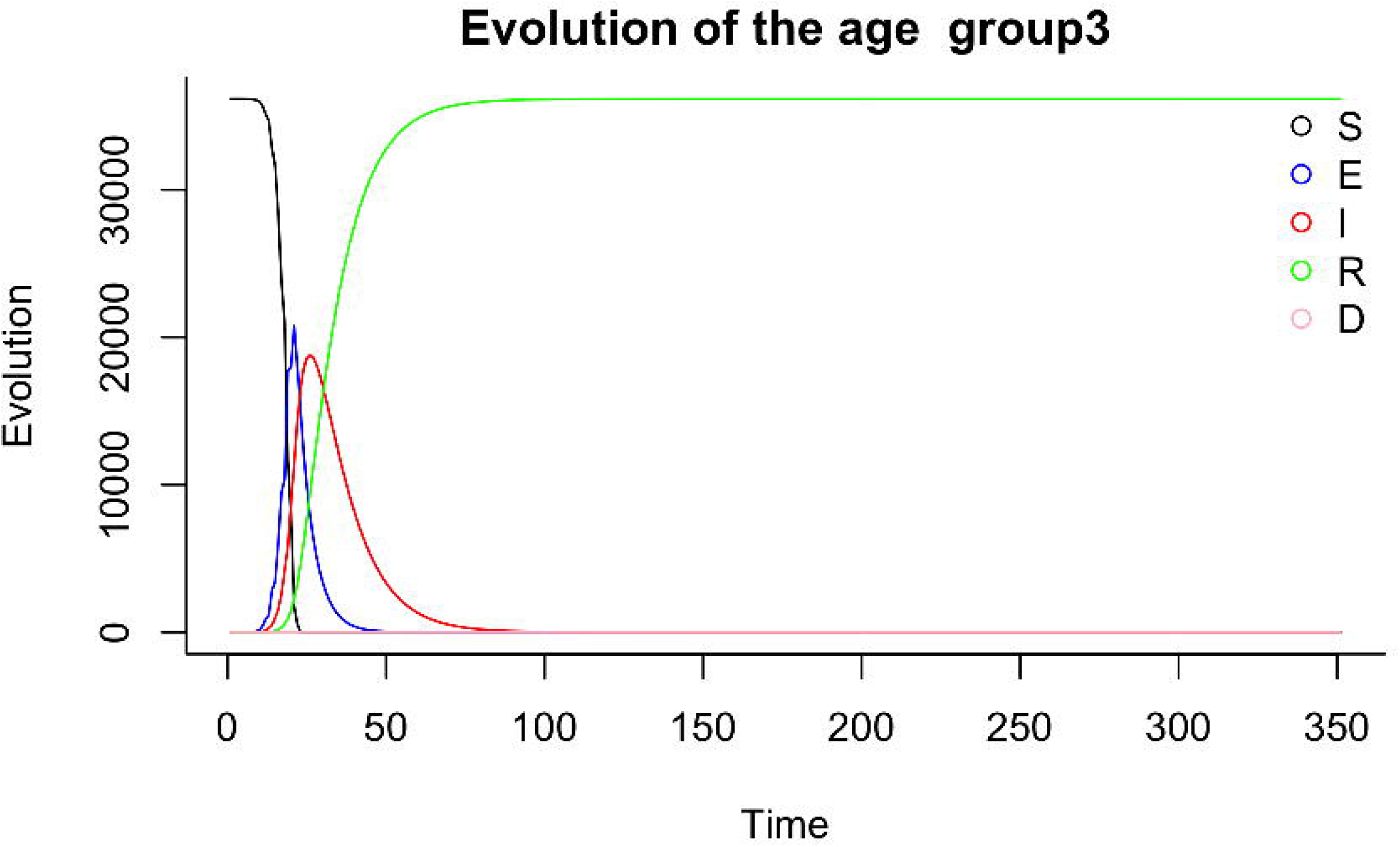

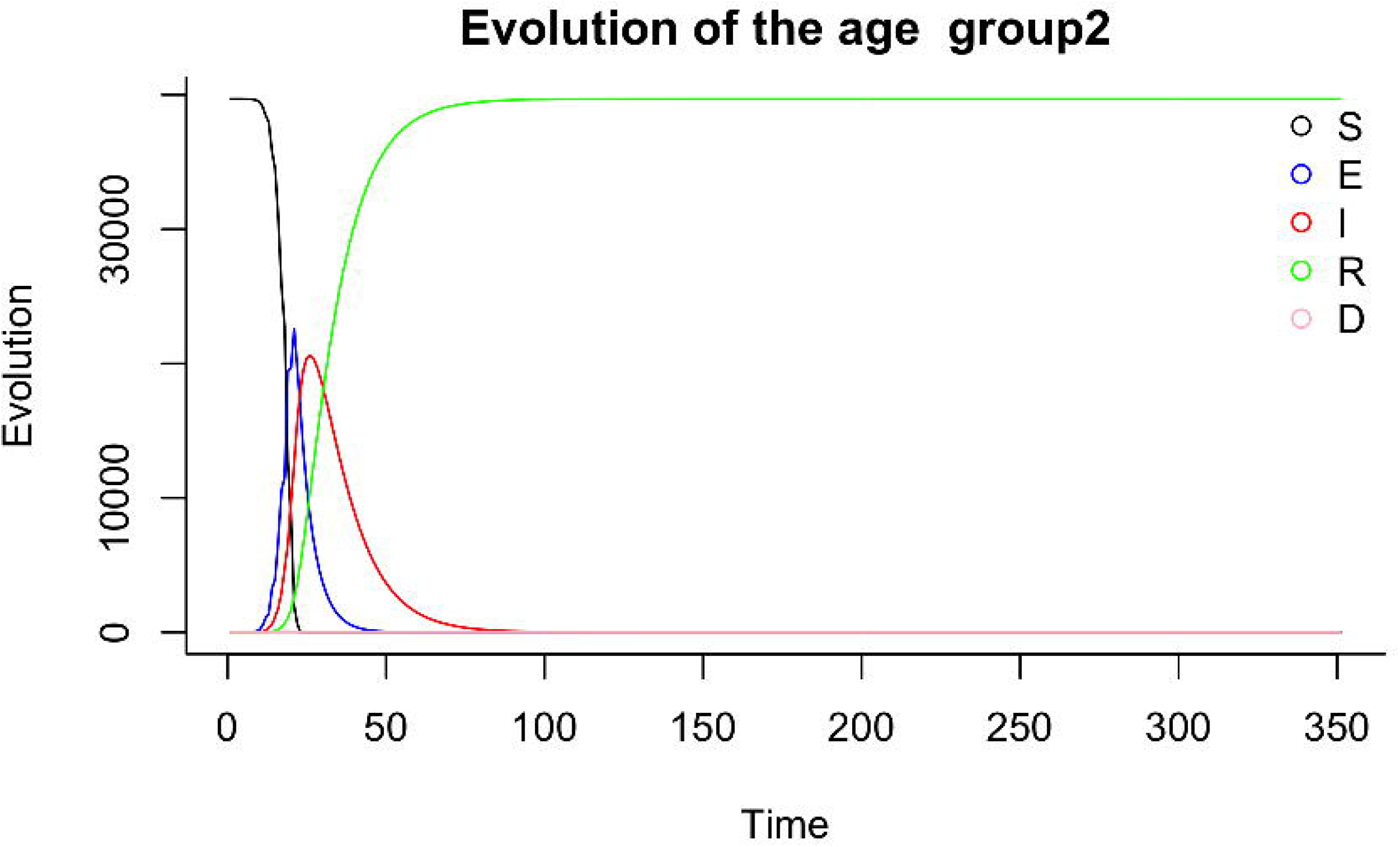

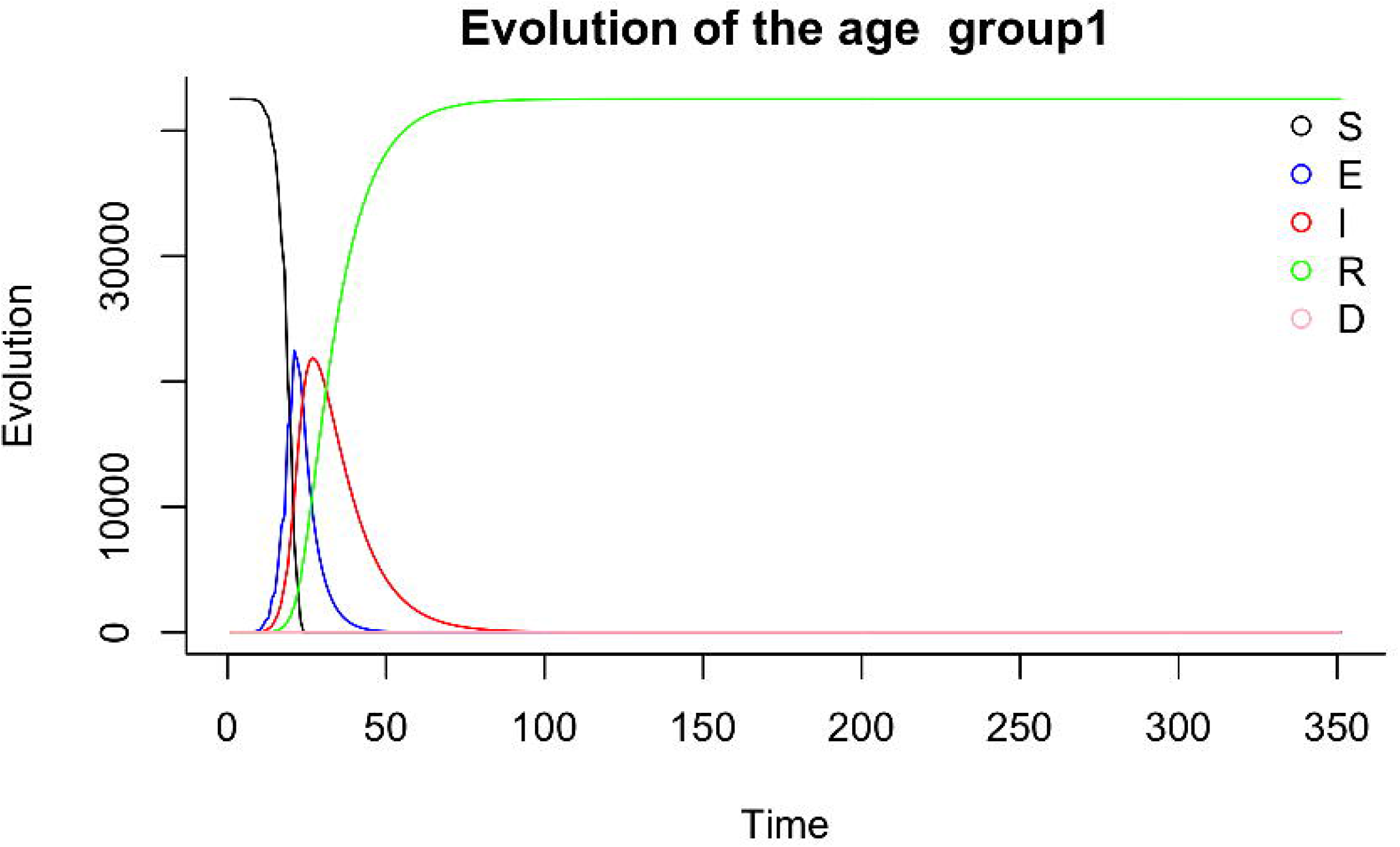

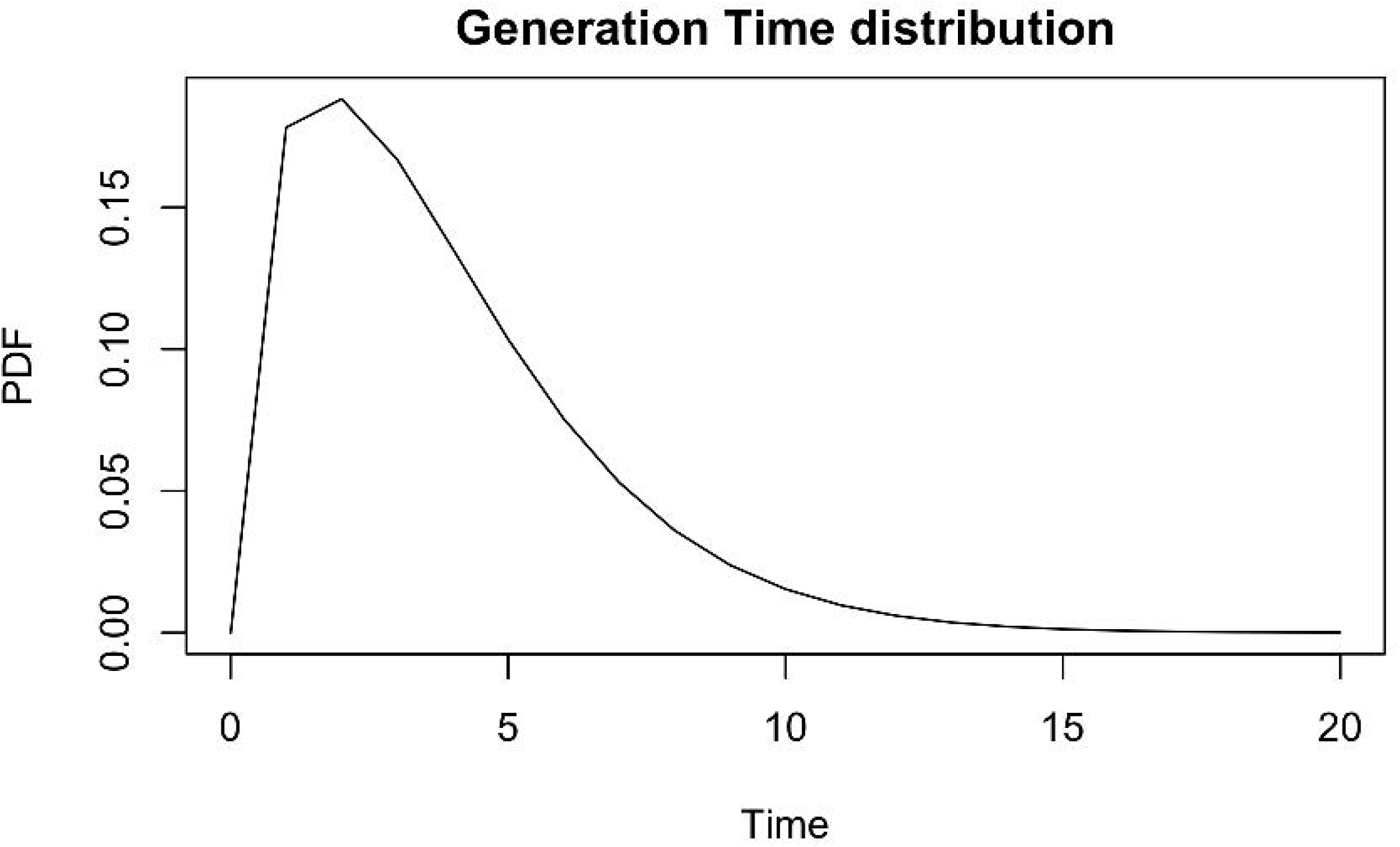

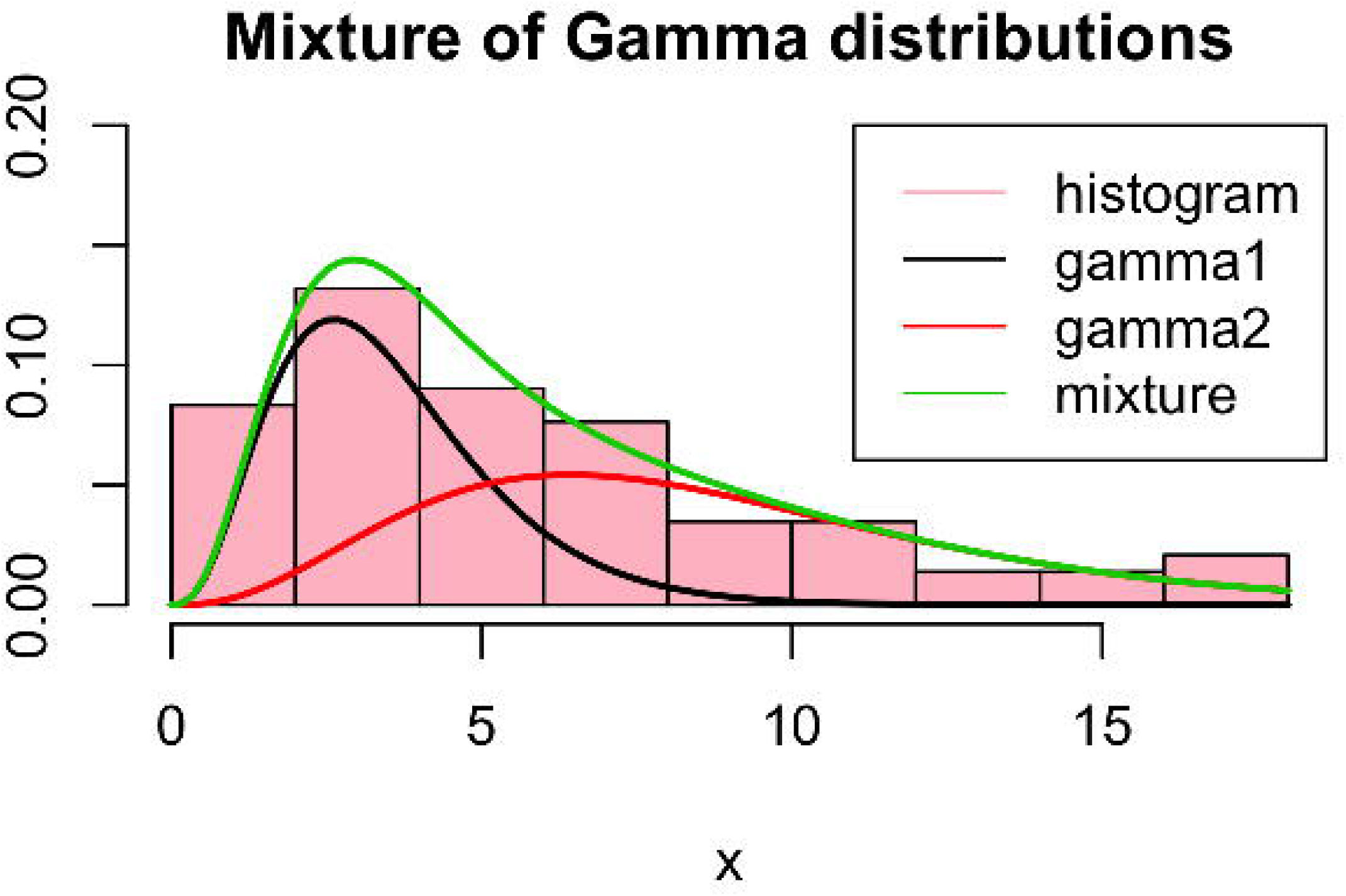

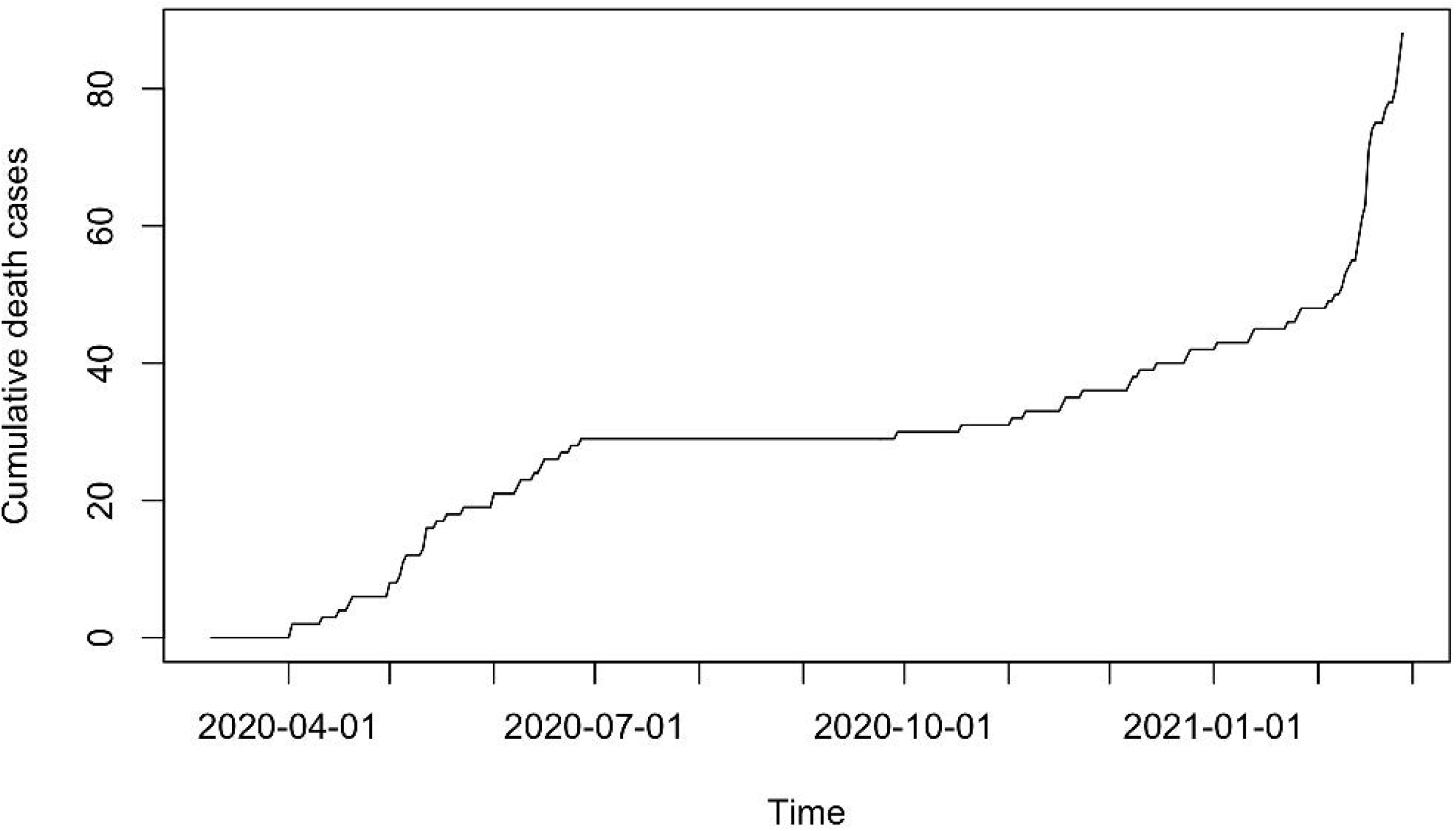

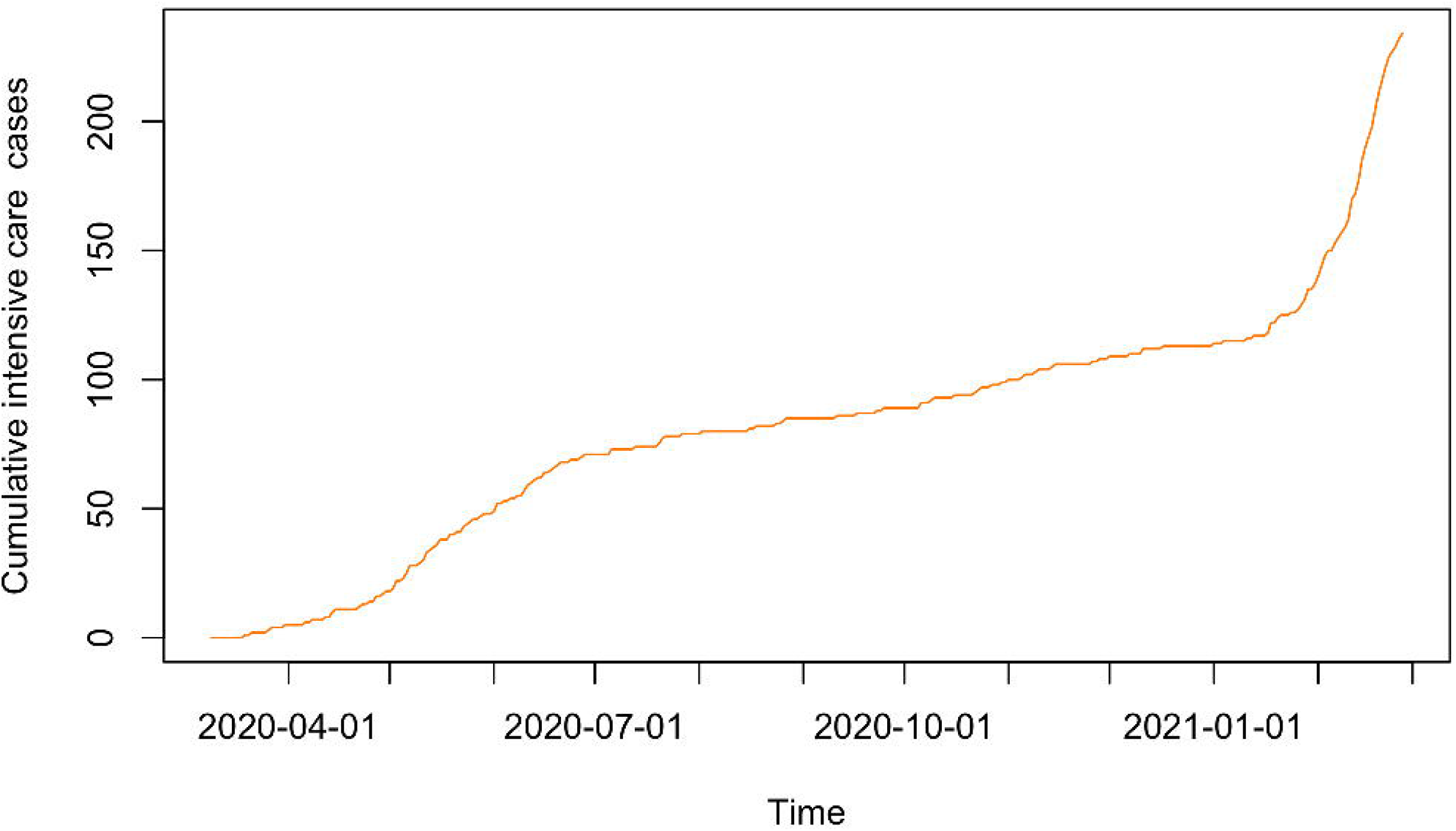

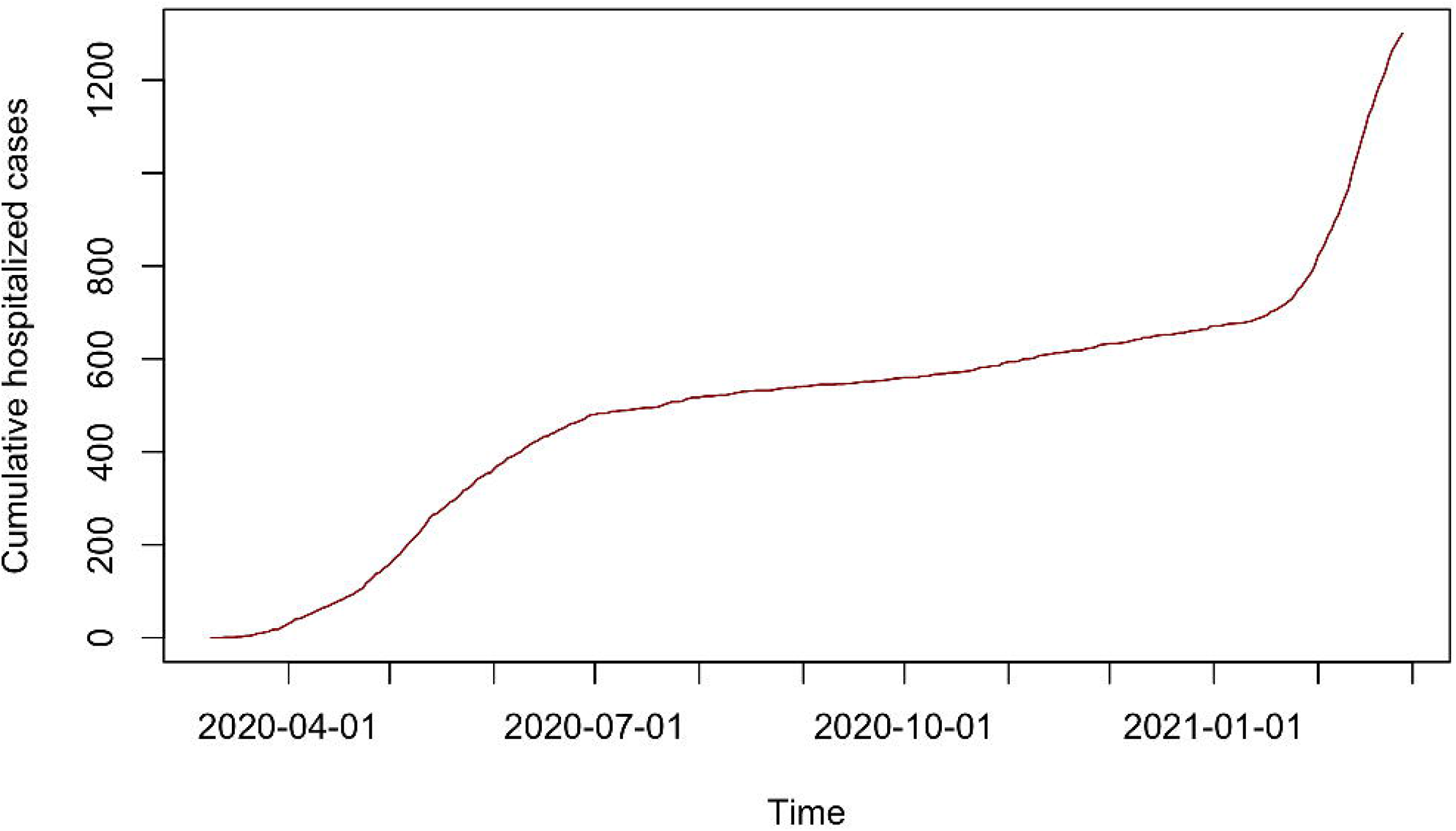

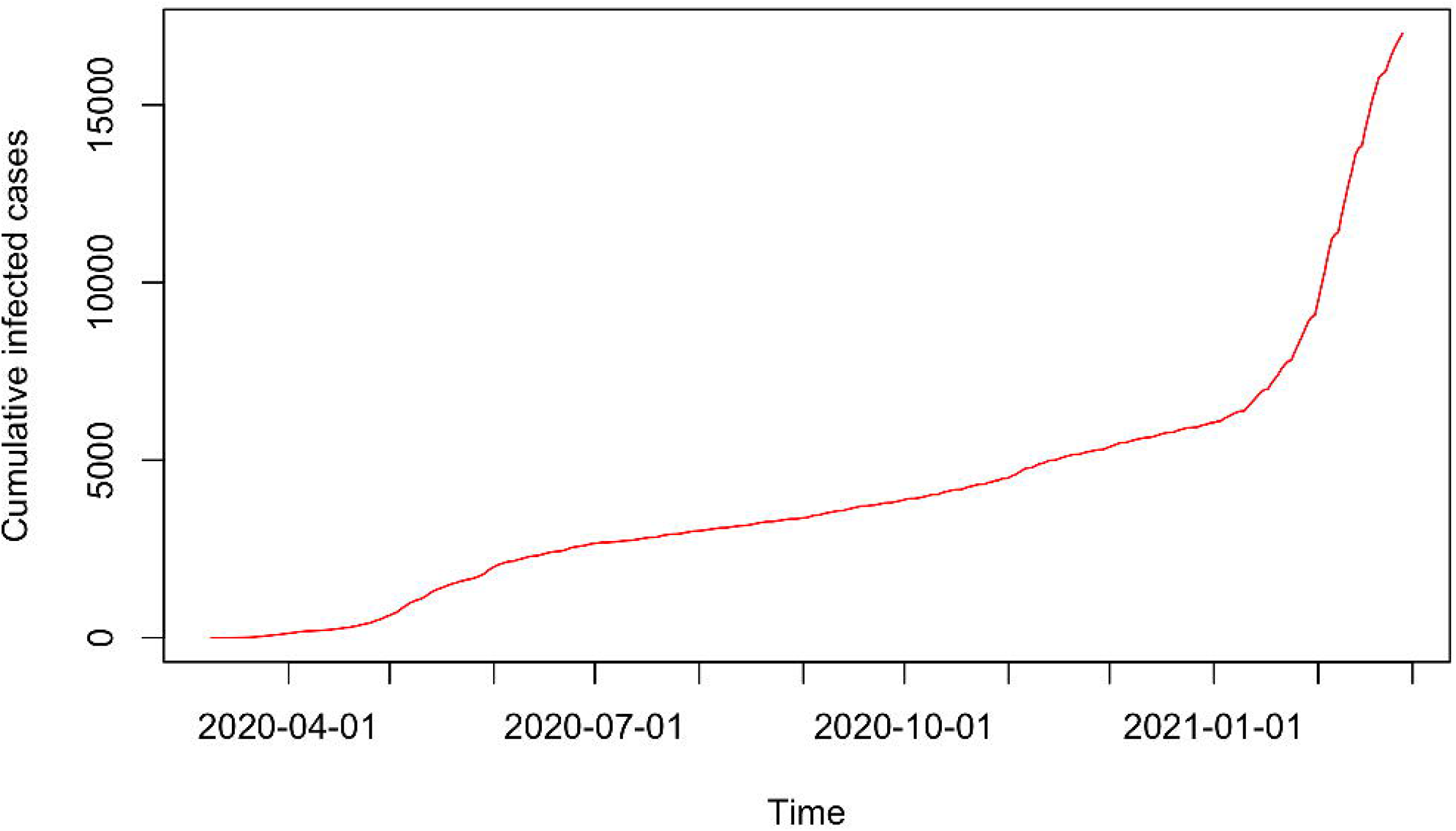

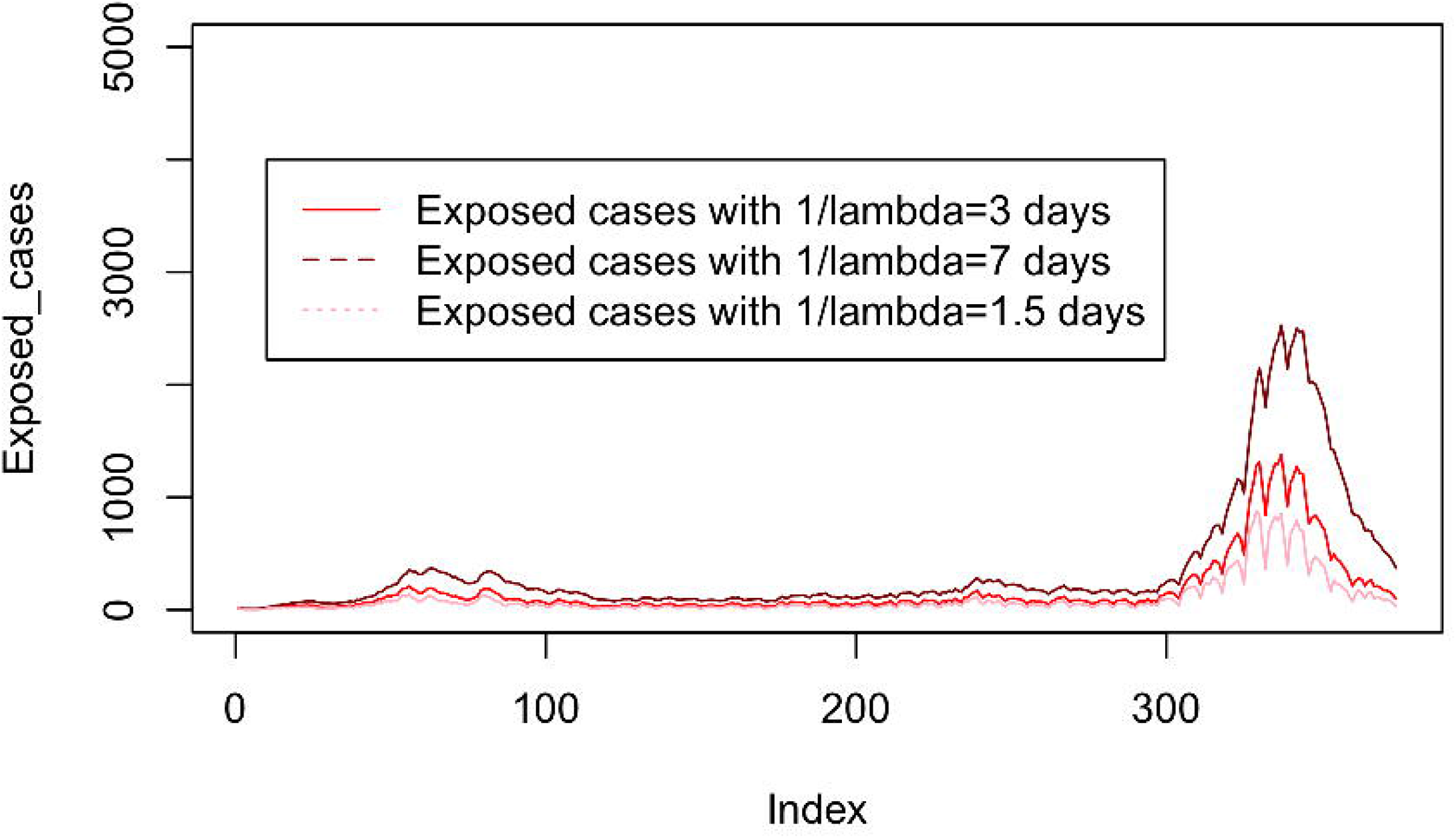

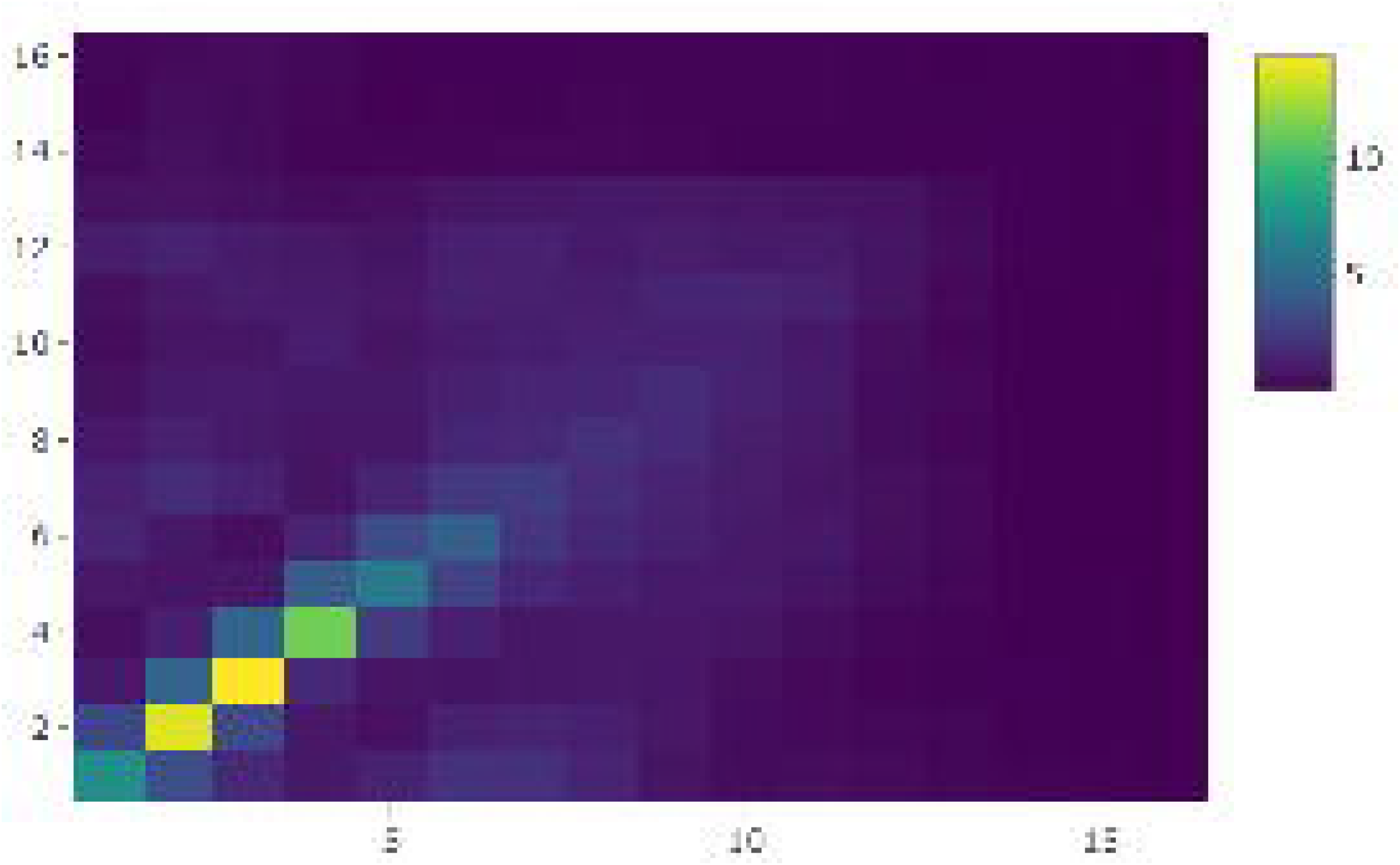

